# SARS-CoV-2 Uses CD4 to Infect T Helper Lymphocytes

**DOI:** 10.1101/2020.09.25.20200329

**Authors:** Natália S. Brunetti, Gustavo G. Davanzo, Diogo de Moraes, Allan J. R. Ferrari, Gabriela F. de Souza, Stefanie P. Muraro, Thiago L. Knittel, Vinícius O. Boldrini, Lauar B. Monteiro, João Victor Virgilio-da-Silva, Gerson S. Profeta, Natália S. Wassano, Luana N. Santos, Victor C. Carregari, Artur H. S. Dias, Flavio P. Veras, Lucas A. Tavares, Julia Forato, Ícaro Castro, Lícia C. Silva-Costa, Andre Palma, Eli Mansour, Raisa G. Ulaf, Ana F. Bernardes, Thyago A. Nunes, Luciana C. Ribeiro, Marcus V. Agrela, Maria Luiza Moretti, Lucas I. Buscaratti, Fernanda Crunfli, Raissa G. Ludwig, Jaqueline A. Gerhardt, Natália Munhoz-Alves, Ana M. Marques, Renata Sesti-Costa, Mariene R. Amorim, Daniel A. T. Texeira, Pierina L. Parise, Matheus C. Martini, Karina Bispo-dos-Santos, Camila L. Simeoni, Fabiana Granja, Virginia C. Silvestrini, Eduardo B. de Oliveira, Vitor M. Faça, Murilo Carvalho, Bianca G. Castelucci, Alexandre B. Pereira, Laís D. Coimbra, Marieli M. G. Dias, Patricia B. Rodrigues, Arilson Bernardo S. P. Gomes, Fabricio B. Pereira, Leonilda M. B. Santos, Louis-Marie Bloyet, Spencer Stumpf, Marjorie C. Pontelli, Sean P. J. Whelan, Andrei C. Sposito, Robson F. Carvalho, Andre S. Vieira, Marco A. R. Vinolo, André Damasio, Licio A. Velloso, Ana Carolina M. Figueira, Luis L. P. da Silva, Thiago M. Cunha, Helder I. Nakaya, Henrique Marques-Souza, Rafael E. Marques, Daniel Martins-de-Souza, Munir S. Skaf, José Luiz Proença-Modena, Pedro M. Moraes-Vieira, Marcelo A. Mori, Alessandro S. Farias

## Abstract

The severe acute respiratory syndrome coronavirus 2 (SARS-CoV-2) is the agent of a major global outbreak of respiratory tract disease known as coronavirus disease-2019 (COVID-19). SARS-CoV-2 infects mainly lungs and may cause several immune-related complications, such as lymphocytopenia and cytokine storm, which are associated with the severity of the disease and predict mortality^1,2^. The mechanism by which SARS-CoV-2 infection may result in immune system dysfunction is still not fully understood. Here we show that SARS-CoV-2 infects human CD4^+^ T helper cells, but not CD8^+^ T cells, and is present in blood and bronchoalveolar lavage T helper cells of severe COVID-19 patients. We demonstrated that SARS-CoV-2 spike glycoprotein (S) directly binds to the CD4 molecule, which in turn mediates the entry of SARS-CoV-2 in T helper cells. This leads to impaired CD4 T cell function and may cause cell death. SARS-CoV-2-infected T helper cells express higher levels of IL-10, which is associated with viral persistence and disease severity. Thus, CD4-mediated SARS-CoV-2 infection of T helper cells may contribute to a poor immune response in COVID-19 patients.

---

Coronavirus disease 2019 (COVID-19) has rapidly spread across the globe^3,4^, being declared a pandemic by the World Health Organization (WHO) on March 11^th^, 2020. COVID-19 has caused millions of deaths around the world. Most of the deaths are associated with acute pneumonia, cardiovascular complications, and organ failure due to hypoxia, exacerbated inflammatory responses and widespread cell death^1,5^. Individuals that progress to the severe stages of COVID-19 manifest marked alterations in the immune response, characterized by reduced overall protein synthesis, cytokine storm, lymphocytopenia and T cell exhaustion^6–8^. In addition to these acute effects on the immune system, a considerable proportion of infected individuals present low titers of neutralizing antibodies^9,10^. Moreover, the levels of antibodies against SARS-CoV-2 decay rapidly after recovery in part of the infected individuals^11^, suggesting that SARS-CoV-2 infection may exert profound and long-lasting complications to adaptive immunity. Recently, it has been shown that SARS-CoV-2 is able to infect lymphocytes^12,13^. In this context, it is urgent to characterize the replicative capacity and the effects of SARS-CoV-2 replication in different immune cells, especially those involved with the formation of immunological memory and effective adaptive response, such as CD4^+^ T lymphocytes.

In what has been proposed to be the canonical mechanism of SARS-CoV-2 infection, the spike glycoprotein of SARS-CoV-2 (sCoV-2) binds to the host angiotensin-converting enzyme 2 (ACE2), after which it is cleaved by TMPRSS2^14^. While TMPRSS2 is ubiquitously expressed in human tissues (**Extended Data Fig. 1**), ACE2 is mainly expressed in epithelial and endothelial cells, as well as in the kidney, testis and small intestine (**Extended Data Fig.1**). Still, a wide variety of cell types are potentially infected by SARS-CoV-2^15–18^, even though some of these cells express very low levels of ACE2. We showed this is the case for lymphocytes (**Extended Data Fig. 2**). These findings suggest that either SARS-CoV-2 uses alternative mechanisms to enter these cells or that auxiliary molecules at the plasma membrane may promote infection by stabilizing the virus until it interacts with ACE2. In agreement with the latter, binding of sCoV-2 to certain cell surface proteins facilitates viral entry^19,20^.

Since the structures of the spike of SARS-CoV-1 (sCoV-1) and the sCoV-2 proteins are similar^21,22^, we used the P-HIPSTer algorithm to uncover human proteins that putatively interact with spike^23^. Seventy-one human proteins were predicted to interact with sCoV-1 (**Extended Data Fig. 3**). We then cross-referenced the proteins with five databases of plasma membrane proteins to identify the ones located on the cell surface (**see Methods for details**). CD4 was the only protein predicted to interact with sCoV-1 that appeared in all five databases (**Extended Data Fig.3**). CD4 is expressed mainly in T helper lymphocytes and has been shown to be the co-receptor to HIV^24^. Since CD4^+^ T lymphocytes orchestrate innate and adaptive immune response^25,26^, infection of CD4^+^ T cells by SARS-CoV-2 could explain lymphocytopenia and dysregulated inflammatory response in severe COVID-19 patients. Moreover, from an evolutionary perspective, infection of CD4^+^ T cells represents an effective mechanism for viruses to escape the immune response^27^.

To test whether human primary T cells are infected by SARS-CoV-2, we purified CD3^+^CD4^+^ and CD3^+^CD8^+^ T cells from the peripheral blood of non-infected healthy controls/donors (HC), incubated these cells with SARS-CoV-2 for 1h, and washed them the three times with PBS. The viral load was measured 24h post-infection. We were able to detect SARS-CoV-2 RNA in CD4^+^ T cells, but not CD8^+^ T cells (**Fig. 1A, Extended Data Fig. 4A**). To confirm the presence of SARS-CoV-2 in the cells, we performed *in situ* hybridization using probes against the viral RNA-dependent RNA polymerase (RdRp) gene, immunofluorescence for sCoV-2 using antibodies against spike protein and transmission electron microscopy (**Fig. 1B and 1C**). In parallel, we infected primary CD4^+^ T cells with the VSV-mCherry-SARS-CoV-2 pseudotype virus (**Extended Data Fig. 4B**). All approaches confirmed that SARS-CoV-2 infects CD4^+^ T cells. Notably, the amount of SARS-CoV-2 RNA remains roughly stable until 48h post infection (**Fig. 1D**). Of note, although most of the data presented here was generated using the ancient SARS-CoV-2 B lineage, CD4^+^ T cells were also infected by the P.1 (gamma) variant (**Extended Data Fig. 4C**). Furthermore, we identified the presence of the negative strand (antisense) of SARS-CoV-2 in the infected cells (**Extended Data Fig. 4D**), demonstrating that the virus replicates in T helper cells. We also found by plaque assay analysis that SARS-CoV-2-infected CD4^+^ T cells release infectious viral particles, although much less efficiently than Vero cells (positive control) (**Fig. 1E and Extended Data Fig. 4E**).

**Figure 1.**
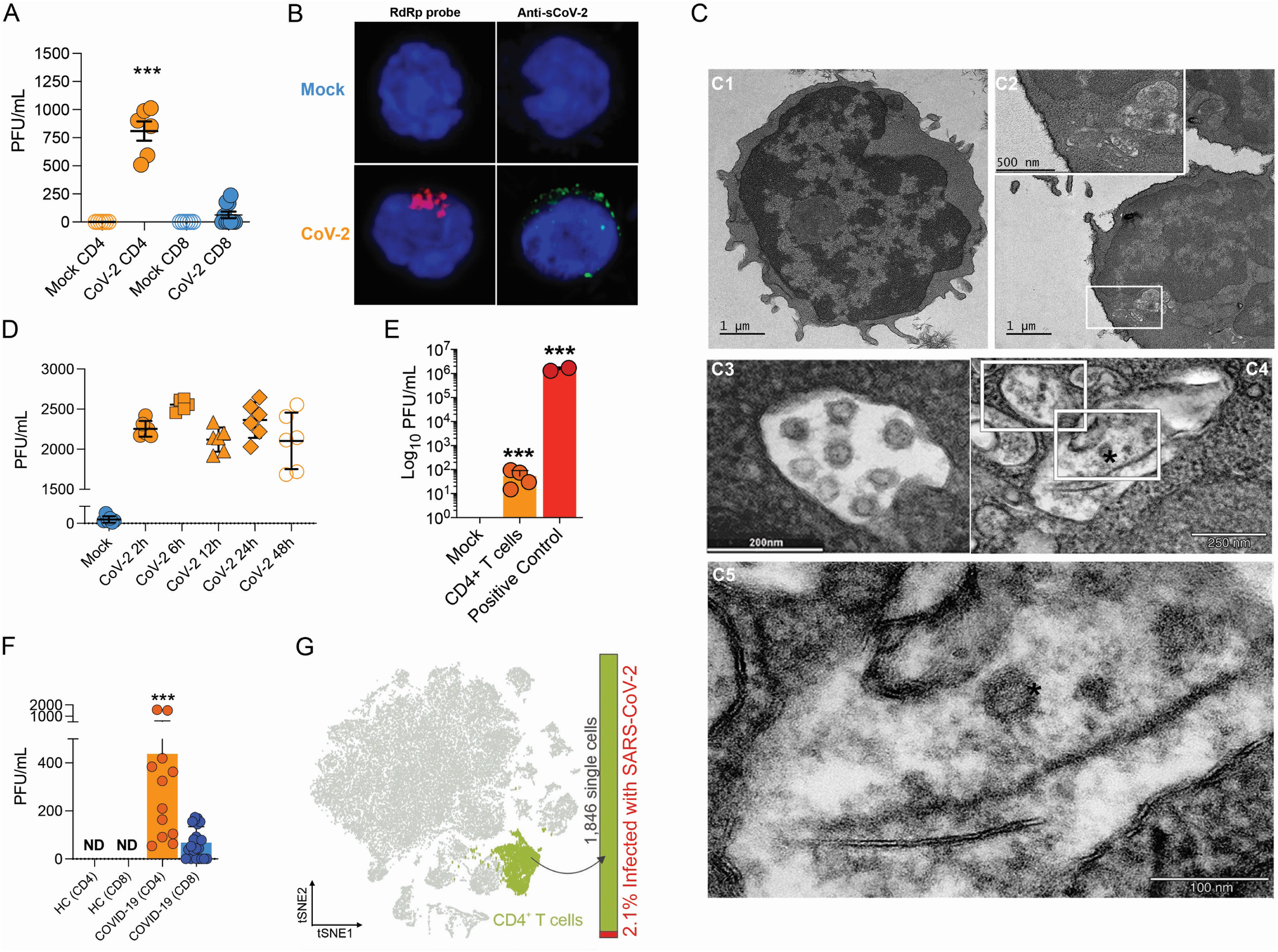
SARS-CoV-2 infects CD4^+^ T cells *in vitro* and *in vivo*. Peripheral blood CD3^+^CD4^+^ and CD3^+^CD8^+^ were infected with mock control or SARS-CoV-2 (CoV-2) (MOI 1) for 1 h under continuous agitation. **(A)** Viral load was assessed by RT-qPCR 24 h after infection. **(B)** Cells were washed, cultured for 24 h, fixed with PFA 4% and stained with RdRp probe for *in situ* hybridization or anti-sCoV-2 for immunofluorescence. Cells were analyzed by confocal microscopy. **(C) C1-C5** - Representative transmission electron microscopy (EM) micrographs showing viral particles (asterisks) inside lymphocytes 2 h after infection. **(D)** Viral load was determined by qPCR 2 h, 6 h, 12 h, 24 h and 48 h after infection. **(E)** Vero cells were incubated with the supernatant of mock control or CoV-2 infected CD4^+^ T cells under continuous agitation for 1 h. The viral load in Vero cells was measured after 72 h using plaque assay. PFU – plaque-forming unit. **(F)** Viral load was measured by RT-qPCR in peripheral blood CD4^+^ and CD8^+^ T cells from healthy controls (HC) and COVID-19 patients. **(G)** CoV-2 RNA detection in CD4^+^ T cells from bronchoalveolar lavage (BAL) single-cell sequencing data revealing the presence of CoV-2 RNA. Data represents mean ± SEM of at least two independent experiments performed in triplicate or duplicate (f). ***p < 0.001 compared to all. ND=not detected.

To investigate whether SARS-CoV-2 infects CD4^+^ T cells *in vivo*, we purified CD4^+^ and CD8^+^ T cells from peripheral blood cells of COVID-19 patients (**Table S1**). Similar to our *ex vivo* experiments, SARS-CoV-2 RNA was detected in CD4^+^ T cells from COVID-19 patients (**Fig. 1F**). Using publicly available single-cell RNA sequencing data^28^, we detected the presence of SARS-CoV-2 RNA in 2.1% of CD4^+^ T cells of the bronchoalveolar lavage (BAL) of patients with severe COVID-19 (**Fig. 1G and Extended Data Fig. 5**). Thus, our data demonstrate that SARS-CoV-2 infects CD4^+^ T cells.

Next, we sought to explore the role of the CD4 molecule in SARS-CoV-2 infection. Based on the putative interaction found using P-HIPSTer, we performed molecular docking analyses and predicted that sCoV-2 receptor binding domain (RBD) directly interacts with the N-terminal domain (NTD) of CD4 Ig-like V type (**Fig. 2A and Extended Data Fig. 6**). Molecular dynamics simulations with stepwise temperature increase were applied to challenge the kinetic stability of the docking model representatives (**Fig. 2B**). Two models remained stable after the third step of simulation at 353 Kelvin and represent likely candidates for the interaction between the CD4 NTD and sCoV-2 RBD (**Fig. 2B**). Additionally, we evaluated the dynamic behavior of closely related binding mode models present in the same cluster as these two models. We observed a strong structural convergence towards the putative model in one case, which indicates plausible and rather stable interaction between CD4 NTD and sCoV-2 RBD (**Extended Data Fig. 6**). The interaction region of RBD with CD4 is predicted to overlap with that of human ACE2 (**Fig. 2C and 2D**). The interaction between CD4 and sCoV-2 was confirmed by co-immunoprecipitation (**Fig. 3A**). Binding affinity isotherms, obtained by fluorescence anisotropy assay, confirmed the physical high affinity interaction between RBD and CD4 (Kd = 22 nM) and spike full length and CD4 (Kd = 27 nM). Considering the similar affinities, these results suggest the interaction interface between sCoV-2 and CD4 occurs at the RBD (**Fig. 3B-C**). Consistent with the hypothesis that CD4-sCoV-2 interaction is required for infection, we observed that pre-incubation with soluble CD4 (sCD4) completely blunted viral load in CD4^+^ T cells exposed to SARS-CoV-2 (**Fig. 3D**). Together, these data demonstrate that sCoV-2 binds to the CD4 molecule.

**Figure 2.**
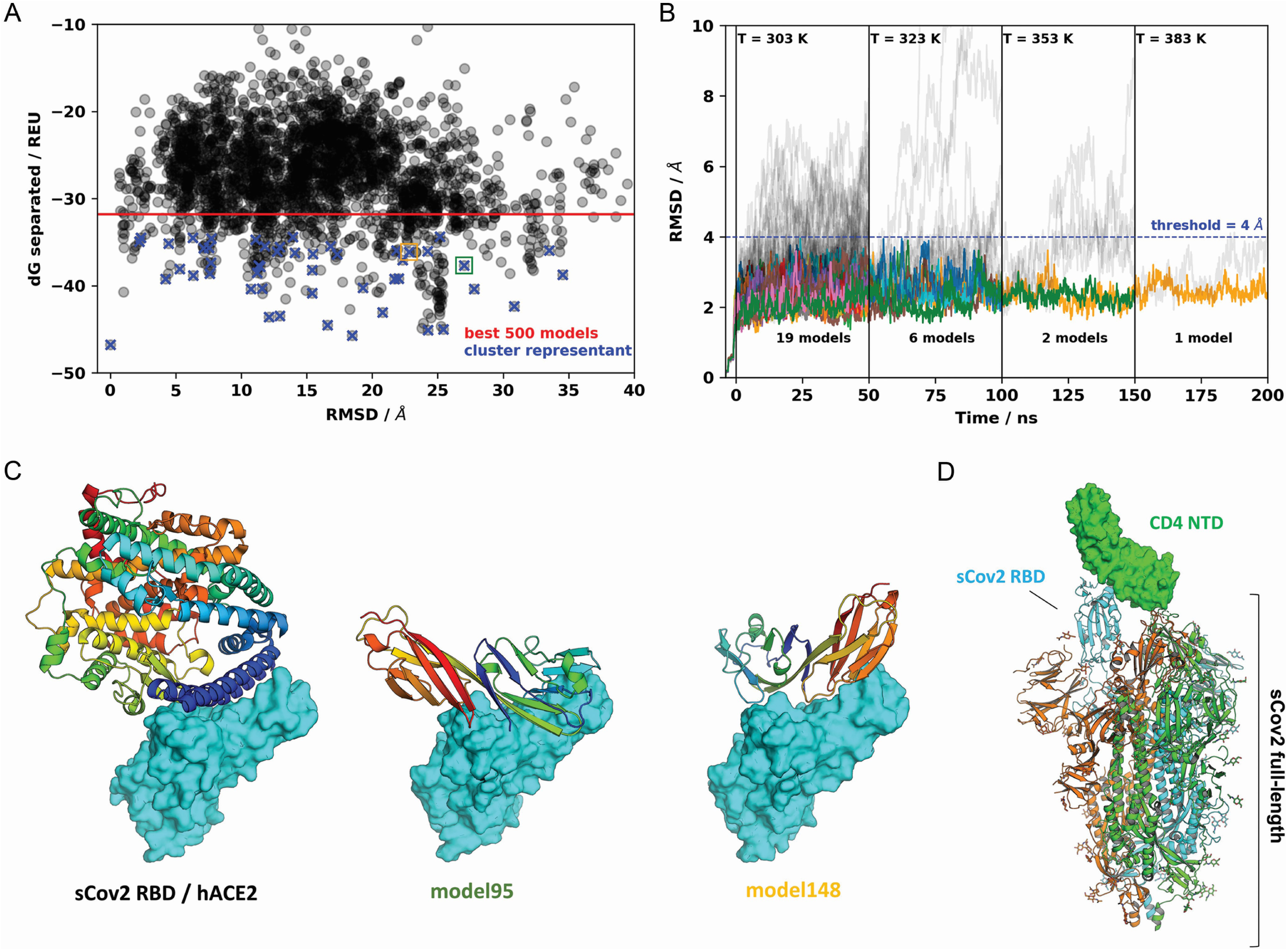
A model for sCoV-2 RBD and CD4 NTD interaction. **(A)** Interaction energy (dG separated) versus RMSD plot shows a high diversity of binding modes with similar energies. Among the 500 best models, 50 cluster best ranked representatives (shown as blue crosses) were selected for further evaluation with molecular dynamics simulations. **(B)**. RMSD to the initial docked complex as a function of molecular dynamics simulation time over 200 ns and four steps of temperature. At each temperature step, well behaved models are depicted as colored curves, while divergent candidates are shown as grey curves. Kinetically stable models making reasonable interactions remain close to their initial docked conformation. Only two models remain stable after the third step of 50 ns at 353K. For both models, resilient contacts throughout simulation are shown in Fig. S5. **(C)** Interaction models of ACE2 and sCoV-2 RBD. The two best candidates according to molecular dynamics simulation, model95 and model148, present distinct binding modes. For the first case, interaction occurs mainly in the N-terminal portion of CD4 NTD, while the latter have important contributions to the central part of this domain. **(D)** Full-length model of sCoV-2 and CD4 NTD interaction obtained by alignment of sCoV-2 RBD from model 148 to the sCoV-2 RBD open state from PDB 6vyb EM structure.

**Figure 3.**
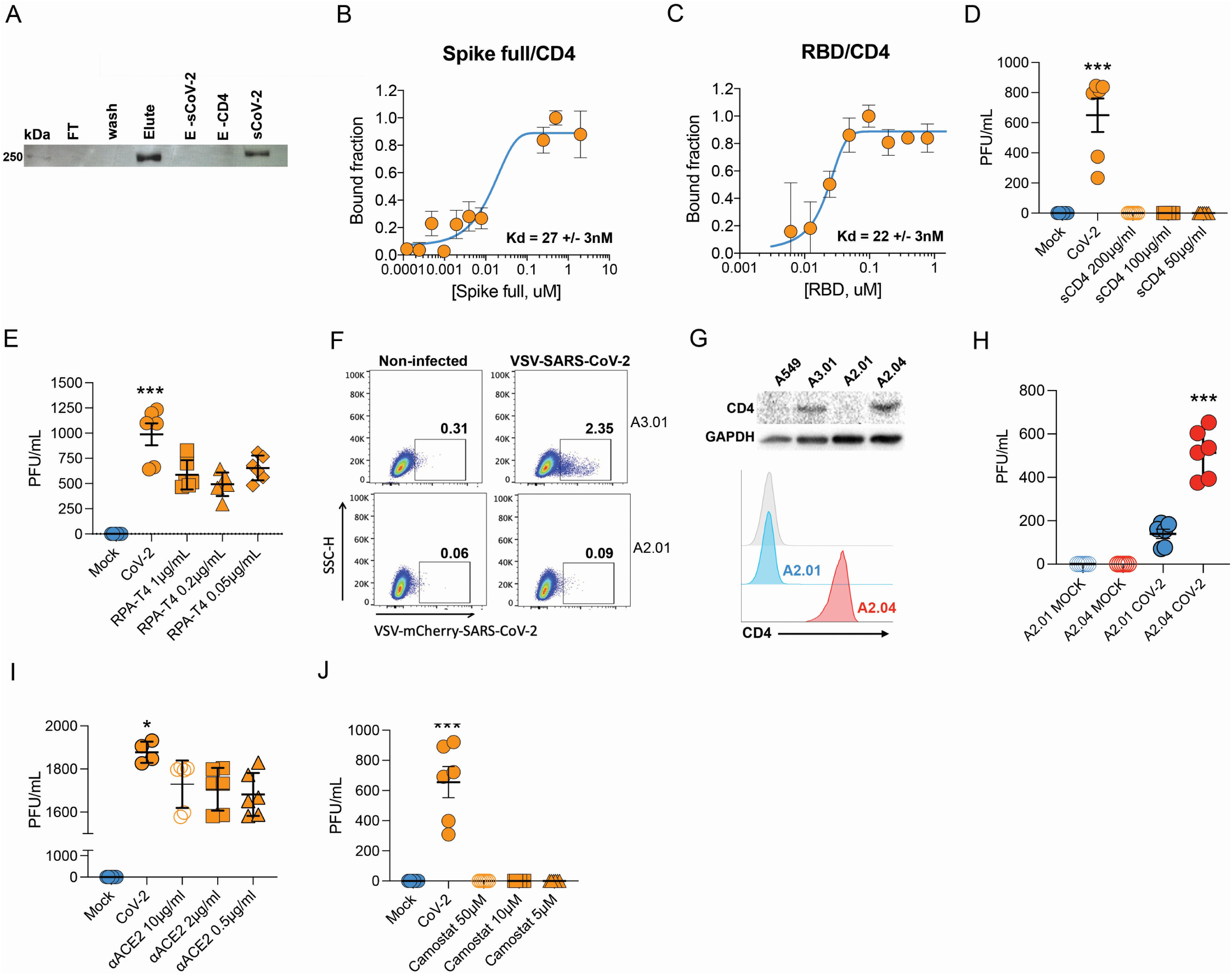
Infection of CD4^+^ T cells by SARS-CoV-2 is dependent on CD4 and ACE2 molecules. Recombinant sCoV-2 (with twin-strep-tag) and CD4 were co-incubated and immunoprecipitated with anti-CD4. Complex formation was determined by affinity blotting using streptavidin-HRP. Fluorescence anisotropy curves of Spike full length and **(B and C)** Spike RBD binding to CD4 labelled protein, presenting dissociation constants (Kd) of 27±3nM and 22±3nM, respectively. The CD4 was labeled with FITC by incubation with fluorescein isothiocyanate probe, at molar ratio 3FITC:1protein, 4°C for 3h. The probe excess was removed by a desalting column (HiTrap 5mL, GE) in a buffer containing 137 mM NaCl, 10 mM Na Phosphate, 2.7 mM KCl, pH of 7.4. To evaluate binding affinities, serial dilutions of Spike (RBD or full length) were performed, from 4.5uM to 100nM, over 20nM of labelled CD4. The measurements were taken using ClarioStar® plate reader (BMG, using polarization filters of 520 nm for emission and of 495 nm for excitation) and data analysis were performed using OriginPro 8.6 software. The Kds were obtained from data fitted to binding curves through the Hill model. (**D**) Cells were infected with mock control or SARS-CoV-2 (CoV-2) in the presence of vehicle or soluble CD4 (sCD4) in different concentrations (200, 100 or 50 μg/ml) for 1h under continuous agitation. Viral load was assessed by RT-qPCR 24 h after infection. **(E)** Primary human CD4^+^ T cells were incubated with IgG control, or monoclonal anti-CD4 (RPA-T4) antibody 18 h prior to infection with mock control or CoV-2. Viral load was determined by RT-qPCR 24 h after infection. **(F)** A2.01 and A3.01 lineages were cultivated with pseudotype (VSV-mCherry-CoV-2). Percentage of infected cells and flow cytometry analysis. **(G)** CD4 expression in A549, A3.01, A2.01 and A2.04 cells by western blotting (up) and flow cytometry analysis of CD4 expression in A2.01 and A2.04 (down). **(H)** Viral load of CoV2 in A2.01 and A2.04. **(I)** Peripheral blood CD4^+^ T cells were incubated with IgG control or anti-ACE2 (αACE2) polyclonal antibody 18 h prior to infection with mock control or CoV-2 for 1 h. Viral load was determined 24 h after infection. **(J)** CD4^+^ T cells were incubated with vehicle or camostat mesylate for 18 h before the infection with mock control or CoV-2 for 1h. Viral load was analyzed by RT-qPCR. Data represents mean ± SEM of at least two independent experiments performed in triplicate. ***p < 0.0001 compared to all; ^###^p< 0.001, ^#^ p < 0.05 compared to CoV-2

To gain further insight into the importance of CD4-sCoV-2 binding to SARS-CoV-2 infection, we purified CD4^+^ T cells and pre-incubated them with a CD4 monoclonal antibody (RPA-T4)^25^. We observed that CD4 inhibition reduced SARS-CoV-2 load (**Fig. 3E**). Moreover, we used human T cell lines that express CD4 (A3.01) or not (A2.01)^29^ (**Extended Data Fig. 7A**). Despite expressing higher levels of ACE2 and TMPRSS2 than primary CD4^+^ T cells (**Extended Data Fig. 2C, 7A and 7B**), the presence of CD4 was sufficient to increase viral load in A3.01 cells when compared to A2.01 (**Extended Data Fig. 7C**). These results were confirmed by using the VSV-mCherry-SARS-CoV-2 pseudotype model (**Fig. 3F**). Importantly, the introduction of CD4 in A2.01 increased viral load (**Fig. 3G and 3H**).

Our immunoprecipitation experiments indicated no physical interaction between CD4 and ACE2 (**data not shown**). Since CD4^+^ T cells have very low *ACE2* expression, we tested whether CD4 alone was sufficient to allow SARS-CoV-2 entry. Inhibition of ACE2 using polyclonal antibody (**Fig. 3I**) diminished SARS-CoV-2 entry in CD4^+^ T cells. Moreover, the inhibition of TMPRSS2 with camostat mesylate reduced SARS-CoV-2 load (**Fig. 3H**). Altogether, these data demonstrate that ACE2, TMPRSS2 and CD4 are all required to allow the infection of CD4^+^ T cells by SARS-CoV-2.

To assess the consequences of SARS-CoV-2 infecting CD4^+^ T cells, we performed mass spectrometry-based shotgun proteomics in CD4^+^ T cells exposed to SARS-CoV-2 *ex vivo*. We found that SARS-CoV-2 infection alters multiple housekeeping pathways associated with the immune system, infectious diseases, cell cycle and cellular metabolism (**Fig. 4A, 4B, Extended Data Fig. 8 and 9**). SARS-CoV-2 exposure elicits alterations associated with “cellular responses to stress”, which include changes in proteins involved in translation, mitochondrial metabolism, cytoskeleton remodeling, cellular senescence and apoptosis (**Fig. 4B and Table S2**). In agreement, *ex vivo* exposure of CD4^+^ T cells with SARS-CoV-2 resulted in 10% reduction of cell viability 24h after infection with a low MOI (0.1) (**Extended Data Fig. 10F**).

**Figure 4.**
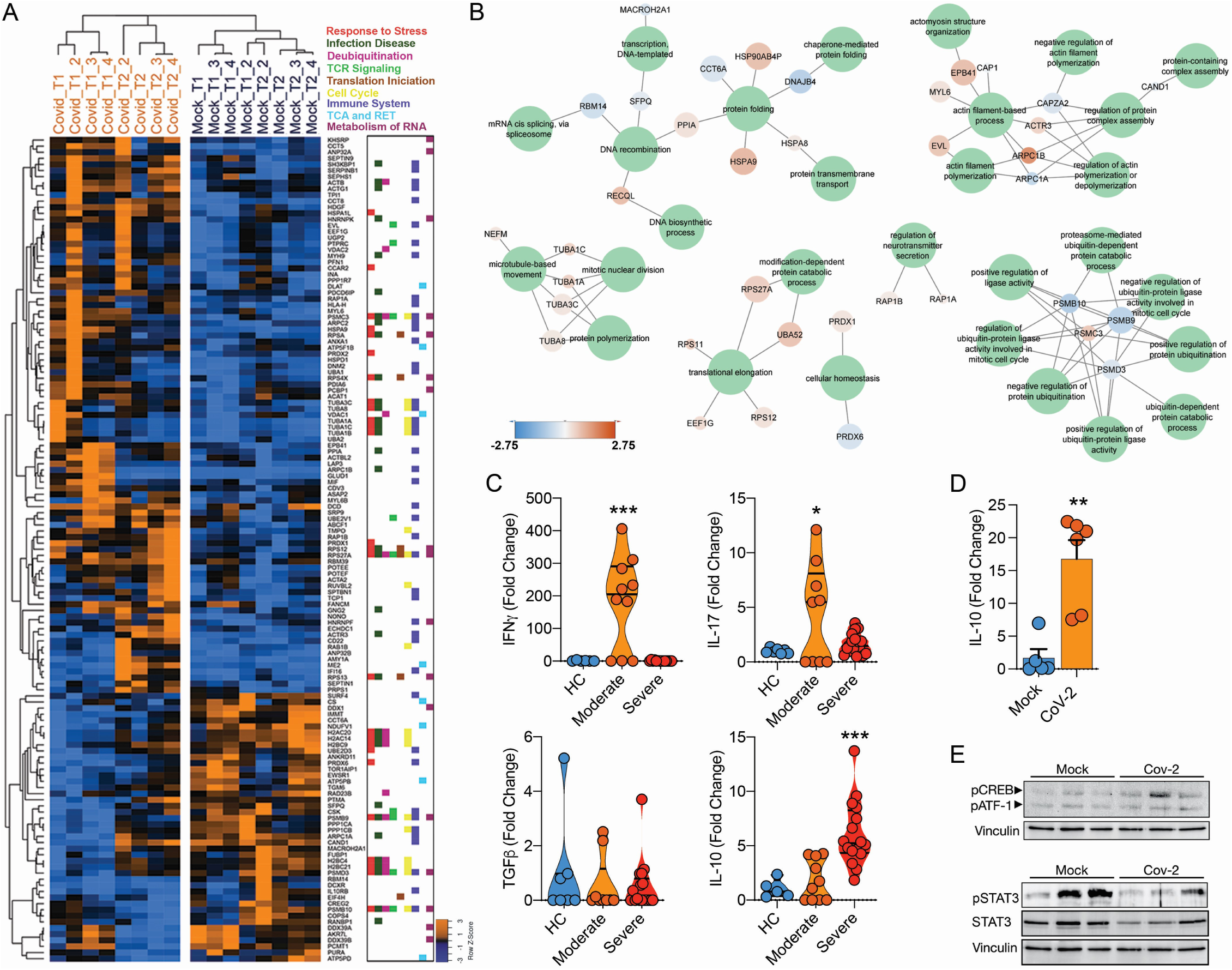
Infection of CD4^+^ T cells by SARS-CoV-2 alters cell function and triggers IL-10 production. **(A)** Heatmap of differentially expressed proteins and their associated biological processes. **(B)** Network of differentially expressed proteins and their associated biochemical pathways. **(C)** Relative expression of IFNγ, IL-17A, TGFβ-1, and IL-10 in peripheral blood CD3^+^CD4^+^ cells from COVID-19 patients (moderate or severe) and healthy control (HC). Data represents mean ± SEM. Each dot represents a patient sample. **(D)** Relative expression of IL-10 in primary CD4^+^ T cells infected with SARS-CoV-2 **(E)** Representative immunoblotting of CREB-1^Ser133^ and STAT3 in Peripheral blood CD3^+^CD4^+^ infected with mock control or SARS-CoV-2.***p < 0.001, **p < 0.00, *p < 0.05 compared to all.

The expression and release of IL-10 has been widely associated with chronic viral infections and determines viral persistence^30^. Noteworthy, increased serum levels of IL-10 are associated with COVID-19 severity^31,32^. We found that IL-10 expression by CD4^+^ T cells was higher in BAL (**Extended Data Fig.10B**) and blood (**Fig. 4C**) of severe COVID-19 patients. These changes were at least in part cell autonomous, since purified CD4^+^ T cells exposed to SARS-CoV-2 also expressed higher levels of IL-10 (**Fig. 4D**). Due to the immunomodulatory role of IL-10, we measured the expression of key pro- and anti-inflammatory cytokines involved in the immune response elicited by CD4^+^ T cells. CD4^+^ T cells from severe COVID-19 patients had decreased expression of IFNγ and IL-17A in relation to cells from patients with the moderate form of the disease or healthy donors (**Fig. 4C**). These results show that SARS-CoV-2 induces IL-10 expression in CD4^+^ T cells, which is associated with suppression of genes that encode key pro-inflammatory cytokines, such as IFNγ and IL-17A, and correlates with disease severity.

The activation of the transcription factor CREB-1 via Ser^133^ phosphorylation induces IL-10 expression^33^. Consistent with IL-10 upregulation, CREB-1 phosphorylation at Ser^133^ was increased in SARS-CoV-2-infected CD4^+^ T cells (**Fig. 4E**). Thus, SARS-CoV-2 infection triggers a signaling cascade that culminates in upregulation of IL-10 in CD4^+^ T cells. Altogether, our data demonstrate that SARS-CoV-2 infects CD4^+^ T cells, impairs cell function, leads to increased IL-10 expression and compromises cell viability, which in turn dampens immunity against the virus and contributes to disease severity.

Impaired innate and adaptive immunity is a hallmark of COVID-19, particularly in patients that progress to the critical stages of the disease^34,35^. Here we propose that the alterations in immune responses associated with severe COVID-19 are at least partially triggered by infection of CD4^+^ T helper cells by SARS-CoV-2 and consequent dysregulation of immune function. T helper cells are infected by SARS-CoV-2 using a mechanism that involves binding of sCoV-2 to CD4 and entry mediated by the canonical ACE2/TMPRSS2 pathway. We propose a model where CD4 stabilizes SARS-CoV-2 on the cell membrane until the virus encounters ACE2 to enter the cell. Moreover, Cecon and colleagues showed that CD4 co-expression with ACE2 in HEK293 cells decreases the affinity and the maximal binding between the sCoV-2 RBD and ACE2^20^, which is in line with our molecular dynamics results and the binding assays (**Fig. 3**) suggesting an intricate mechanism of modulation of SARS-CoV-2 infection in CD4^+^ T cells (**Fig. 2C, 2D, Extended Data Fig. 6**). Taken together, these data indicate that in CD4-expressing cells, direct binding of sCoV-2 to the CD4 molecule represents a potential mechanism to favor viral entry.

Once in CD4^+^ T cells, SARS-CoV-2 leads to protein expression changes consistent with alterations in pathways related to stress response, apoptosis and cell cycle regulation which, if sustained, culminate in cell dysfunction and may lead to cell death. SARS-CoV-2 also results in phosphorylation of CREB-1 transcription factor and induction of its target gene IL-10 in a cell autonomous manner.

IL-10 is a powerful anti-inflammatory cytokine and has been previously associated with viral persistence^30^. Serum levels of IL-10 increase during the early stages of the disease – when viral load reaches its peak – and may predict COVID-19 outcome^31,32^. This increase occurs only in patients with the severe form of COVID-19^32^. In contrast, we found IFNγ and IL-17A to be upregulated in CD4^+^ T cells of patients with moderate illness, indicating a protective role for these cytokines. However, in patients with the severe illness, the expression of IFNγ and IL-17A in CD4^+^ T cells is dampened. IL-10 is a known suppressor of Th1 and Th17 responses and it is likely to contribute to the changes in IFNγ and IL-17A. These features will ultimately reflect in the quality of the immune response, which in combination with T cell death and consequent lymphopenia, may result in transient/acute immunodeficiency and impair adaptive immunity in severe COVID-19 patients^6–8^.

How long these alterations in T cell function persist *in vivo* and whether they have long-lasting impacts on adaptive immunity remains to be determined. Hence, avoiding T cell infection by blocking sCoV-2-CD4 interaction and boosting T cell resistance against SARS-CoV-2 might represent complementary therapeutic approaches to preserve immune response integrity and prevent patients from progressing to the severe stages of COVID-19.

## Supporting information

Original Gel

## Data Availability

All data are storage in the university databank and will be provided under reasonable request.

## Author Contributions

A.S.F., N.S.B., G.G.D., V.O.B., L.B.M., T.L.K., J.V.V-LS, R.G.L., P.B.R. and A.B.S.P.G. performed most of the experimental work. B.G.C., A.B.P., L.D.C., M.C. and R.E.M. performed electron microscopy. G.S.P., N.S.W. and J.A.G. performed co-immunoprecipitation assay and immunoblotting. A.C.M.F and M.M.G.D performed fluorescence anisotropy assays. L.N.S., L.I.B. and H.M-S. performed *in situ* hybridization and immunofluorescence analysis. V.C.S., E.B.O. and V.M.F provided resources. V.C.C., L.C.S-C., F.C. and D.M-S. performed proteomic analyses. D.M., R.F.C., I.C. and H.I.N. performed bioinformatic analyses. A.J.R.F., A.H.S.D., and M.S. performed docking and molecular modeling analyses. A.P., E.M., R.G.U., A.F.B., T.A.N., L.C.R., M.V.A., M.L.M., H.R.C., FBP, A.C.S., R.S.C. and L.A.V. selected and recruited COVID-19 patients and healthy donors. G.F.S, S.P.M., J.F., N.S.B., M.R.A., D.A.T.T., P.L.P., M.C.M., K.B-S., C.L.S., F.G. and J.L.P.M. conducted the experiments with SARS-CoV-2 in the BSL-3 facility. F.P.V., L.A.T., L-M. B., S.S., M.C.P., S.P.J.W., N.S.B., N.M.A. and A.M.M. generate and performed experiments with pseudotype and T cell lines. A.S.F., M.A.M., P.M.M-V., D.M-S., M.S., L.M.B.S., M.A.R.V., A.S.V., A.D., L.A.V., R.E.M., J.L.P.M., H.M-S., H.I.N., T.M.C. and L.L.P.S. directed parts of the study and provided reagents. A.S.F. and M.A.M. coordinated the study and wrote the manuscript with inputs from all the co-authors.

## Acknowledgment

The authors acknowledge the technical support of Elzira E. Saviani, Paulo A. Baldasso and Mariana Ozello Baratti. The authors would like to thank Dr. Leda Castilho (UFRJ), for providing sCoV-2 recombinant protein and Dr. Hernandes F. de Carvalho (UNICAMP), for providing valuable reagents for ICC. We thank the National Institute of Science and Technology of Photonics Applied to Cell Biology (INFABIC) for aid with confocal microscopy. We thank Antonio Borges, Fabiano Montoro and LNNano/ CNPEM for the use of the electron microscopy facility (TEM-C2-26912, TEM-C2-26935). We acknowledge the Spectroscopy and Calorimetry Laboratory of the Brazilian Biosciences National Laboratory (LNBio), CNPEM, Campinas, Brazil, for the support in fluorescence anisotropy assays.

## Methods

### Subjects

The samples from patients diagnosed with COVID-19 were obtained at the Clinical Hospital of the University of Campinas (SP-Brazil). Both COVID-19 patients and healthy subjects included in this work signed a term of consent approved by the University of Campinas Committee for Ethical Research (CAAE: 32078620.4.0000.5404, CAEE: 30227920.9.0000.5404). Human blood samples from severe COVID-19 patients analyzed in this study were obtained from individuals admitted at the Clinics Hospital, University of Campinas and included in a clinical trial (UTN: U1111-1250-1843). Besides, *in vitro* experiments were performed with buffy coats from healthy blood donors provided by The hematology and hemotherapy Center of the University of Campinas (CAAE: 31622420.0.0000.5404).

### Diagnosis

Nasopharyngeal swabs or bronchoalveolar lavage fluids (BALF) were tested for SARS-CoV-2 by real time qPCR. The tests were performed in the Laboratory of High-Performance Molecular Diagnostic (LDMAD) or Laboratory of Clinical Pathology, all located at HC-UNICAMP. The samples were aliquoted and extracted manually using the MagMAX™ Viral/Pathogen Nucleic Acid Isolation Kit (Applied Biosystems). Some RNAs were automatically isolated by MagMAX™ Express-96 Magnetic Particle Processor (Applied Biosystems) following the latest protocol. The real time qPCRs were performed in duplicates using TaqMan™ Fast Virus 1-Step Master Mix (Applied Biosystems) for the detection of SarbecoV E-gene with specific primers (10 μM) and probe (5 μM). Primers sequences: Forward-ACAGGTACGTTAATAGTTAATAGCGT; Reverse-ATATTGCAGCAGTACGCACACA. Probe:6FAM-ACACTAGCCATCCTTACTGCGCTTCG-QSY. Patients were considered positives for COVID-19 whose curve was detected less Ct=38. The qPCRs were performed using Applied Biosystems™ 7500 Fast Real-time PCR system.

### Blood samples collection and lymphocyte separation

Each COVID-19 patient had heparin and plain blood tubes collected. Whole blood, serum and plasma samples were separated. Peripheral blood mononuclear cells (PBMCs) from COVID-19 patients and buffy coats were obtained through the Histopaque-1077 density gradient (Sigma-Aldrich). Samples were diluted in Hanks (1:1) and gently poured into 15 or 50 mL conical tubes containing 3 or 10 mL of Histopaque, respectively. Then, the samples were centrifuged at 1400 rpm for 30 minutes at 4°C without acceleration and brake. After PBMCs layer was collected into a new tube, lymphocytes were sorted and incubated overnight with RPMI 1640 (Gibco) containing 10% Fetal Bovine Serum (FBS) and 1% Penicillin-Streptomycin (P/S) incubated at 37°C with 5% CO2 atmosphere.

### A2.01, A2.04 and A3.01 cell Lineages

The lineages were obtained from NIH AIDS Reagent Program (Maryland, USA) and maintained in RPMI 1640 (Gibco) containing 10% Fetal Bovine Serum (FBS) and 1% Penicillin-Streptomycin (P/S), incubated at 37°C with 5% CO2 atmosphere. A3.01 is a lymphoblastic leukemia cell line that expresses a large amount of CD4 molecules, while A2.01 does not show the expression of CD4. A2.01 cells were transfected to express CD4 with green fluorescent protein (GFP) and to facilitate the understanding of manuscript we choose to call it A2.04.

### Virus

HIAE-02 SARS-CoV-2/SP02/human/2020/BRA (GenBank accession number MT126808.1) was isolated from the second confirmed case in Brazil and kindly donated by Professor Dr. Edson Luiz Durigon (University of São Paulo, Brazil). SARS-CoV 2 virus stocks were propagated in vero cell line (CCL-81, ATCC) and supernatant was harvested at 2 dpi. The viral titers were determined by plaque assays on Vero cells. Vero cells were propagated in Dulbecco’s Modified Eagle’s Medium (DMEM) supplemented with 10% FBS, 1% P/S, and maintained at 37°C in a 5% CO^2^ atmosphere.

### Flow cytometry

CD4^+^ T cells were sorted with FACSJazz and FACS Melody (BD Biosciences) as CD4^+^CD3^+^ and, in some experiments, CD8^-^CD3^+^. CD8+ T cells were sorted as CD8^+^CD3^+^ or CD4^+^CD3^+^ (See list of antibodies in supplementary Table 3). We performed flow cytometry with a lineage marker, Lin CD3, CD14, CD16, CD19, CD20 e CD56 to avoid contaminations. Sorted cells were incubated overnight in RPMI 1640 media containing 10% FBS and 1% P/S at 37°C with 5% CO2 atmosphere for posterior infection. All flow cytometry analyses were performed in FACS Symphony A5 (BD Biosciences). All antibodies used in the present study were sodium azide free.

### DNA probe biotinylated synthesis

Probe synthesis is based on two PCRs steps both using the same primers (primer sense: AACACGCAAATTAATGCCTGTCTG and primer antisense: GTAACAGCATCAGGTGAAGAAACA) for the RdRp gene. The first ‘amplifying’ PCR mix contains 1x buffer 5x, 1.5 mM of MgCl2, 0.2 mM of dNTP, 0.1 μM of each primer, 1 μL o cDNA (1:10), 1.5 U Taq polymerase (GoTaq® DNA Polymerase, Promega) in a final volume of 50 μL, submitted to 35 cycles of PCR including denaturation at 95ºC for 30 sec, annealing at 55ºC for 30 sec, 72ºC for 30 sec, and extension at 72ºC for 5 min. PCR samples were analyzed by electrophoresis on an agarose gel 1% in order to verify a final amplicon of 300 pb. The second labeling PCR contains slight modifications of the first, one using 0.2 mM of d (C, G, A) TP and 0.2 μM of 16-dUTP instead of the regular dNTP, and 1 μL of the previous PCR as a template (*9*).

### Plaque assays

Vero cells were grown in 24-well plates up to 80% confluency. Supernatant samples from 24h-infected lymphocytes were added and incubated at 37°C with 5% CO^2^ for an hour for virus adsorption. Samples were replaced with a semi-solid overlay medium (1% w/v Carboxy Methyl Cellulose in complete DMEM) and incubated at 37°C with 5% of CO^2^ for 3-4 days. Plates were fixed in 10% w/v PFA, stained in 1% w/v methylene Blue and results were expressed as viral plaque-forming units per milliliter of sample (PFU/mL).

### Co-immunoprecipitation

Co-immunoprecipitation was performed using Pierce protein A/G magnetics beads (ThermoFisher Scientific cat. no. 88803) immobilized with anti-CD4 antibody (Rhea Biotech cat. num. IM0566). Briefly, 100 μg of magnetic bead was washed in PBS containing 0.05% Tween-20 and then incubated with 10 μg anti-CD4 antibody for 2 hours at 4°C with mixing. For complex formation, we co-incubated 60 pmoles of recombinant CD4 (Sino Biological cat. num. 10400-H08H) and 60 pmoles of Spike S1 Twin-Strep-Tag recombinant protein expressed and generously provided by Dr. Leda Castilho from Federal University of Rio de Janeiro (*1*). Incubation was performed in 300 μL of PBS and kept overnight at 4°C with gentle agitation. After incubation, the recombinant protein solution was transferred to a 1.5 mL tube with the washed immobilized beads and incubated for 1h at room temperature. The complex was precipitated and proteins that did not bind to the beads were subsequently removed during the washes. The protein was eluted from the bead using Laemmli buffer and heat (95° for 5 min). The sample was analyzed by affinity blot using a streptavidin-HRP detection (Biolegend cat. num. 405210). Full range Rainbow™ (cat. num. RPN800E) was used as the molecular weight marker.

### Target selection

The P-HIPSTER database (http://phipster.org last access: June/01/2020) was used to find potential SARS-CoV-2 spike-human interactions. P-HIPSTER discoveries are predicted using protein similarities of known interactions (*2*). Only interactions with a final LR score ≥100 were used in the analysis. According to the authors, this threshold has a validation rate of 76%. To evaluate if these proteins were found on the cellular membrane, four databases were used. The Human Protein Atlas (https://www.proteinatlas.org last access: June/01/2020) contains tissue and anatomical protein annotations. 1917 proteins classified as “Cell Membrane” were retrieved. Panther GO is a gene set enrichment analysis tool (Mi H, PANTHER version 14: More genomes, a new PANTHER GO-slim and improvements in enrichment analysis tools). 1504 genes annotated as “integral components of plasma membrane” (GO:0005887) were retrieved.

EnrichR (https://amp.pharm.mssm.edu/Enrichr/ last access: June/01/2020) is a web based app that integrates various enrichment analysis tools (*3*) which library was used to retrieve JENSEN compartment annotations (https://compartments.jensenlab.org/ last access: June/01/2020) for “external side of plasma membrane” (212), “plasma membrane” (1148) and “cell surface” (715). To find common elements between the datasets and generate the Venn diagram, jvenn (http://jvenn.toulouse.inra.fr/app/ last access: 01/06/2020) was used (*4*).

### Gene set enrichment analysis

EnrichR was also used for enrichment analysis for SARS-CoV-2 spike protein predicted interactions. GO Molecular Function data was downloaded and enrichments with an adjusted p-value <0.05 were retrieved.

### Global expression analysis of ACE2, TMPRSS2 and CD4

To evaluate which tissues and cells express ACE2, TMPRSS2 and CD4, expression data (both protein and consensus mRNA expression, NX) from The Human Protein Atlas (https://www.proteinatlas.org, last access: July/07/2020) (*5*) and microarray expression from BioGPS (http://biogps.org/#goto=welcome, last access: July/072020) (*6*) were downloaded and plotted. In the Human Protein Atlas data, Only NX expression level ≥ 1 and protein detection ≥ “low” of at least one of the three genes was plotted. In the BioGPS data, this threshold was ≥10 a.u. (arbitrary units).

### sCoV-2-ACE2 interacting peptide (βACE2) design and synthesis

A peptide representing the interacting region of SARS-CoV-2 Spike protein with ACE2 and present at the recognition binding domain (RBD) was designed based on the tri-dimensional structure recently described (*7*). Based on the structure, we selected an unstructured and continuous segment of 16 amino acid residues (THR487-GLY502), which contained most of the interaction points to ACE2. We added to the segment a substitution of CYS488 to SER, which intended to avoid random disulfide bonds formation and structural alterations, keeping the same hydrophilicity in the interacting region. An amidated CYS residue was added at the C-terminal of the peptide in order to allow simple and specific conjugation with accessories detection molecules. The complete 17 amino acid residues (_487_NCYFPLQSYGFQPTNG_502_C), was synthesized using standard FMOC solid phase peptide synthesis chemistry as previously described (*8*) at a 100 μM scale, using RINK-amide resin 0.7mmol/g (Advanced chemteck CAT SA5130). All FMOC-aas (advanced chemteck) were used with 2.5 excess. FMOC-aa coupling reaction assisted by 6 cycles of 2 minutes in a home-made microwave device. At the end of coupling reactions, the peptide was cleaved from the resin using for 2 hours in a solution of 88% trifluoroacetic acid solution, 4% water, 4% triisopropylsilane, anisole 2 % + 30 mg DTT. Cleaved peptide was precipitated with ethylbutylether and then purified on C_18_ seppack solid phase extraction cartridges (10mg, Sigma-Aldrich) using water:acetonitrile solvent system.

### Proteomic Analysis

Both of the T-Cells Covid infected and Mock were resuspended in in RIPA buffer (150 mM NaCl, 1 mM EDTA, Tris-HCL 100mM, 1% triton-x final volume) with freshly added protease and phosphatase inhibitors (Protease Inhibitor Cocktail, SIGMA). After 3 cycles of 30 seconds of ultra-sonication for mechanical cell lysis, the protein amount was quantified by the Pierce BCA protein assay kit (Thermo Scientific).

Aiming to obtain a higher yield we performed the FASP protocol for subsequent analyses (Distler U., and Tenzer S., 2016). The FASP protocol is a method that allows to concentrate the proteins and clean up the samples through washing steps in a microcolumn tip with a 10kDa MW cut off, and to perform the tryptic digestion in this column. Ten micrograms of protein amount were used to carry out the FASP protocol, where the samples were reduced, alkylated and later digested using trypsin. An amount of 100 fmol/μl of digestion of Enolase from Saccharomyces cerevisiae was added to each sample as internal standard, then separation of tryptic peptides was performed on an ACQUITY MClass System (Waters Corporation). 1 μg of each digested samples has been loaded onto a Symmetry C18 5μm, 180μm × 20mm precolumn (Waters Corp.) and subsequently separated by a 120 min reversed phase gradient at 300 nL/min (linear gradient, 2–85% CH3CN over 90 min) using a HSS T3 C18 1.8 μm, 75 μm × 150 mm nanoscale LC column (Waters Corp.) maintained at 40 °C. After peptides separation, the ionized peptides were acquired by a Synapt G2-Si (Waters corp.). Differential protein expression was evaluated with a data-independent acquisition (DIA) of shotgun proteomics analysis by Expression configuration mode using the Ion Mobility cell (HDMSe). All spectra have been acquired in Ion Mobility Mode by applying a wave velocity for the ion separation of 800m/s and a transfer wave velocity of 175m/s. The mass spectrometer operated in “Expression Mode”, switching between low (4 eV) and high (25–60 eV) collision energies on the gas cell, using a scan time of 0.5 s per function over 50–2000 m/z. The processing through low and elevated energy, added to the data of the reference lock mass ([Glu1]-Fibrinopeptide B Standard, Waters Corp.) provides a time-aligned inventory of accurate mass-retention time components for both low and elevated-energy (EMRT, exact mass retention time).

Each sample was run in four technical replicates. Continuum LC-MS data from three replicate experiments for each sample was processed for qualitative and quantitative analysis using the software Progenesis QC for Proteomics (PLGS, Waters Corp.). The qualitative identification of proteins was obtained by searching in the Homo sapiens database (UniProtKB/Swiss-Prot Protein reviewed 2020). The expression analysis was performed considering technical replicates available for each experimental condition following the hypothesis that each group is an independent variable. The protein identifications were based on the detection of more than two fragment ions per peptide, and more than two peptides measured per protein. The list of normalized proteins was screened according to the following criteria: protein identified in at least 70% of the runs from the same sample and only modulated proteins with a p< 0.05 were considered significant. Raw data are available in ProteomeXchange database under accession number PXD020967 (Username; Password: q3qYRlYQ).

### In situ Hybridization

Cells were infected for 24 hours with the SARS-CoV-2 virus (MOI of 1) or mock, as described before, washed with PBS, fixed with PFA 4% for 15 minutes at room temperature, and followed by two washes with PBS 1x pH 7,4 DEPC to further seed the cells on silanized glass slides kept on a pre-warmed surface until liquid is evaporated. The CD4+ cells were pelleted at 1500 rpm for 5 min when necessary. The slides were washed with PBS and pre-treated with 2% H2O2 in methanol for 30 minutes, to bleach auto- and avoid non-specific fluorescence, washed twice with PBST, treated with PK (10 μg/ mL) for two minutes, followed by PBST-Glycin (2 mg/ mL) for 10 minutes, and a second fixation step with PFA 4% for 10 minutes and a couple of more washes with PBST at room temperature. Each sample was pre-hybridized with hybridization solution (Hyb) (50% formamide, 10% dextran sulfate, 2xSSC pH 7 and 100 ug de DNA from salmon) without the probe for at least two hours at 37ºC in humidity box, and then incubated with Hyb containing the biotinylated probe (100 μL of the probe in 500 μL de Hyb) pre-denaturated (85ºC for 10 min) in a humidity box overnight at 37ºC. The slides were then washed for 20 min at 37ºC with 50% Hyb in 2xSSC, 25% Hyb in 2xSSC, and then followed by 2 washes for 10 min each with 2xSSC, 0,2xSSC, and PBS. All samples were incubated with streptavidin fluorescent (Streptavidin, DyLight® 594 Conjugated, Thermo Scientific, #21842, 1:300) for two hours at room temperature in the humidity box protected from the light (*9, 10*). The slides were washed with PBST, incubated with DAPI (Santa Cruz Biotechnology - SC-3598) diluted 1:1000 in in PBST for 5 minutes at room temperature protected from the light, and mounted in an aqueous mounting solution for confocal imaging.

### Detection of SARS-CoV-2 antisense strand

cDNA was generated using SuperScript™ III First-Strand Synthesis SuperMix (ThermoFisher) as described previously (*12*). 60 ng of RNA were incubated at 65°C for 5 minutes with 1X annealing buffer and 250 nM of a primer complementary to the anti-sense CoV-2 strand containing a non-viral tag at the 5’ end. The sample was chilled at 4°C for 1 minute and incubated at 55°C for 50 minutes with SuperScript™ III/RNaseOUT™ and 1X master mix for reverse transcription (RT). The sample was heated to 85°C for enzyme inactivation. The antisense strand was amplified in a PCR using 100 nM of primers complementary to the non-viral tag and to the antisense CoV-2 strand. As a control for false-priming (*13*), we performed the RT without the tagged reverse primer and a PCR using both the RT and the PCR primers at 100 nM. The sense strand was also amplified as a control with the same method using non-tagged primers for the RT and PCR. The samples were loaded in a 4% agarose gel in TBE containing 1X SybrSafe (ThermoFisher).

### Immunoblotting

Cells were infected for 3 hours with the SARS-CoV-2 virus (MOI of 1) or mock, washed with PBS and the protein was extracted using lysis buffer [Tris HCl 100mM pH 7.5; EDTA 1mM; NaCl 150 mM; 1% Triton-100X; 1x protease and phosphatase inhibitors]. The protein quantification was performed using Pierce BCA protein assay Kit (Thermo Fisher Scientific). Next, 5 μg protein of each sample was placed Laemmli buffer and heat at 95ºC during 5 minutes. Protein content was separated by 10% SDS-PAGE and transferred to PVDF membrane (cat. num. 1620177-BioRad). After the transference, membranes were blocked with StartingBlock ™ (37539-Thermo Fisher Scientific) during 30 minutes, and next incubated overnight with anti-phospho-CREB (cat. num. 9198-CellSignaling, dilution 1:1000), p-STAT3 (cat. num 8059-Santa Cruz, dilution 1:250) or Total STAT3 (cat. num 4904-Cell Signaling dilution 1:1000). Membranes were washed and incubated during 1 hour with horseradish peroxidase-conjugated secondary antibodies Rabbit IgG HRP (cat. num. RPN4301-GE Healthcare, diluted 1:10000 in block buffer). Membranes were then washed, and incubated with Immobilon Western Chemiluminescent HRP Substrate (cat num. WBKLS0500-Millipore), and the images were acquired in ChemiDocTM Gel Imaging System (Biorad). The samples were normalized with anti-Vinculin antibody (cat num. ab91459-Abcam). The image analysis was performed using software Image J.

### Immunofluorescence

Cells were prepared onto silanized glass slides as previously described for in situ hybridization, followed by a washing step using PBST 0,1M pH 7,4. To avoid autofluorescence, the cells were treated with 2% H2O2 methanol for 30 minutes, washed with PBST, and treated with glycine 0,1M in PBST for 10 minutes at room temperature. The samples were then washed and treated with 1% Bovine Serum Albumin (BSA) solution in PBST for 30 minutes, to block nonspecific epitopes. Cells were incubated with SARS-CoV-2 Spike S1 Antibody (HC2001) (GenScript - A02038) diluted 1:100 in BSA 1% solution in PBST, and incubated overnight at 4ºC in a humid box. The slides were then washed and incubated with Anti-Human IgG Alexa 488 (ThermoFisher - A11013) diluted 1:500 in BSA 1% solution in PBST for two hours at room temperature in a humid box, protected from the light. The samples were washed again, incubated DAPI (Santa Cruz Biotechnology - SC-3598) diluted 1:1000 in BSA 1% solution in PBST for five minutes at room temperature protected from the light, and mounted in an aqueous mounting solution for confocal imaging.

Microscopy images were acquired with a Zeiss LSM880 with Airyscan on an Axio Observer 7 inverted microscope (Carl Zeiss AG, Germany) with a C Plan-Apochromat 63x/1.4 Oil DIC objective, 4x optical zoom. Prior to image analysis, raw .czi files were automatically processed into deconvoluted Airyscan images using Zen Black 2.3 software. For DAPI were acquired convencional confocal image using 405 nm laser line for excitation and pinhole set to 1 AU.

### Transmission electron microscopy

T Lymphocyte cell cultures were pelleted by centrifugation at 500g for 10 min. Cell culture supernatant was removed and cells were resuspended in fixative solution (2.5% glutaraldehyde in 1M sodium cacodylate containing 3mM calcium chloride) and kept overnight (16h) in 4°C. Cells were pelleted (1500g for 2min) and washed in cacodylate buffer with calcium chloride for a total five times. Post-fixation was performed with 1% reduced osmium tetroxide plus 0.8% potassium ferrocyanide in a cacodylate buffer with calcium chloride for 2h at 4°C covered from light. Pellet was recovered by centrifugation at 1500g at 4°C and washed three times in ddH_2_O for 2 min. Cells were resuspended in 4% low melting agarose (apx. 50°C) and centrifuged at 1500g for 10min at 30°C and kept 20 min in ice for solidification. Agarose pellets were trimmed in small blocks and stained for contrast enhancement with 2% aqueous uranyl acetate overnight at 4°C. Later, samples were washed in ddH_2_O and dehydrated in increasing concentrations of ethanol (20, 50, 70, 80, 90, 100% twice), 1:1 ethanol:acetone, and acetone (twice), 20 min each. Samples were embedded in Embed-812 (Electron Microscopy Science, USA, #14120) following manufacturer’s recommendations. Ultrathin sections (70nm) were stained mounted in 200 mesh copper grids and stained with 2% aqueous uranyl acetate and Reynold’s lead citrate. Finished grids were imaged in a JEOL JEM-1400 Transmission Electron Microscope (120kV accelerating voltage) and/or Helios Nanolab Dualbeam 660 Scanning Transmission Electron Microscope (30kV accelerating voltage) at Electron Microscopy Laboratory, Brazilian Nanotechnology National Laboratory-LNNano, CNPEM.

### RNA extraction cDNA synthesis

Total RNA was extracted from samples using TRI Reagent (Sigma) according to manufacturer’s instructions and quantified using NanoDrop 200 Spectrophotometer (Thermo Scientific). cDNA was synthesized using GoScript Reverse Transcriptase Kit (Promega) or High Capacity cDNA Reverse Transcription Kit (Applied Biosystems) following manufacturer’s instructions.

### qPCR

Real time PCR (qPCR) was performed using QuantiNova SYBR Green PCR Kit (Qiagen) with specific primers (HPRT - Forward: GACCAGTCAACAGGGGACAT, Reverse: AACACTTCGTGGGGTCCTTTTC; IL10 - Forward: GCCTAACATGCTTCGAGATC; Reverse: CTCATGGCTTTGTAGATGCC; TGFB1 - Forward: AAGTTGGCATGGTAGCCCTT, Reverse: GCCCTGGATACCAACTATTGC; IFNG Forward: TTTAATGCAGGTCATTCAGATGTA, Reverse: CACTTGGATGAGTTCATGTATTGC and IL17A Forward: TCCCACGAAATCCAGGATGC, Reverse: TGTTCAGGTTGACCATCACAGT. Expression levels of each gene were normalized to housekeeping gene HPRT. Gene expression fold change was calculated with the ΔΔCt method. Viral load was determined using specific SARS-CoV-2 N1 primers, as described (*11*) and using qPCRBIO Probe 1-Step Go Lo-Rox (PCR Biosystem) with specific SarbecoV E-gene primers and probe. For the preparation of a standard curve, serial dilutions of SARS-CoV-2 were used. qPCR was performed using BIO-RAD CFX384 Touch Real-Time PCR Detection System and ThermoFisher QuantStudio 3.

Relative expression of the viral envelope (E), nucleocapsid (N) and RNA-dependent RNA polymerase (RdRp) genes from i*n vitro* CD3^+^CD4^+^ cells were detected using GeneFinder™ COVID-19 Plus Real*Amp*Kit (OSANG Healthcare Co.). Expression level of each gene was normalized to internal control of kit and gene relative expression was calculated through to 2^-ΔCT^. For this kit, the qPCR was performed using Applied Biosystems™ 7500 Fast Real-time PCR system.

### Single-Cell mRNA-seq

We re-analyze the scRNAseq data from 6 samples of bronchoalveolar lavage fluid immune cells of severe COVID-19 patients (C145, C146, C148, C149 e C152) by downloading the respective single cell processed data matrices from GEO under the accession number GSE145926. The number of viral transcripts mapped to SARS-CoV-2 were previously integrated by their authors as an additional feature into data matrices called “NcoV” used to quantify the viral load of Sars-Cov-2. The data matrices were then imported to R version 3.6.3 and analyzed using Seurat v3.1 (Stuart et al, 2019).

A control quality filtering was applied to remove low quality cells considering a gene number between 200 and 6,000, UMI count > 1,000 and mitochondrial gene percentage < 0.1. After the filtering step, a total of 37,197 cells and 25722 features (including SARS-CoV-2 viral load) were left for downstream analysis. The filtered gene-barcode matrix of all samples was integrated with FindIntegrationAnchors and IntegrateData functions considering the first 50 dimensions of CCA. Then, the filtered data matrix was normalized using the ‘LogNormalize’ method in Seurat with a scale factor of 10000. The top 3,000 variable genes were then identified using the ‘vst’ method in Seurat with FindVariableFeatures function. Variables ‘nCount_RNA’ and ‘percent.mito’ were regressed out in the scaling step and PCA was performed using the top 3,000 variable genes with 100 dimensions. Then, UMAP and tSNE was performed on the top 40 principal components for visualizing the cells. Additionally, a clustering analysis was performed on the first 40 principal components using a resolution of 0.8. Then, differential gene expression analysis was performed with Wilcoxon Rank Sum test using FindAllMarkers function to obtain a list of significant gene markers for each cluster of cells. Significant gene markers were considered with adjusted P-value <0.05 and average Log Fold Change > 0.25 among cells. T cell clusters annotation was performed using an evidence-based score approach with scCATCH (Shao et al, 2020) based on differential gene markers list. Finally, CD4+ T cell subpopulations were then annotated considering a high differential expression of CD4 CCR7 and IL32 gene markers (**Extended Fig. 5**). Considering the methodology applied above, a total of 1846 single cells were identified as CD4+ T cells.

To measure infection of SARS-CoV-2 in CD4+ T cells, T cells were considered infected assuming a threshold with at least 10 percent of viral load expression. The applied threshold resulted in 39 single cells infected with SARS-CoV-2.

### Docking

The RosettaDock4.0 protein docking protocol (*12, 14*) was applied to model the interaction between CD4 N-terminal domain (NTD) (*15*) (PDB ID 1wio, residues 1-178) and sCov2 RBD (*16*) (PDB ID 6lzg, residues 333-527) as described previously. Briefly, the first step consisted of generating an ensemble of 100 conformers for each target. The two-stage automated Rosetta docking protocol simulates the physical encounter of the proteins and maximizes interactions, which may lead to binding. In the first stage (global docking), a rigid body translation-rotation of one of the proteins samples possible interaction modes using a low resolution, centroid-based representation of the sidechains. RosettaDock4.0 also mimics binding-induced conformational adaptations by assessing a pre-generated list of conformers.

This is an important feature for cases where backbone perturbation is inherent to the interface of interaction. Additionally, a score term named Motif Dock Score ranks low-resolution models according to their chance to generate promising high-resolution models, reducing the number of candidates to proceed to the next stage. In the second stage (local docking) the centroid mode is converted into full-atom representation, where small random rigid body perturbations take place, followed by sidechain relaxation that aims to optimize local interactions. A first set of models consisted in generating 10,000 low-resolution candidates using global docking with 3 Å translation and 8 degrees rotation perturbation parameters. The 2,000 highest-ranked models according to Motif Dock Score proceeded to the local docking stage using default parameters (0.1 Å translation and 3 degrees rotation perturbation parameters). 25 high-resolution candidates were generated for each low-resolution input model totalizing 50,000 models. A second set of models was generated by first randomizing CD4 NTD orientation previously to docking run. For this strategy, 40,000 low resolution and 300,000 high resolution models were generated similarly to the description above. As an alternative docking approach, monomeric chains were also docked using HADDOCK 2.4 server (*17*), which returned 40 model candidates spread into 13 clusters. 800 additional models were generated using these models as input to the high-resolution docking stage at RosettaDock4.0. Next, best ranked 2,000 models according to interface score (I_sc) were fully relaxed with Relax application (beta_genpot score function, Rosetta Package) and evaluated with InterfaceAnalyzer application (Rosetta package) to extract interface metrics. 500 top ranked models according to interface binding energy (dG_separated) were evaluated for pair-wise structural similarity with LovoAlign (*18*) and were clustered at 2.0 Å radius. Best ranked models of each of the first 50 clusters were selected for molecular dynamics simulation. This set of models is available at https://github.com/ajrferrari/CD4-RBD-interaction-models.

### Molecular Dynamics Simulations

Molecular dynamics (MD) simulations have been used to discriminate among protein complexes decoys by challenging their thermal stability (*19*). All-atom MD simulations were carried out with pmemd within AMBER20 software suite for 50 representative models of CD4 NTD sCov2 RBD interaction obtained as described above. Glycosylation at sCov2 RBD N343 residue (*20, 21*) was built using the Glycam web server. Each system was solvated in a periodic octahedral water box such that the initial structure was more than 24 Å of its closest image. Neutralizing ions (Na^+^ and Cl^-^) were added to a final concentration of 0.15 M (*22*). The ff14SB force field (*23*) was used for modeling proteins, the GLYCAM06 force field (*24*) for modeling the glycan, and water was described with the TIP3P model (*25*). Electrostatic interactions were evaluated using the Particle Mesh Ewald algorithm (*26*), nonbonded interactions were truncated at 9 Å, bonds involving hydrogen atoms were constrained at their equilibrium values, and a time step of 2 fs was used for the numerical integration of the equations of motion. Before production runs, all systems were equilibrated in five sequential steps: i) 2500 steps of steepest descent line minimization followed by 2500 of conjugate gradient minimization; ii) 1 ns simulation at 303 K in the NVT ensemble with harmonic restraints of 10 kcal mol^-1^ Å^-2^ to protein and glycan atoms; iii) 1 ns simulation at 303 K in the NPT ensemble with harmonic restraints of 10 kcal mol^-1^ Å^-2^ to protein and glycan atoms; iv) 1 ns simulation at 303 K in the NPT ensemble with harmonic restraints of 10 kcal mol^-1^ Å^-2^ to protein backbone atoms and; iv) 1 ns simulation at 303 K in the NPT ensemble without restraints. Subsequently, production runs for assessment of the kinetic stability of the proposed structural models (*19*) were performed in the NPT ensemble at four sequential steps: i) 50 ns simulation at 303 K; ii) 50 ns simulation at 323 K; iii) 50 ns simulation at 353 K; and iv) 50 ns simulation at 383 K. At each production step, models were evaluated according to their RMSD deviation to the initial model such that models with RMSD greater than 4 Å did not proceed to the next step. Additionally, for the data contained in Figure Sx, triplicate production runs were performed for 100 ns at 303K in the NPT ensemble. All trajectory analysis was performed with CPPTRAJ (*27*). Model structural visualization and image rendering were performed either in PyMOL (*The PyMOL Molecular Graphics System, Version 2*.*0 Schrödinger, LLC*, n.d.) or VMD (*28*).

### Fluorescence anisotropy

Human CD4 (Sino Biological / 10400-H08H), ACE2 (Genscript), and purified TR (human thyroid receptor – negative control) proteins were labeled with FITC by incubation with fluorescein isothiocyanate probe, at molar ratio 3FITC:1protein, 4°C for 3h. The probe excess was removed by a desalting column (HiTrap 5mL, GE) in a buffer containing 137 mM NaCl, 10 mM Na Phosphate, 2.7 mM KCl, pH of 7.4.

To evaluate the affinities between RBD and CD4, TR or ACE2, serial dilutions of RBD (0.01 to 10 μM) were performed. To evaluate the affinity between Spike full length (LECC/UFRJ LM220720) and CD4, serial dilutions of Spike (0.01 to 4,5 μM) were performed. The anisotropy curves were assembled in three replicates, in black 384-well plates (Greiner). After serial dilution set up, each point was incubated with 20 nM of the labelled proteins (CD4, ACE2, or TR) for at least 2h at 4°C. For each fluorescence curve, the mixtures were submitted to fluorescence anisotropy measurements using ClarioStar® plate reader (BMG, using polarization filters of 520 nm for emission and of 495 nm for excitation). Data analysis was performed using OriginPro 8.6 software and Kd was obtained from data fitted to binding curves through Hill model.

---

**Extended Data Fig. 1.**
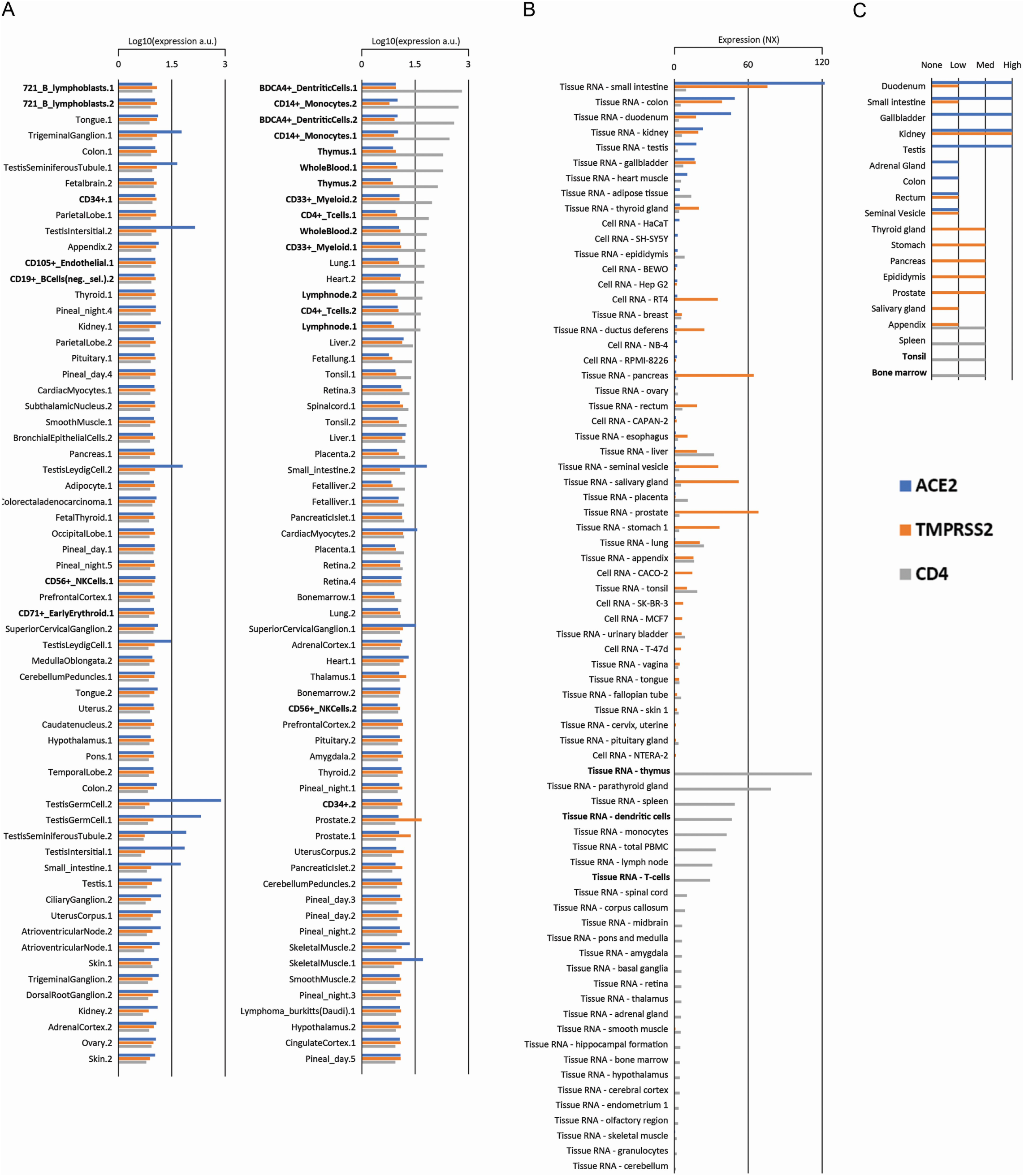
**(A)** Expression of ACE2, TMPRSS2 and CD4 in multiple tissues and cell types according to BioGPS database and The Human Protein Atlas. **(B)** NX expression denotes consensus mRNA expression between multiple databases. **(C)** Protein detection through immunohistochemistry data was plotted based on The Human Protein Atlas annotations (Quantity classifications are subjective). Top 25 cells/tissues with the highest expression of CD4 are marked.

**Extended Data Fig. 2.**
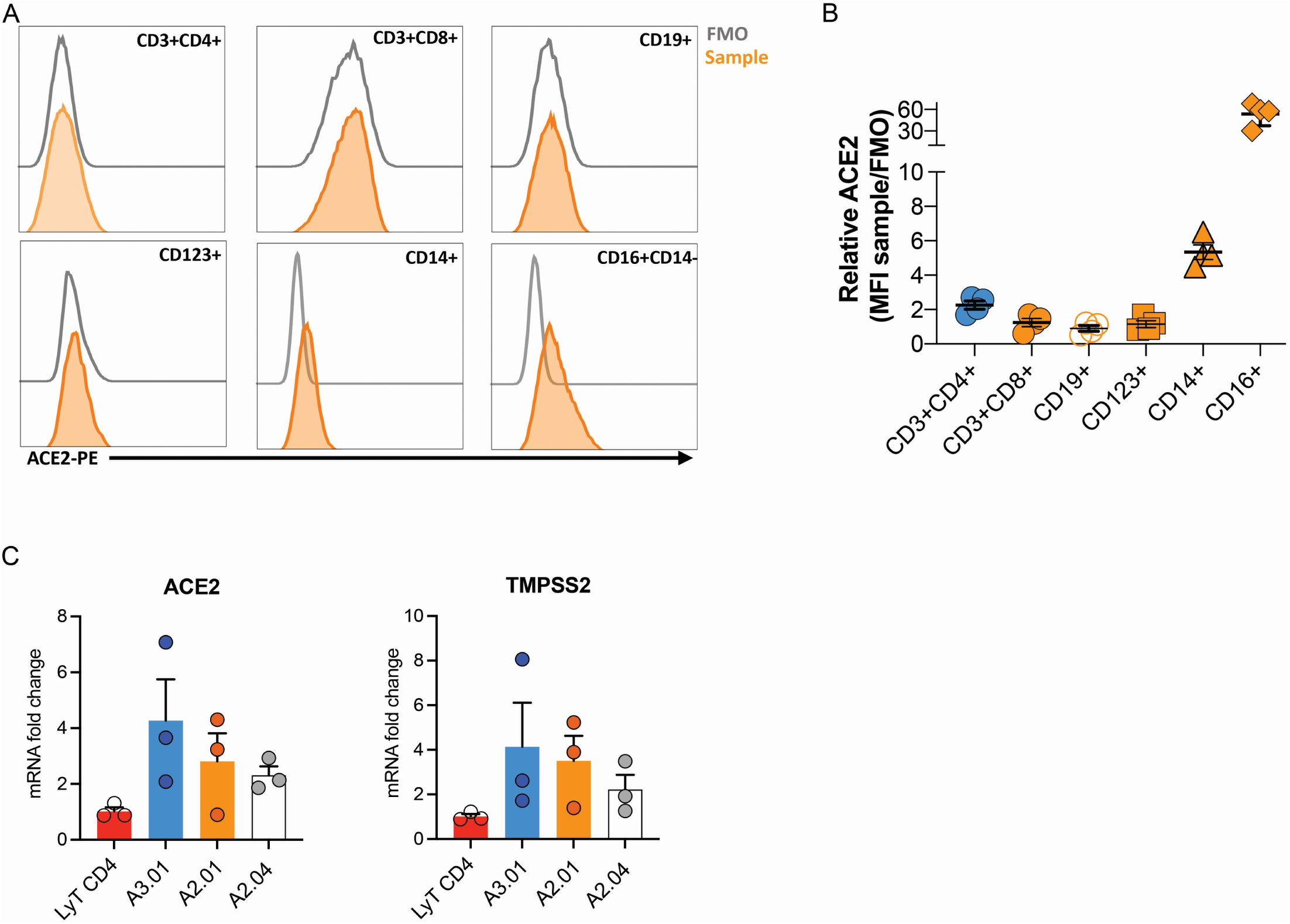
**(A-B)**. Flow cytometry analysis of ACE2 in peripheral blood cells (FMO: gray and anti-ACE2: orange): CD3+CD4+, CD3+CD8+, CD19+, CD123+, CD14+ and CD16+CD14-**(C)** mRNA expression of ACE2 and TMPRSS2 in T lymphocytes, A3.01, A2.01 and A2.04 cells.

**Extended Data Fig. 3.**
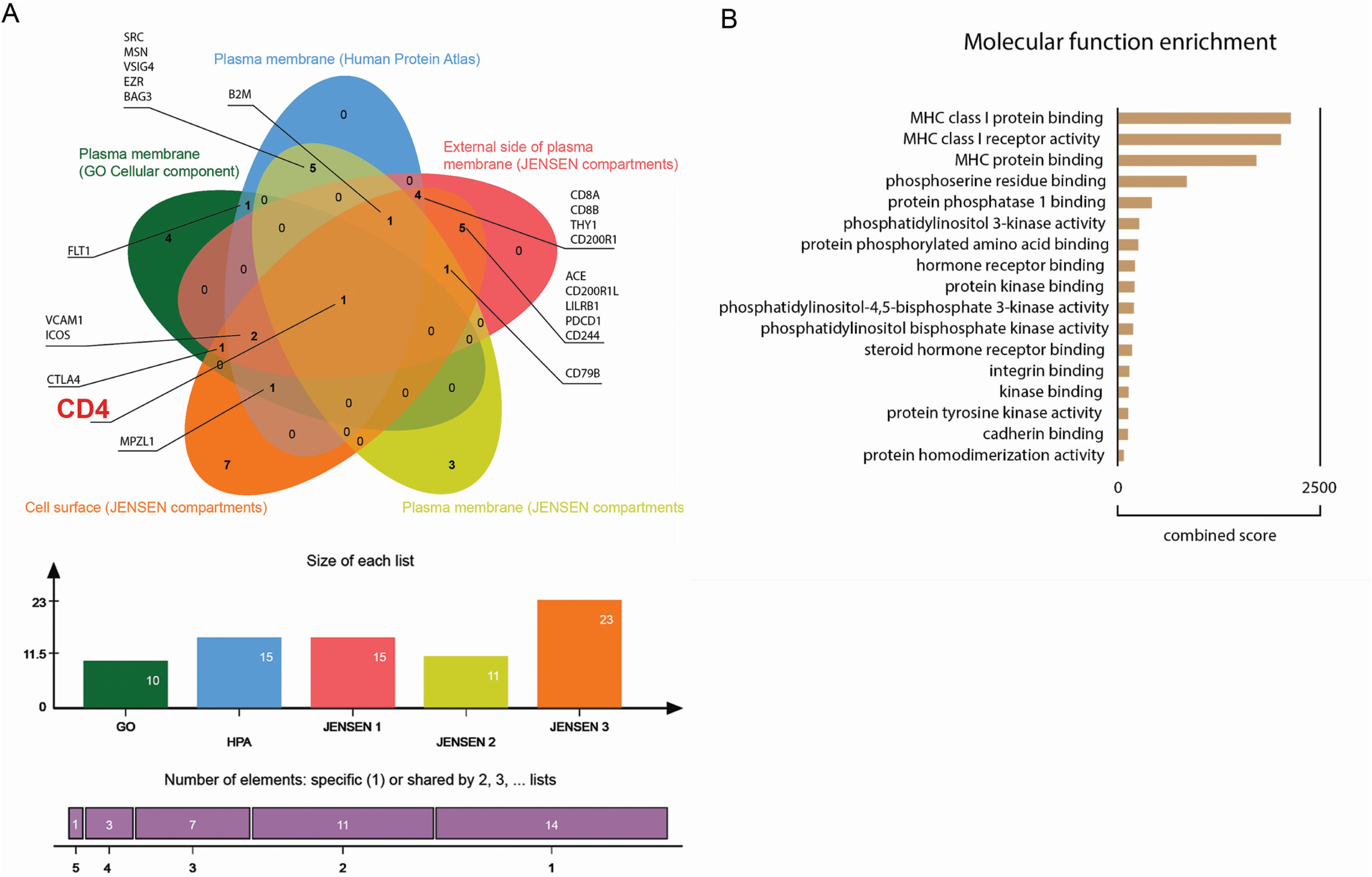
**(A)** Venn diagram of human proteins that were predicted by P-HIPSTer to interact with sCoV-1 and considered as part of cellular membrane in at least one database. Proteins found in at least two databases are annotated. (**B)** Molecular function enrichment of 71 predicted human proteins that interact with sCoV-1 according to P-HIPSTer.

**Extended Data Fig. 4.**
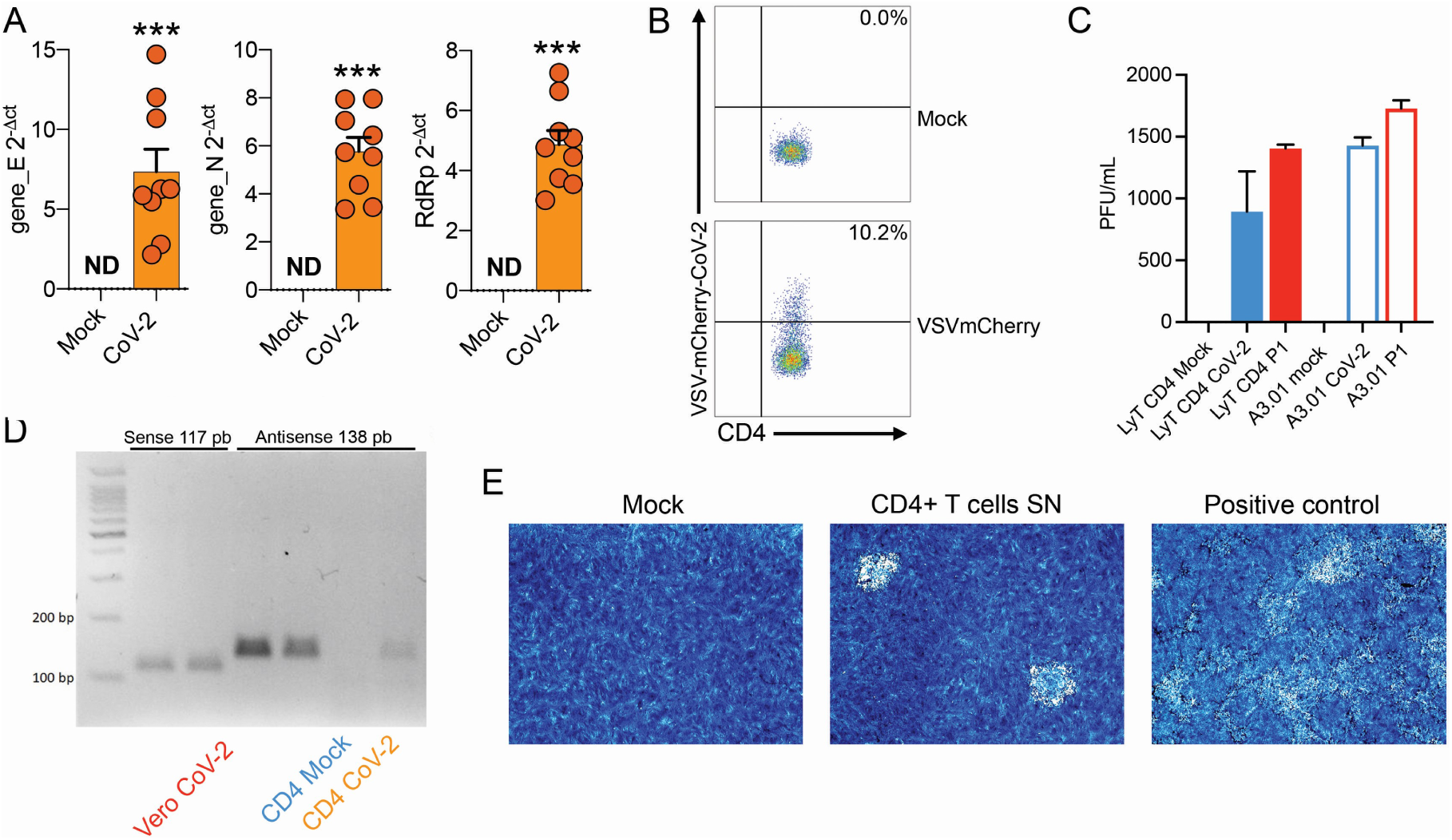
(**A**) Relative expression of the viral envelope (E), nucleocapsid (N) and RNA-dependent RNA polymerase (RdRp) genes in peripheral blood CD4 + T cells infected with mock control or CoV-2. **(B)** Primary human CD4+ T cells were incubated with pseudotype (VSV-mCherry-CoV-2). **(C)** Peripheral blood CD3^+^CD4^+^ was infected with mock control or SARS-CoV-2 (CoV-2) lineages (B and P.1) (MOI 1) for 1 h under continuous agitation. Viral load was assessed by RT-qPCR 24 h after infection. (**D**) PCR for the antisense CoV-2 strand was performed in CD4^+^ T and Vero (positive control) cells infected with mock or CoV-2. **(E)** Vero cells were incubated with the supernatant of mock control or CoV-2 infected CD4 + T cells under continuous agitation for 1 h. The viral load in Vero was measured after 72 h using plate assay. PFU – plaque-forming units.

**Extended Data Fig. 5.**
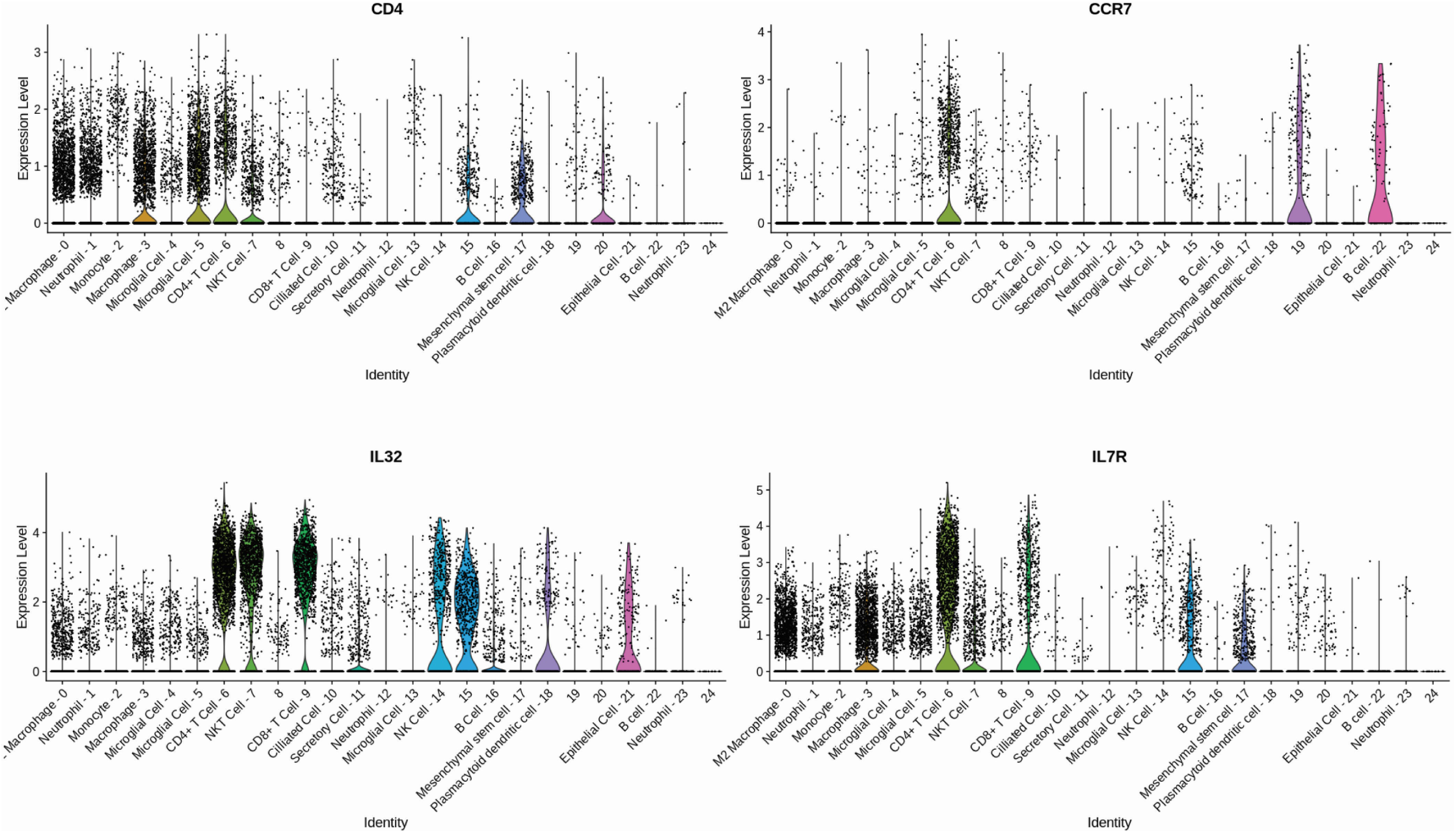
Gene signature of individual CD4+ T cells population. CD4+ T cell subpopulations were then annotated considering a high differential expression of CD4, CCR7, IL7R and IL32 gene markers.

**Extended Data Fig. 6.**
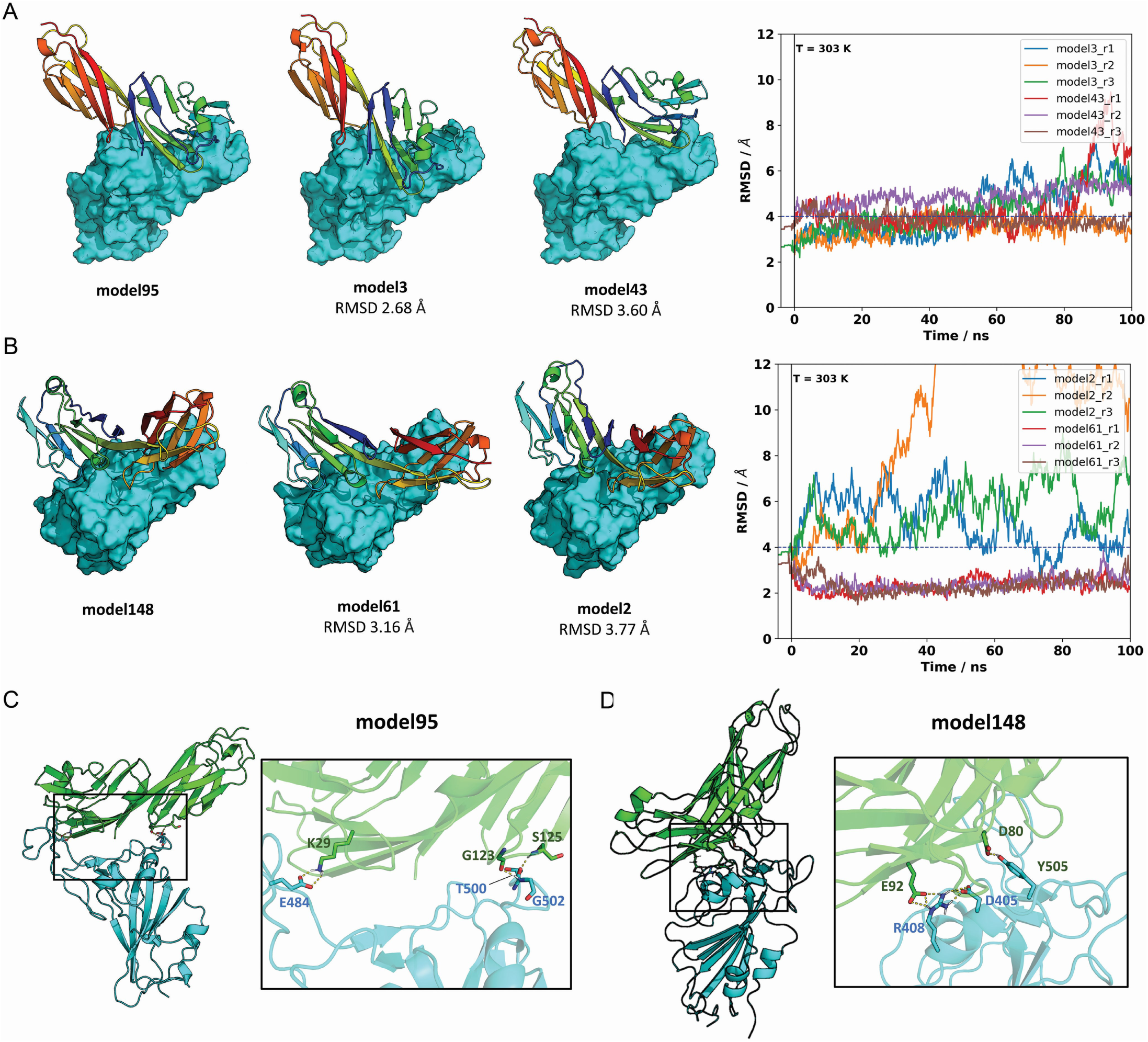
The two closest cluster members to model 95 **(A)** and model 148 **(B)** were subjected to 100 ns MD simulations at 303 K in triplicate. Neighboring models 3 and 43, in the RMSD range of ∼2.6 to 3.8 A to the reference model 95, represent closely related binding modes. Similarly, models 2 and 61 are closely related to cluster representative model 148. The right panels show the RMSD relative to the reference model as a function of trajectory time. Model 61, initially at 3.16 A from its cluster representative (model 148), converged to the structure of model 148 for all runs. This result provides additional evidence that model 148 is a likely candidate for CD4-RBD binding. **(C)** For model 95, three polar contacts are well defined in the interface region: CD4 lysine 29 is hydrogen-bonded to sCov2 glutamic acid 484; glycine carbonyl oxygen and amide hydrogen of serine 125 from CD4 make h-bonds to amide hydrogen of glycine 502 and threonine 500 of sCov2, respectively. (**D)** For model 148, two polar contacts are persistent: one hydrogen bond between CD4 aspartic acid 80 and sCov2 tyrosine 505 and a second hydrogen bond between CD4 glutamic acid 92 and sCov2 arginine 408 whose side chain orientation is also supported by a third residue, sCov2 aspartic acid 405, which completes a three-residue hydrogen bond network.

**Extended Data Fig. 7.**
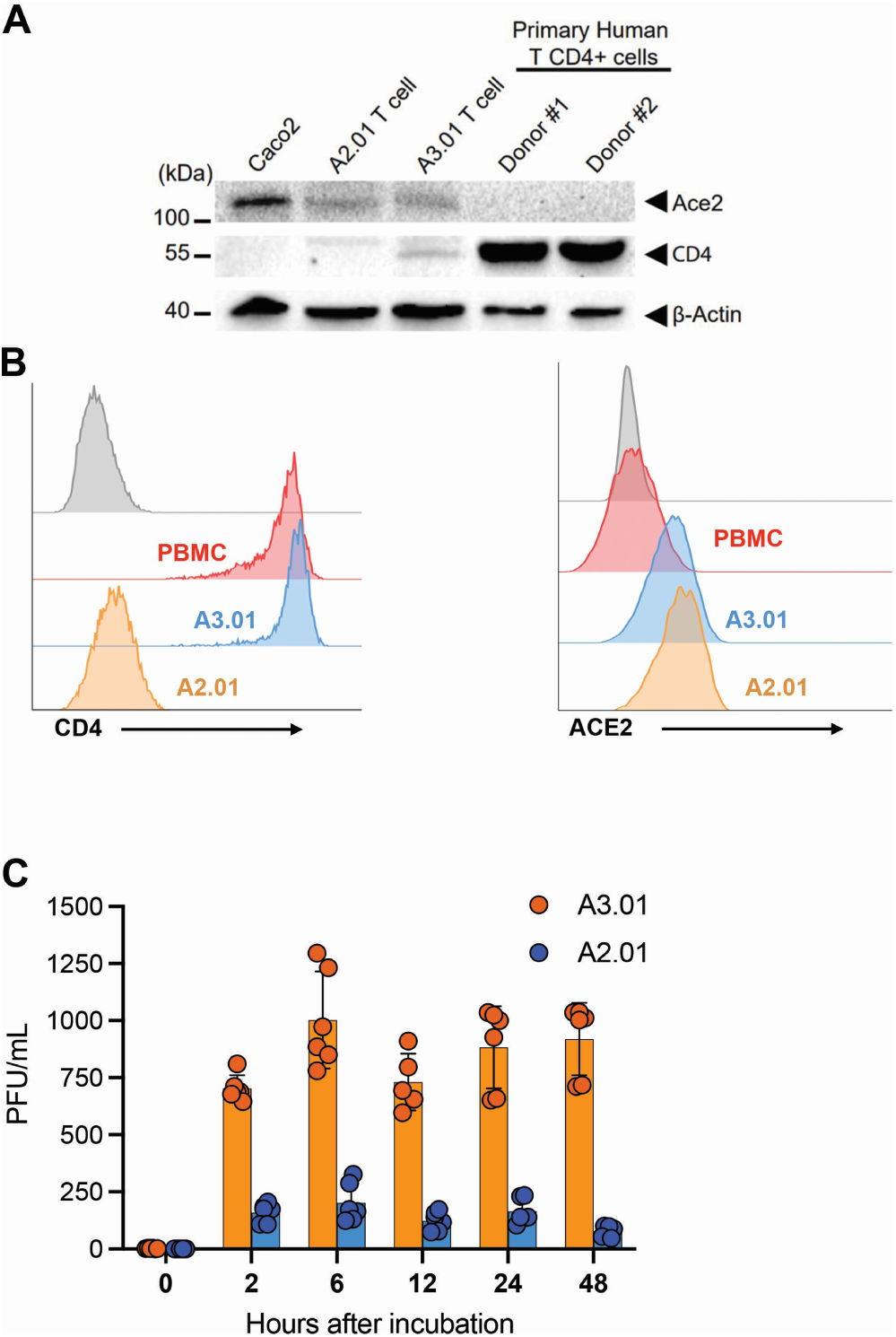
(**A**) Western Blotting and flow cytometry analysis of ACE2 and CD4 in Caco-2, A2.01, A3.01 cells and CD4+ T lymphocytes. **(B)** Flow cytometry analysis of CD4 (left) and ACE2 (right) expression primary CD4 + T cells (PBMC), A2.01 and A3.01.(**C**) Temporal viral load of SARS-CoV-2 in A2.01 and A3.01 lineages cells.

**Extended Data Fig. 8.**
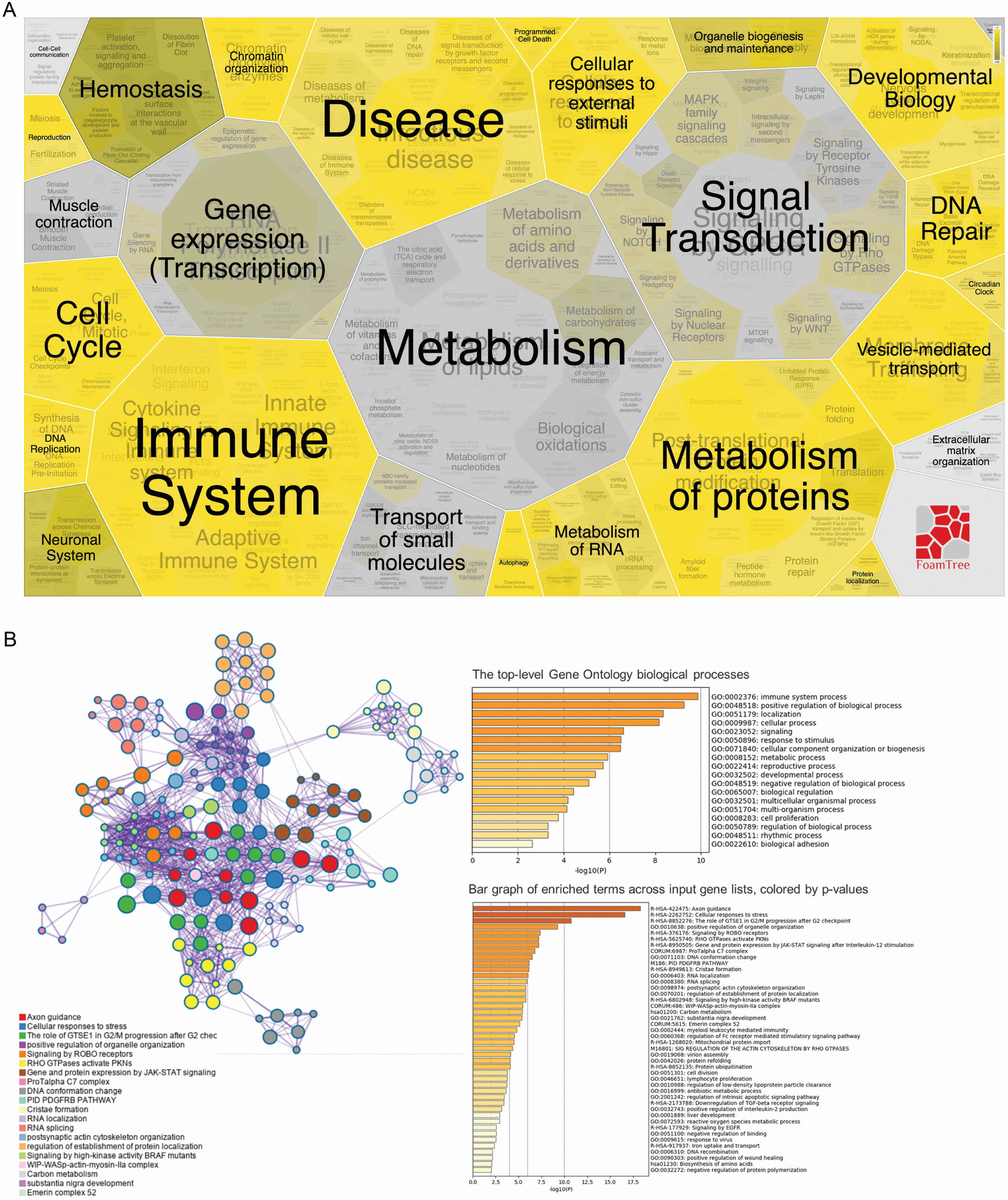
**(A)** Foam Tree according to the biological processes affected by the differentially expressed proteins{Jassal:2019hz}. **(B)** Network of enriched terms colored by cluster ID; nodes that share the same cluster ID are typically close to each other{Zhou:2019bm}. **(C)** Bar graph of enriched terms across input gene lists, colored by p-values{Zhou:2019bm}.

**Extended Data Fig. 9.**
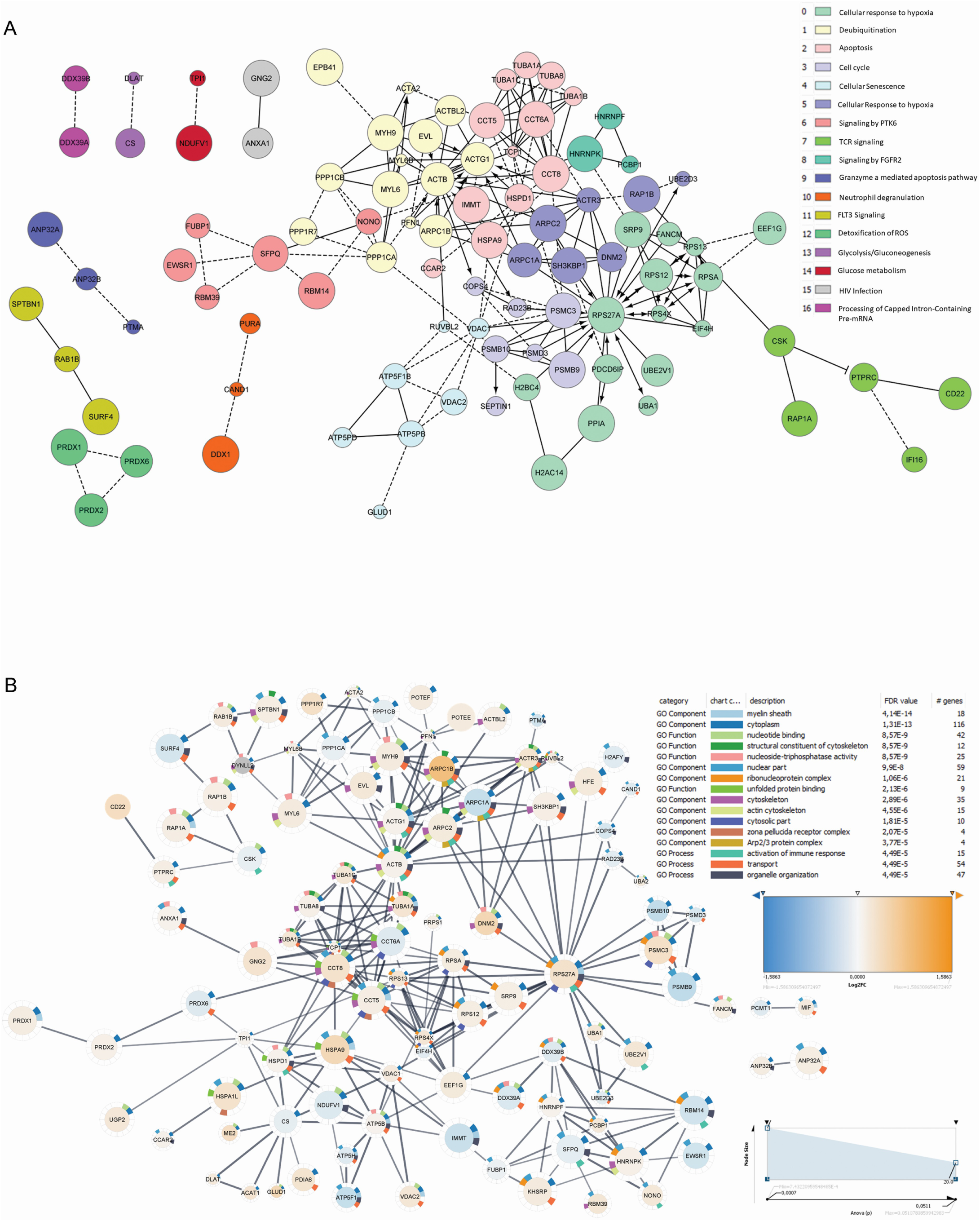
**(A)** Functional interaction network of proteins (identified by their gene name). **(B)** Protein interaction (identified by their gene name) of significantly altered proteins in infected CD4 + T cells with 0.7 confidence score and functional enrichment for top 5 gene ontology terms. Node size is related to p-value and node colors show the direction of expression.

**Extended Data Fig. 10.**
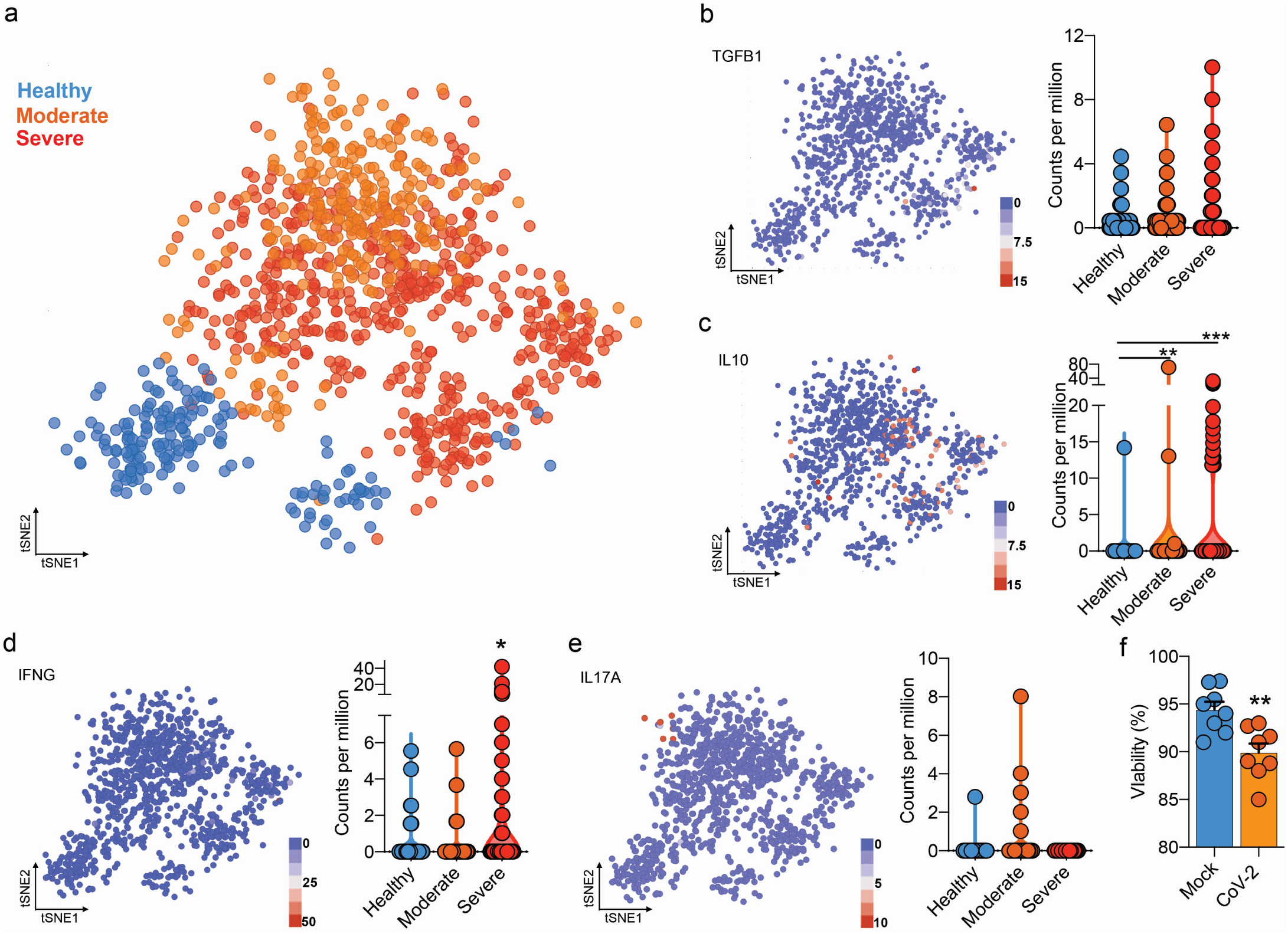
**(A)** Flowchart of the of t-SNE-based analysis{Strazar:2019bn} of human BAL (subclustered of CD4 + T cells) of Healthy, Moderate and Severe COVID-19 patients. **(B)** Gene expression of TGFB1 in counts per million. **(C)** Gene expression of IL10 in counts per million. **(D)** Gene expression of IFNG in counts per million. **(E)** Gene expression of IL17A in counts per million. Each dot represents a cell. **(F)** Viability of CD4+ T cells 24 h after incubation with mock (blue) or CoV-2 (orange). *** p < 0.001, ** p < 0.01, * p < 0.05

