## Supplementary material for "SARS-CoV-2 Uses CD4 to Infect T Helper Lymphocytes": Table S1

### Supplementary Table 1

| Subject | Gender | Age range (y) Mean (SD) | Admittance O2 saturation (%) Mean (SD) |
|---|---|---|---|
| **Healthy Donors** | | | |
| H19 20 000001 | M | 26-30 | - |
| H19 20 000002 | M | 41-45 | - |
| H19 20 000003 | M | 46-50 | - |
| H19 20 000004 | F | 51-55 | - |
| H19 20 000005 | M | 41-45 | - |
| H19 20 000006 | F | 36-40 | - |
| H19 20 000007 | M | 31-35 | - |
| H19 20 000008 | F | 26-30 | - |
| H19 20 000009 | M | 46-50 | - |
| | M/F: 6:3 | 36.4 (9.60) | |
| **COVID-19 Patients** | | | |
| C19 20 000001 | F | 36-40 | 86 ra |
| C19 20 000002 | F | 41-45 | 91 ra |
| C19 20 000003 | M | 36-40 | 100 nrb |
| C19 20 000004 | M | 61-65 | 92 ra |
| C19 20 000005 | F | 41-45 | 89 ra |
| C19 20 000006 | F | 76-80 | 90 ra |
| C19 20 000007 | F | 56-60 | 92 ra |
| C19 20 000008 | F | 56-60 | 92 ra |
| C19 20 000009 | F | 56-60 | 94 ra |
| C19 20 000010 | F | 36-40 | 90 ra |
| C19 20 000011 | M | 41-45 | 87 ra |
| C19 20 000012 | M | 76-80 | 86 ra |
| C19 20 000013 | F | 41-45 | 98 ra |
| C19 20 000014 | M | 31-35 | 92 ra |
| C19 20 000015 | M | 36-40 | 93 ra |
| C19 20 000016 | M | 66-70 | 87 ra |
| | M/F: 7:9 | 50.9 (15) | 91.2 (3.9) |

*Patients IDs are not identifying in the manuscript.
