## Supplementary material for "SARS-CoV-2 Uses CD4 to Infect T Helper Lymphocytes": Table S2

### Supplementary Table 2

| Accession | Peptide count | Unique peptides | Confidence score | Anova (p) | q Value | Max fold change | Power | Highest mean condition | Lowest mean condition | Description | TCell_Covid_T1 | TCell_Covid_T1_2 | TCell_Covid_T1_3 | TCell_Covid_T1_4 | TCell_Covid_T2 | TCell_Covid_T2_2 | TCell_Covid_T2_3 | TCell_Covid_T2_4 | Tcell_Mock_T1 | Tcell_Mock_T1_2 | Tcell_Mock_T1_3 | Tcell_Mock_T1_4 | Tcell_Mock_T2 | Tcell_Mock_T2_2 | Tcell_Mock_T2_3 | Tcell_Mock_T2_4 |
|---|---|---|---|---|---|---|---|---|---|---|---|---|---|---|---|---|---|---|---|---|---|---|---|---|---|---|
| Q92499 | 29 | 20 | 173,2808 | 6,257E-06 | 0,00200347 | 1,539248432 | 0,99999672 | MOCK | COVID | ATP-dependent RNA helicase DDX1 OS=Homo sapiens OX=9606 GN=DDX1 PE=1 SV=2 | 342583,3194 | 425063,6459 | 329087,4603 | 337830,4066 | 447929,5169 | 496038,3335 | 412128,0037 | 432093,4775 | 569559,1917 | 683818,748 | 635072,8777 | 658824,734 | 545488,1793 | 550696,8544 | 659661,8042 | 657496,9054 |
| Q16891 | 13 | 5 | 74,7282 | 4,8212E-05 | 0,00847867 | 1,642193556 | 0,99964405 | MOCK | COVID | MICOS complex subunit MIC60 OS=Homo sapiens OX=9606 GN=IMMT PE=1 SV=1 | 222135,398 | 260495,4765 | 272042,821 | 260573,6635 | 254084,2943 | 260725,9062 | 355988,0701 | 376579,504 | 413041,0113 | 438623,5404 | 509143,3244 | 543615,042 | 369891,5908 | 370044,3664 | 534027,6949 | 537281,8442 |
| Q96PK6 | 6 | 4 | 35,1185 | 0,00029518 | 0,01890337 | 1,497073646 | 0,99322479 | MOCK | COVID | RNA-binding protein 14 OS=Homo sapiens OX=9606 GN=RBM14 PE=1 SV=2 | 13580,83191 | 13511,46857 | 11083,42669 | 11005,40307 | 11800,05768 | 13517,07012 | 12477,51139 | 13178,04814 | 19310,9433 | 21693,55721 | 15227,17551 | 15104,42207 | 17795,56278 | 28048,72871 | 16530,23824 | 16227,01302 |
| O15260 | 3 | 1 | 24,8594 | 0,00055212 | 0,02705479 | 1,440777462 | 0,98483349 | MOCK | COVID | Surfeit locus protein 4 OS=Homo sapiens OX=9606 GN=SURF4 PE=1 SV=3 | 9125,447391 | 5820,682667 | 9036,991484 | 10058,71395 | 9013,314921 | 10312,68824 | 8617,666621 | 7551,066266 | 11350,55761 | 15826,38582 | 10138,83495 | 10599,27239 | 13369,35917 | 12107,44785 | 13153,21676 | 13641,65054 |
| P28065 | 8 | 4 | 58,371 | 0,00055738 | 0,02705479 | 1,831125139 | 0,98465856 | MOCK | COVID | Proteasome subunit beta type-9 OS=Homo sapiens OX=9606 GN=PSMB9 PE=1 SV=2 | 192043,9603 | 248821,9104 | 141765,5058 | 142149,513 | 311469,3532 | 336026,2583 | 237659,2591 | 232285,0575 | 381434,3041 | 577331,5128 | 507957,3699 | 504356,1438 | 373115,8928 | 410829,308 | 299615,0764 | 318697,2432 |
| P40227 | 20 | 15 | 179,955 | 0,00081181 | 0,02705479 | 1,300905301 | 0,97623998 | MOCK | COVID | T-complex protein 1 subunit zeta OS=Homo sapiens OX=9606 GN=CCT6A PE=1 SV=3 | 185450,6232 | 201993,269 | 193493,7193 | 223443,289 | 211423,1631 | 254129,2152 | 242421,8306 | 266531,9265 | 262272,3573 | 261575,7117 | 294196,0961 | 304895,401 | 256702,6936 | 259782,3269 | 337053,7144 | 337685,2746 |
| Q08334 | 1 | 1 | 5,3036 | 0,00082875 | 0,02705479 | 2,133275938 | 0,97568524 | MOCK | COVID | Interleukin-10 receptor subunit beta OS=Homo sapiens OX=9606 GN=IL10RB PE=1 SV=2 | 1191,400454 | 2161,812122 | 1226,995784 | 1263,110526 | 2133,829934 | 1909,478201 | 1440,102371 | 1409,201133 | 3408,730669 | 2738,300964 | 1536,915688 | 2115,528044 | 4209,710535 | 5509,724821 | 3802,200736 | 3848,142682 |
| Q6UB99 | 10 | 4 | 55,1211 | 0,00127457 | 0,02932251 | 2,406148216 | 0,96149647 | MOCK | COVID | Ankyrin repeat domain-containing protein 11 OS=Homo sapiens OX=9606 GN=ANKRD11 PE=1 SV=3 | 16600,65314 | 16431,1074 | 23549,02873 | 26798,22204 | 44238,66517 | 41243,43408 | 66827,10544 | 67157,4369 | 105260,2932 | 88825,33976 | 113358,5312 | 133104,2045 | 62613,96993 | 41455,82991 | 87006,70651 | 97066,65233 |
| P49821 | 9 | 3 | 60,325 | 0,00142054 | 0,02962052 | 1,314005021 | 0,95704116 | MOCK | COVID | NADH dehydrogenase [ubiquinone] flavoprotein 1_ mitochondrial OS=Homo sapiens OX=9606 GN=NDUFV1 PE=1 SV=4 | 124099,9275 | 131068,6975 | 136415,3129 | 126484,6204 | 129273,9276 | 160696,2962 | 172805,1802 | 186476,0263 | 147695,0533 | 209148,1061 | 201596,7005 | 183395,2578 | 208462,4515 | 181250,5561 | 213496,4682 | 188819,7323 |
| Q16777 | 9 | 1 | 145,6485 | 0,00169755 | 0,033972 | 1,347851228 | 0,9488464 | MOCK | COVID | Histone H2A type 2-C OS=Homo sapiens OX=9606 GN=H2AC20 PE=1 SV=4 | 2001250,16 | 2430763,893 | 2543078,003 | 2496980,442 | 2474840,756 | 3379269,437 | 3014097,617 | 3288243,996 | 2756349,438 | 3792385,24 | 3402914,836 | 3405408,336 | 3780864,021 | 3661664,069 | 4290566,787 | 4061880,323 |
| P23246 | 26 | 16 | 203,1556 | 0,00181808 | 0,03528141 | 1,200835217 | 0,9453843 | MOCK | COVID | Splicing factor_ proline- and glutamine-rich OS=Homo sapiens OX=9606 GN=SFPQ PE=1 SV=2 | 246911,5904 | 251358,1981 | 247030,5829 | 262088,3673 | 279775,9596 | 316202,2729 | 301777,1573 | 335548,2275 | 337873,4019 | 318869,8849 | 349976,7742 | 327843,8036 | 300005,585 | 378603,4192 | 345960,7764 | 331568,6464 |
| Q99878 | 23 | 11 | 137,8538 | 0,0021945 | 0,03772822 | 1,352969859 | 0,93495106 | MOCK | COVID | Histone H2A type 1-J OS=Homo sapiens OX=9606 GN=H2AC14 PE=1 SV=3 | 2081165,228 | 2535014,817 | 2682380,748 | 2654883,976 | 2659750,752 | 3600490,923 | 3295253,966 | 3530207,204 | 2913356,163 | 4056941,819 | 3643593,884 | 3693564,872 | 3998850,52 | 3863568,036 | 4586258,169 | 4415138,845 |
| Q92747 | 3 | 0 | 16,5805 | 0,00503265 | 0,07006271 | 1,705274403 | 0,87024445 | MOCK | COVID | Actin-related protein 2/3 complex subunit 1A OS=Homo sapiens OX=9606 GN=ARPC1A PE=2 SV=2 | 32539,24855 | 21804,69366 | 21665,3624 | 30613,33888 | 69030,92938 | 91242,40667 | 53371,85678 | 57263,22384 | 70544,34826 | 100508,1358 | 82727,0538 | 70756,82672 | 79701,29142 | 91536,32588 | 76114,47287 | 71905,59861 |
| Q5JTV8 | 6 | 3 | 39,181 | 0,00707005 | 0,07957215 | 1,977068967 | 0,83385995 | MOCK | COVID | Torsin-1A-interacting protein 1 OS=Homo sapiens OX=9606 GN=TOR1AIP1 PE=1 SV=2 | 64539,26604 | 38396,19669 | 77656,0893 | 75078,549 | 53817,92862 | 47776,23635 | 105485,2052 | 125750,7889 | 177449,9349 | 93770,46338 | 204583,6625 | 179897,7441 | 125024,3363 | 52021,08624 | 168359,6815 | 162398,6925 |
| Q01844 | 18 | 8 | 119,8252 | 0,00745529 | 0,07957215 | 1,510760577 | 0,82762945 | MOCK | COVID | RNA-binding protein EWS OS=Homo sapiens OX=9606 GN=EWSR1 PE=1 SV=1 | 67178,43059 | 64608,89588 | 91646,53506 | 109385,5942 | 101062,1562 | 82968,97462 | 136560,6115 | 130683,3773 | 153480,6742 | 129519,4716 | 183512,0761 | 177007,2191 | 101853,7517 | 96095,90231 | 170182,7456 | 172927,3328 |
| P30041 | 15 | 9 | 140,1295 | 0,00915761 | 0,08814793 | 1,294338636 | 0,80205906 | MOCK | COVID | Peroxiredoxin-6 OS=Homo sapiens OX=9606 GN=PRDX6 PE=1 SV=3 | 192596,8696 | 209964,3813 | 192892,9087 | 185014,2774 | 222938,6225 | 236806,6316 | 250507,2381 | 262682,8611 | 257733,4687 | 294238,7745 | 363493,6811 | 306811,8958 | 244449,4894 | 191922,043 | 312306,1686 | 298542,7494 |
| P41240 | 16 | 8 | 104,7941 | 0,01078765 | 0,09735873 | 1,164899369 | 0,78008285 | MOCK | COVID | Tyrosine-protein kinase CSK OS=Homo sapiens OX=9606 GN=CSK PE=1 SV=1 | 118812,5397 | 131724,3283 | 119370,7184 | 117799,1893 | 144240,5632 | 135572,3276 | 142647,0708 | 164573,3872 | 141667,808 | 165816,3975 | 169010,1669 | 154634,2822 | 133525,1383 | 176435,7286 | 158394,5091 | 152480,0618 |
| P62136 | 13 | 2 | 97,1703 | 0,01137449 | 0,10116897 | 1,160286836 | 0,77267458 | MOCK | COVID | Serine/threonine-protein phosphatase PP1-alpha catalytic subunit OS=Homo sapiens OX=9606 GN=PPP1CA PE=1 SV=1 | 137829,5294 | 151790,0954 | 141331,7693 | 132727,7567 | 165985,8564 | 178129,4143 | 159040,5729 | 163031,8099 | 185831,8947 | 209356,3236 | 187075,0154 | 189817,3614 | 152139,1963 | 174108,2279 | 161768,5619 | 166901,6816 |
| O00148 | 26 | 11 | 151,3534 | 0,01298471 | 0,10941243 | 1,365274119 | 0,75352126 | MOCK | COVID | ATP-dependent RNA helicase DDX39A OS=Homo sapiens OX=9606 GN=DDX39A PE=1 SV=2 | 337880,5222 | 492952,9282 | 626724,7031 | 535976,2926 | 318855,3065 | 290821,1012 | 293581,0644 | 326167,484 | 664396,0987 | 421450,8661 | 607662,8593 | 657647,2577 | 490294,0503 | 529804,3585 | 521727,229 | 507240,3398 |
| O75390 | 10 | 5 | 68,7388 | 0,0144894 | 0,11663779 | 1,194675331 | 0,73698325 | MOCK | COVID | Citrate synthase_ mitochondrial OS=Homo sapiens OX=9606 GN=CS PE=1 SV=2 | 74265,15324 | 79653,26207 | 99382,14294 | 102265,8858 | 89008,46322 | 80184,2711 | 73130,05303 | 93858,62647 | 102148,2013 | 120828,2001 | 113700,0382 | 104396,5899 | 98087,53679 | 78912,45349 | 107721,5583 | 100619,5229 |
| Q7Z4W1 | 2 | 2 | 12,9957 | 0,01492638 | 0,11663779 | 7,012182231 | 0,73239813 | MOCK | COVID | L-xylulose reductase OS=Homo sapiens OX=9606 GN=DCXR PE=1 SV=2 | 139,8335954 | 305,7137888 | 7,033775927 | 0 | 147,2109062 | 394,4480765 | 21,23544713 | 98,32608583 | 759,3489837 | 1065,104712 | 67,14300201 | 127,5373283 | 1799,936418 | 2790,764666 | 596,8833395 | 603,4618697 |
| P62140 | 11 | 1 | 86,6422 | 0,01633288 | 0,12022409 | 1,193323228 | 0,71823755 | MOCK | COVID | Serine/threonine-protein phosphatase PP1-beta catalytic subunit OS=Homo sapiens OX=9606 GN=PPP1CB PE=1 SV=3 | 115162,9718 | 130818,2832 | 114706,1523 | 112648,0985 | 153947,1094 | 163643,5384 | 148332,4657 | 153226,3597 | 168541,113 | 192814,415 | 172715,5033 | 174024,5022 | 139746,5146 | 162432,2906 | 143386,7419 | 150026,6206 |
| O75367 | 10 | 5 | 83,4891 | 0,01841835 | 0,12820679 | 1,275497303 | 0,69873653 | MOCK | COVID | Core histone macro-H2A.1 OS=Homo sapiens OX=9606 GN=MACROH2A1 PE=1 SV=4 | 44402,08082 | 59907,27386 | 57895,88636 | 52243,92711 | 69193,1928 | 94590,53485 | 74356,67084 | 81031,39676 | 88992,33129 | 79538,78311 | 76063,8877 | 75496,9472 | 96915,50705 | 107109,7845 | 78177,40978 | 78337,44901 |
| Q8NHP1 | 6 | 1 | 40,2846 | 0,01913934 | 0,12850342 | 1,691162008 | 0,69236365 | MOCK | COVID | Aflatoxin B1 aldehyde reductase member 4 OS=Homo sapiens OX=9606 GN=AKR7L PE=2 SV=7 | 190886,4067 | 345811,7731 | 495832,3375 | 404432,548 | 97959,88675 | 80698,18532 | 92033,00303 | 99969,60581 | 498197,428 | 232395,5153 | 453111,9832 | 497098,3099 | 320861,2634 | 326826,1185 | 379112,8348 | 349381,1507 |
| P62807 | 19 | 0 | 296,2216 | 0,01926362 | 0,12850342 | 1,201256414 | 0,69128278 | MOCK | COVID | Histone H2B type 1-C/E/F/G/I OS=Homo sapiens OX=9606 GN=H2BC4 PE=1 SV=4 | 1287046,014 | 1605577,958 | 1601882,769 | 1583092,349 | 2158268,283 | 2025864,953 | 1784873,308 | 1936962,064 | 1956663,892 | 2010049,61 | 1829151,541 | 1821261,293 | 2186894,763 | 2521055,33 | 2341032,35 | 2131741,607 |
| P40306 | 6 | 2 | 37,6859 | 0,01975683 | 0,13005855 | 1,795229341 | 0,687043 | MOCK | COVID | Proteasome subunit beta type-10 OS=Homo sapiens OX=9606 GN=PSMB10 PE=1 SV=1 | 141724,9851 | 163703,076 | 116374,7534 | 114133,2156 | 45735,2354 | 63174,45316 | 32126,92469 | 32473,81701 | 95228,71343 | 181769,1414 | 108241,5302 | 93348,52646 | 182684,5868 | 242115,2392 | 204631,8498 | 165599,5139 |
| Q8IUH2 | 2 | 2 | 16,2927 | 0,02026924 | 0,13104746 | 1,534966428 | 0,68271967 | MOCK | COVID | Protein CREG2 OS=Homo sapiens OX=9606 GN=CREG2 PE=1 SV=1 | 20539,60788 | 23939,21118 | 15894,76054 | 15014,60249 | 11156,12149 | 11076,23363 | 9012,036642 | 10188,34434 | 25347,07959 | 33214,59957 | 15442,81675 | 14297,82825 | 26678,97403 | 29251,30335 | 16989,39216 | 18094,19376 |
| P24539 | 3 | 1 | 21,8855 | 0,02046353 | 0,13104746 | 1,719547777 | 0,68110148 | MOCK | COVID | ATP synthase F(0) complex subunit B1_ mitochondrial OS=Homo sapiens OX=9606 GN=ATP5PB PE=1 SV=2 | 3945,213553 | 6280,230645 | 8302,002647 | 12326,5568 | 12243,73752 | 9392,446892 | 16530,14208 | 15868,06294 | 21843,92922 | 9193,600713 | 22048,40396 | 20866,87963 | 13972,74065 | 10569,60651 | 26162,69793 | 21311,78904 |
| Q13838 | 19 | 6 | 171,5072 | 0,02168863 | 0,13227898 | 1,297003128 | 0,67115286 | MOCK | COVID | Spliceosome RNA helicase DDX39B OS=Homo sapiens OX=9606 GN=DDX39B PE=1 SV=1 | 329276,8643 | 486795,9411 | 634852,4595 | 537388,101 | 377831,82 | 368100,3829 | 341357,2144 | 376595,9521 | 685742,4685 | 405526,8727 | 635760,7927 | 679134,9279 | 498453,5215 | 512602,3913 | 542417,7283 | 517873,8556 |
| Q96AE4 | 26 | 14 | 192,9837 | 0,02331804 | 0,13457303 | 1,120658242 | 0,65855615 | MOCK | COVID | Far upstream element-binding protein 1 OS=Homo sapiens OX=9606 GN=FUBP1 PE=1 SV=3 | 204992,3643 | 246518,217 | 215148,9309 | 206922,9111 | 222248,8632 | 258279,2458 | 234715,0805 | 255634,8396 | 250779,4455 | 267821,5843 | 222933,4348 | 234611,4504 | 278136,1403 | 291208,4507 | 262443,9613 | 259075,3408 |
| Q16778 | 26 | 8 | 279,0157 | 0,02353234 | 0,13457303 | 1,221474343 | 0,65694977 | MOCK | COVID | Histone H2B type 2-E OS=Homo sapiens OX=9606 GN=H2BC21 PE=1 SV=3 | 1175465,004 | 1605689,792 | 1449289,618 | 1429832,351 | 2092850,344 | 2039594,812 | 1740428,179 | 1845723,194 | 1849761,317 | 2043453,251 | 1739843,34 | 1782167,152 | 2060840,61 | 2528568,675 | 2271398,779 | 2065917,349 |
| Q00577 | 3 | 3 | 16,1835 | 0,03077718 | 0,16223784 | 1,72860792 | 0,60837194 | MOCK | COVID | Transcriptional activator protein Pur-alpha OS=Homo sapiens OX=9606 GN=PURA PE=1 SV=2 | 6593,836189 | 6426,85478 | 4706,884237 | 7304,777893 | 1543,838032 | 1995,782543 | 3196,127913 | 3234,898524 | 4500,424802 | 4826,525632 | 12332,69723 | 8111,885386 | 5333,117476 | 4850,422324 | 8795,890837 | 11755,49954 |
| P43487 | 3 | 2 | 16,737 | 0,03090751 | 0,16223784 | 1,287371099 | 0,60758618 | MOCK | COVID | Ran-specific GTPase-activating protein OS=Homo sapiens OX=9606 GN=RANBP1 PE=1 SV=1 | 35199,96415 | 35551,54606 | 26304,37218 | 25411,88648 | 23098,28691 | 31772,34068 | 20636,25738 | 20422,06245 | 31401,19807 | 41353,61061 | 25465,74712 | 26630,97621 | 36986,25913 | 45473,15541 | 36867,87476 | 36978,79939 |
| O75947 | 12 | 5 | 123,1735 | 0,03188416 | 0,16466499 | 1,266889433 | 0,60178241 | MOCK | COVID | ATP synthase subunit d_ mitochondrial OS=Homo sapiens OX=9606 GN=ATP5PD PE=1 SV=3 | 136193,5293 | 162730,6798 | 160310,8958 | 148736,7986 | 105608,3923 | 120750,3115 | 110758,3739 | 111040,0328 | 138417,2883 | 143818,146 | 147648,7359 | 122631,5017 | 183967,5697 | 169233,1898 | 223480,439 | 208801,8178 |
| P54727 | 8 | 6 | 62,9991 | 0,03407226 | 0,16479976 | 1,224586395 | 0,58929426 | MOCK | COVID | UV excision repair protein RAD23 homolog B OS=Homo sapiens OX=9606 GN=RAD23B PE=1 SV=1 | 54323,15456 | 40467,01915 | 68758,43139 | 66587,90826 | 48905,85856 | 58706,78114 | 59600,39015 | 62855,12138 | 63785,47196 | 57220,36186 | 73211,88211 | 70781,2751 | 75230,88858 | 53562,04885 | 82762,44847 | 87005,99418 |
| O95932 | 4 | 1 | 25,0076 | 0,03414587 | 0,16479976 | 1,369867203 | 0,58888585 | MOCK | COVID | Protein-glutamine gamma-glutamyltransferase 6 OS=Homo sapiens OX=9606 GN=TGM6 PE=1 SV=3 | 4981,899379 | 4166,293042 | 6255,928729 | 7650,851148 | 10269,96742 | 9049,688943 | 9107,777027 | 10998,52903 | 11940,42564 | 9212,874216 | 14093,93842 | 12828,82323 | 7404,157117 | 8290,858221 | 10917,47235 | 10902,03409 |
| Q9BT78 | 3 | 2 | 17,0035 | 0,03460106 | 0,16479976 | 1,396926829 | 0,5863768 | MOCK | COVID | COP9 signalosome complex subunit 4 OS=Homo sapiens OX=9606 GN=COPS4 PE=1 SV=1 | 7937,356259 | 8342,693915 | 6897,329754 | 6646,278315 | 5425,493139 | 5657,890329 | 4251,676403 | 3794,392005 | 7388,561931 | 10180,23081 | 6203,425663 | 5100,407909 | 10716,82374 | 11976,80925 | 8340,688362 | 8476,96521 |
| P22061 | 6 | 3 | 63,6515 | 0,03560083 | 0,16763676 | 1,302502915 | 0,58096173 | MOCK | COVID | Protein-L-isoaspartate(D-aspartate) O-methyltransferase OS=Homo sapiens OX=9606 GN=PCMT1 PE=1 SV=4 | 73302,16407 | 121262,4355 | 124039,3816 | 123849,4947 | 67201,87702 | 67018,00964 | 51678,01721 | 53961,69813 | 119061,4308 | 98103,6781 | 117025,8225 | 133229,7253 | 95153,29286 | 115705,3078 | 102420,5709 | 108014,9445 |
| O43242 | 8 | 5 | 41,644 | 0,03686765 | 0,17233479 | 1,368065321 | 0,57428182 | MOCK | COVID | 26S proteasome non-ATPase regulatory subunit 3 OS=Homo sapiens OX=9606 GN=PSMD3 PE=1 SV=2 | 42878,6839 | 65081,57024 | 46229,44173 | 34563,93824 | 84158,73878 | 62217,1981 | 71679,8444 | 82223,1732 | 83704,37282 | 102682,7127 | 68260,82513 | 51900,96043 | 93851,00912 | 112385,0025 | 82859,44478 | 73384,19791 |
| Q93079 | 47 | 25 | 289,6837 | 0,037773 | 0,17278317 | 1,205135418 | 0,56962657 | MOCK | COVID | Histone H2B type 1-H OS=Homo sapiens OX=9606 GN=H2BC9 PE=1 SV=3 | 1463240,503 | 1893837,115 | 2184516,532 | 2179039,043 | 2789932,706 | 2372461,819 | 2656235,727 | 2880535,546 | 2461110,207 | 2453289,675 | 2612654,736 | 2731152,925 | 2595179,831 | 2955536,554 | 3216682,556 | 3172745,673 |
| Q15056 | 5 | 3 | 37,1306 | 0,03905598 | 0,17613563 | 1,244153019 | 0,56319051 | MOCK | COVID | Eukaryotic translation initiation factor 4H OS=Homo sapiens OX=9606 GN=EIF4H PE=1 SV=5 | 16187,58989 | 19977,82876 | 16808,42536 | 14242,69291 | 17868,13341 | 18346,08075 | 12751,9766 | 12723,39783 | 21667,59798 | 17699,92952 | 15764,81048 | 14893,29334 | 23092,91333 | 27635,03021 | 19894,66527 | 19730,70507 |
| P61077 | 3 | 1 | 21,702 | 0,04230919 | 0,18431701 | 1,235196658 | 0,54765539 | MOCK | COVID | Ubiquitin-conjugating enzyme E2 D3 OS=Homo sapiens OX=9606 GN=UBE2D3 PE=1 SV=1 | 26988,38204 | 24948,24324 | 33316,11016 | 30577,54476 | 40754,29674 | 41727,67977 | 44474,37388 | 47229,27985 | 39295,99151 | 42909,25087 | 44440,16396 | 50645,89838 | 38501,74291 | 36336,97026 | 49101,62385 | 56995,04153 |
| P06454 | 4 | 2 | 29,4505 | 0,04317008 | 0,18599542 | 1,518551055 | 0,54371936 | MOCK | COVID | Prothymosin alpha OS=Homo sapiens OX=9606 GN=PTMA PE=1 SV=2 | 1350,145048 | 6403,639219 | 1380,92651 | 6010,712662 | 18555,9891 | 3844,286359 | 24784,39397 | 29139,46038 | 17450,79612 | 19519,11489 | 18268,29331 | 17268,68035 | 13046,07221 | 16919,76959 | 18411,76494 | 18016,6952 |
| Q86VP6 | 27 | 16 | 163,1445 | 0,04356578 | 0,18599542 | 1,26383171 | 0,54193336 | MOCK | COVID | Cullin-associated NEDD8-dissociated protein 1 OS=Homo sapiens OX=9606 GN=CAND1 PE=1 SV=2 | 455988,166 | 451217,6893 | 451956,1914 | 397035,275 | 759851,6793 | 884103,7806 | 823919,0079 | 917523,2378 | 868006,5144 | 883506,2469 | 868439,9017 | 748383,3906 | 727261,4323 | 765017,5106 | 850455,5301 | 787040,3089 |
| P50990 | 18 | 9 | 129,5723 | 6,6553E-07 | 0,0004262 | 1,436322147 | 1 | COVID | MOCK | T-complex protein 1 subunit theta OS=Homo sapiens OX=9606 GN=CCT8 PE=1 SV=4 | 195839,2631 | 216988,4629 | 164677,1525 | 158621,0857 | 193469,9226 | 205535,5584 | 187823,6685 | 192938,2655 | 124002,299 | 140103,7861 | 125679,778 | 129985,1544 | 133903,6824 | 145184,8903 | 126175,6269 | 130364,0761 |
| P38646 | 38 | 28 | 309,1544 | 5,2959E-05 | 0,00847867 | 1,741236766 | 0,99957477 | COVID | MOCK | Stress-70 protein_ mitochondrial OS=Homo sapiens OX=9606 GN=HSPA9 PE=1 SV=2 | 617781,7852 | 718417,6913 | 673706,8634 | 622786,5127 | 407212,9948 | 470521,9965 | 392974,6131 | 413082,612 | 311289,7646 | 317425,8373 | 318259,8321 | 325741,9498 | 264784,0184 | 279609,0724 | 330733,8946 | 331132,1564 |
| P51858 | 4 | 4 | 29,4206 | 0,00012868 | 0,01648169 | 1,629871036 | 0,99801655 | COVID | MOCK | Hepatoma-derived growth factor OS=Homo sapiens OX=9606 GN=HDGF PE=1 SV=1 | 305395,7232 | 413827,2536 | 384877,4671 | 376112,8041 | 409151,5478 | 450294,6049 | 335585,3189 | 385412,548 | 202882,1264 | 273384,529 | 175737,6362 | 175631,8269 | 266013,2275 | 360074,3612 | 204012,3224 | 220116,3892 |
| P11171 | 7 | 4 | 37,196 | 0,00017441 | 0,01861549 | 1,526370549 | 0,99681927 | COVID | MOCK | Protein 4.1 OS=Homo sapiens OX=9606 GN=EPB41 PE=1 SV=4 | 61955,58614 | 55853,56967 | 80057,39255 | 78466,98186 | 53466,91344 | 51043,46909 | 56559,66102 | 65682,51968 | 48490,53428 | 33579,3679 | 47790,61522 | 44701,99491 | 37755,38439 | 31219,88237 | 42443,61294 | 43614,91327 |
| P61224 | 13 | 1 | 101,6347 | 0,00022316 | 0,01890337 | 1,167591925 | 0,99542468 | COVID | MOCK | Ras-related protein Rap-1b OS=Homo sapiens OX=9606 GN=RAP1B PE=1 SV=1 | 132036,6768 | 138568,3688 | 128371,9555 | 138775,1603 | 138525,2584 | 136333,8162 | 151642,932 | 158723,9529 | 117992,0934 | 119759,0582 | 124884,7093 | 128384,2133 | 107289,0959 | 119828,3469 | 118758,0899 | 124894,293 |
| P62937 | 47 | 28 | 421,0777 | 0,0002473 | 0,01890337 | 1,23116083 | 0,99470236 | COVID | MOCK | Peptidyl-prolyl cis-trans isomerase A OS=Homo sapiens OX=9606 GN=PPIA PE=1 SV=2 | 2444081,61 | 2562061,199 | 2766197,686 | 2674132,979 | 2347025,392 | 1995842,618 | 2222888,087 | 2414219,764 | 1916795,245 | 2015622,698 | 2085220,929 | 2100076,752 | 2002686,837 | 1755472,141 | 1946433,432 | 1956661,582 |
| O15144 | 19 | 12 | 160,1736 | 0,00028095 | 0,01890337 | 1,245137027 | 0,99366922 | COVID | MOCK | Actin-related protein 2/3 complex subunit 2 OS=Homo sapiens OX=9606 GN=ARPC2 PE=1 SV=1 | 210394,6649 | 225316,641 | 189462,4444 | 183208,1361 | 175445,2425 | 189740,4896 | 165828,4934 | 170101,3125 | 158728,9572 | 138884,6198 | 153491,8226 | 141748,5792 | 161436,3473 | 136124,5663 | 161844,8969 | 160054,5112 |
| P42167 | 23 | 11 | 162,0464 | 0,00038198 | 0,02223799 | 1,176385668 | 0,99044649 | COVID | MOCK | Lamina-associated polypeptide 2_ isoforms beta/gamma OS=Homo sapiens OX=9606 GN=TMPO PE=1 SV=2 | 175341,8993 | 196826,8005 | 190158,0697 | 180133,2105 | 165602,2769 | 175496,6749 | 203853,5807 | 208782,1662 | 152240,348 | 143932,7544 | 160545,9583 | 167612,1838 | 155235,2336 | 157768,2297 | 168022,9333 | 166499,6409 |
| P60660 | 17 | 10 | 157,8662 | 0,00061059 | 0,02705479 | 1,232277549 | 0,98289002 | COVID | MOCK | Myosin light polypeptide 6 OS=Homo sapiens OX=9606 GN=MYL6 PE=1 SV=2 | 340597,0836 | 401554,7043 | 357008,4475 | 367261,1062 | 306156,5862 | 369533,0887 | 329463,109 | 353814,4627 | 296636,5627 | 348526,2611 | 283513,3538 | 310912,3682 | 255282,7006 | 281973,8233 | 263269,6066 | 252703,6826 |
| Q06830 | 29 | 18 | 244,7585 | 0,0006586 | 0,02705479 | 1,204251347 | 0,98129607 | COVID | MOCK | Peroxiredoxin-1 OS=Homo sapiens OX=9606 GN=PRDX1 PE=1 SV=1 | 530217,6401 | 506105,8186 | 464674,3698 | 437310,1003 | 444399,4955 | 436548,6173 | 513429,8935 | 536597,2635 | 390404,1032 | 385601,5818 | 424477,1516 | 431925,3125 | 335510,9499 | 409091,6843 | 410938,0484 | 425070,7834 |
| P62979 | 4 | 3 | 35,5409 | 0,00080638 | 0,02705479 | 1,458479113 | 0,97641821 | COVID | MOCK | Ubiquitin-40S ribosomal protein S27a OS=Homo sapiens OX=9606 GN=RPS27A PE=1 SV=2 | 41676,28831 | 28667,73553 | 42761,41597 | 35564,21683 | 36454,50649 | 30876,62238 | 39828,48902 | 42971,67606 | 26042,81847 | 23306,36369 | 28225,13014 | 29681,27478 | 21237,40913 | 16799,90355 | 29997,62238 | 29581,0787 |
| Q6S8J3 | 36 | 0 | 418,9712 | 0,00098182 | 0,02705479 | 1,232127487 | 0,9707193 | COVID | MOCK | POTE ankyrin domain family member E OS=Homo sapiens OX=9606 GN=POTEE PE=2 SV=3 | 3751357,877 | 4699442,64 | 4456789,956 | 4657394,165 | 4295345,149 | 4728669,42 | 4848515,109 | 5429772,797 | 3371378,375 | 3631807,658 | 4170925,32 | 4346838,336 | 3523837,19 | 3354082,416 | 3780919,025 | 3741861,438 |
| P0DPH7 | 23 | 0 | 260,0714 | 0,00100742 | 0,02705479 | 1,184347997 | 0,96989783 | COVID | MOCK | Tubulin alpha-3C chain OS=Homo sapiens OX=9606 GN=TUBA3C PE=1 SV=1 | 777387,8438 | 682686,9675 | 616071,6074 | 596569,9042 | 656432,5191 | 705190,0686 | 610840,4854 | 673531,2524 | 590377,1864 | 606864,3065 | 514003,9418 | 531566,7595 | 562090,6661 | 627786,7966 | 534499,8488 | 523644,8273 |
| Q9UHD8 | 38 | 19 | 280,351 | 0,00101393 | 0,02705479 | 1,234467473 | 0,96968958 | COVID | MOCK | Septin-9 OS=Homo sapiens OX=9606 GN=SEPTIN9 PE=1 SV=2 | 628193,1293 | 718505,1867 | 652531,2026 | 631115,6367 | 567911,5609 | 668264,606 | 618287,7287 | 706833,0897 | 514188,8197 | 535344,4892 | 581428,5582 | 652044,6301 | 467552,9593 | 431576,2272 | 506939,1394 | 516497,4225 |
| P35579 | 176 | 102 | 1618,8482 | 0,00107264 | 0,02747645 | 1,226977011 | 0,96781804 | COVID | MOCK | Myosin-9 OS=Homo sapiens OX=9606 GN=MYH9 PE=1 SV=4 | 3295653,202 | 4338582,699 | 3410344,535 | 3719157,129 | 3754152,415 | 4694706,067 | 3866079,401 | 4085328,36 | 3093208,255 | 3312217,065 | 2948161,479 | 2980121,174 | 2911394,416 | 3577930,173 | 3264679,459 | 3311299,957 |
| P59768 | 1 | 0 | 12,3138 | 0,00120317 | 0,02932251 | 1,415857553 | 0,96371096 | COVID | MOCK | Guanine nucleotide-binding protein G(I)/G(S)/G(O) subunit gamma-2 OS=Homo sapiens OX=9606 GN=GNG2 PE=1 SV=2 | 3379,829163 | 2386,722248 | 4125,075893 | 3566,921922 | 3440,704039 | 4291,192466 | 2774,255066 | 3583,91598 | 2490,854095 | 2931,335784 | 2103,281606 | 2557,576016 | 2798,638001 | 1947,946873 | 2329,797107 | 2297,76571 |
| P30740 | 13 | 6 | 116,3098 | 0,00129403 | 0,02932251 | 1,188188839 | 0,96089711 | COVID | MOCK | Leukocyte elastase inhibitor OS=Homo sapiens OX=9606 GN=SERPINB1 PE=1 SV=1 | 155761,7634 | 200820,6579 | 175569,5015 | 175414,8677 | 164750,0743 | 183663,5148 | 195217,8345 | 197647,7605 | 146631,1501 | 166957,7446 | 160586,5995 | 166137,9149 | 134467,9812 | 138796,6414 | 150270,1807 | 155525,28 |
| Q96B97 | 36 | 24 | 236,3476 | 0,00132786 | 0,02932251 | 1,174246413 | 0,95985903 | COVID | MOCK | SH3 domain-containing kinase-binding protein 1 OS=Homo sapiens OX=9606 GN=SH3KBP1 PE=1 SV=2 | 196357,1319 | 224841,9056 | 179708,6281 | 182037,3098 | 166215,7196 | 199815,0519 | 195478,3797 | 210025,576 | 160712,7755 | 174079,0958 | 181436,2111 | 171438,3256 | 151929,1085 | 154756,1137 | 164322,3125 | 165136,533 |
| P61978 | 37 | 29 | 332,7659 | 0,00143386 | 0,02962052 | 1,142588997 | 0,95663942 | COVID | MOCK | Heterogeneous nuclear ribonucleoprotein K OS=Homo sapiens OX=9606 GN=HNRNPK PE=1 SV=1 | 732747,1533 | 721679,3045 | 760315,6348 | 722487,1402 | 697903,2421 | 836251,7576 | 710360,719 | 778821,6812 | 571659,2185 | 678008,9057 | 631945,8183 | 612099,906 | 709000,837 | 717383,7312 | 646741,661 | 649879,7119 |
| P49458 | 5 | 3 | 30,8213 | 0,00197593 | 0,0372169 | 1,177455568 | 0,94094072 | COVID | MOCK | Signal recognition particle 9 kDa protein OS=Homo sapiens OX=9606 GN=SRP9 PE=1 SV=2 | 138457,36 | 133398,7262 | 156333,0404 | 151428,9476 | 162273,1572 | 144053,8988 | 144672,9505 | 160093,0923 | 124326,7669 | 110443,9808 | 132335,5425 | 149125,0652 | 124029,3622 | 111657,5184 | 121603,7296 | 137735,8732 |
| P48643 | 26 | 16 | 203,9768 | 0,00206429 | 0,03772822 | 1,180467953 | 0,93849703 | COVID | MOCK | T-complex protein 1 subunit epsilon OS=Homo sapiens OX=9606 GN=CCT5 PE=1 SV=1 | 155247,0566 | 175424,9601 | 164145,7728 | 163335,0581 | 126292,2141 | 175362,6872 | 143815,6084 | 144174,3342 | 128534,6686 | 125796,8803 | 125266,251 | 130718,8283 | 132559,2883 | 135015,2721 | 141547,6475 | 137597,6455 |
| P62834 | 14 | 6 | 85,0996 | 0,00221721 | 0,03772822 | 1,186098079 | 0,93433905 | COVID | MOCK | Ras-related protein Rap-1A OS=Homo sapiens OX=9606 GN=RAP1A PE=1 SV=1 | 103717,3857 | 122948,2522 | 105806,6601 | 111691,4711 | 90180,09023 | 87971,39923 | 103541,2588 | 104997,0019 | 84660,52994 | 88714,43604 | 85786,36452 | 91964,2407 | 75787,71393 | 91951,13843 | 88693,01546 | 92935,65991 |
| Q92945 | 21 | 13 | 150,2832 | 0,00223873 | 0,03772822 | 1,23729403 | 0,93376101 | COVID | MOCK | Far upstream element-binding protein 2 OS=Homo sapiens OX=9606 GN=KHSRP PE=1 SV=4 | 468429,7035 | 440710,1683 | 482380,9079 | 458977,9578 | 365758,2316 | 568870,4231 | 496339,8754 | 546922,4623 | 410240,8938 | 394040,4508 | 384503,7304 | 412510,4486 | 324872,1079 | 375884,8449 | 412477,969 | 379632,8153 |
| P17980 | 16 | 5 | 106,9384 | 0,00233971 | 0,03841898 | 1,504608113 | 0,93107138 | COVID | MOCK | 26S proteasome regulatory subunit 6A OS=Homo sapiens OX=9606 GN=PSMC3 PE=1 SV=3 | 373092,2706 | 542214,9569 | 449508,9461 | 448523,8288 | 293789,5656 | 440914,5929 | 284183,4458 | 315411,0053 | 249231,2071 | 369913,309 | 210481,2807 | 241240,3695 | 233596,5214 | 327626,3619 | 228098,8664 | 231811,0461 |
| P01893 | 8 | 1 | 71,1605 | 0,00242872 | 0,03888346 | 1,258700213 | 0,92873125 | COVID | MOCK | Putative HLA class I histocompatibility antigen_ alpha chain H OS=Homo sapiens OX=9606 GN=HLA-H PE=5 SV=3 | 130036,67 | 147350,9696 | 109612,6261 | 111683,1174 | 100080,1485 | 89267,30087 | 115372,8717 | 128882,1571 | 89918,6876 | 84553,83645 | 98400,32704 | 100750,7661 | 95554,41117 | 93210,85082 | 90255,39615 | 88029,20043 |
| P34931 | 16 | 4 | 148,1187 | 0,00336297 | 0,05127696 | 1,440384814 | 0,90572475 | COVID | MOCK | Heat shock 70 kDa protein 1-like OS=Homo sapiens OX=9606 GN=HSPA1L PE=1 SV=2 | 613897,2864 | 762162,5407 | 516619,3347 | 482835,2732 | 664541,6114 | 743425,3702 | 557467,7741 | 589673,1712 | 391370,4242 | 433836,6143 | 293195,3391 | 282034,6461 | 524728,4425 | 583867,5467 | 472157,3419 | 441938,1829 |
| P39687 | 16 | 4 | 147,848 | 0,00346283 | 0,05157159 | 1,192304153 | 0,90341762 | COVID | MOCK | Acidic leucine-rich nuclear phosphoprotein 32 family member A OS=Homo sapiens OX=9606 GN=ANP32A PE=1 SV=1 | 229941,149 | 263053,0163 | 230959,4876 | 223055,0784 | 207386,3721 | 304668,5256 | 225894,2675 | 231612,7046 | 207274,7447 | 203067,2249 | 192356,3014 | 207480,4146 | 175475,3942 | 192726,7949 | 220075,3442 | 208994,864 |
| Q9UI08 | 22 | 15 | 184,4183 | 0,00478826 | 0,0681417 | 1,282503485 | 0,87506694 | COVID | MOCK | Ena/VASP-like protein OS=Homo sapiens OX=9606 GN=EVL PE=1 SV=2 | 967164,4583 | 1227033,948 | 942327,0916 | 947716,0456 | 945319,141 | 1344442,57 | 854378,0218 | 920236,1124 | 651630,2782 | 960690,9533 | 687111,7896 | 784303,37 | 716353,321 | 914667,6045 | 844736,8844 | 794186,3031 |
| Q01082 | 109 | 62 | 713,211 | 0,00527165 | 0,07182851 | 1,19386631 | 0,86563475 | COVID | MOCK | Spectrin beta chain_ non-erythrocytic 1 OS=Homo sapiens OX=9606 GN=SPTBN1 PE=1 SV=2 | 1946642,492 | 1952736,63 | 2181402,147 | 2083792,256 | 1997277,687 | 2335445,989 | 2567934,96 | 2622841,676 | 1622432,024 | 1838275,094 | 2004901,607 | 1909600,398 | 1680770,245 | 1710254,093 | 2007950,937 | 2041606,685 |
| O43150 | 3 | 1 | 19,1564 | 0,00601495 | 0,07569097 | 1,709832826 | 0,85191734 | COVID | MOCK | Arf-GAP with SH3 domain_ ANK repeat and PH domain-containing protein 2 OS=Homo sapiens OX=9606 GN=ASAP2 PE=1 SV=3 | 77243,34471 | 92022,20136 | 123416,3615 | 118209,8048 | 43014,82588 | 73129,34769 | 97246,41011 | 95116,35719 | 40048,84977 | 30565,77267 | 50237,64009 | 57550,93894 | 42070,03164 | 41417,69724 | 83299,70781 | 75551,46151 |
| P60709 | 59 | 0 | 951,5297 | 0,0060279 | 0,07569097 | 1,114870664 | 0,85168609 | COVID | MOCK | Actin_ cytoplasmic 1 OS=Homo sapiens OX=9606 GN=ACTB PE=1 SV=1 | 13844160,56 | 16312172,35 | 15073220,32 | 14696809,74 | 12759028,1 | 15547237,93 | 14613822,95 | 15464458,45 | 11701320,17 | 13955220,77 | 13336693,5 | 13550650,82 | 12895896,8 | 13567930,51 | 13697825,49 | 13415211,32 |
| P63261 | 90 | 24 | 944,2832 | 0,00665816 | 0,07896023 | 1,11673948 | 0,84072804 | COVID | MOCK | Actin_ cytoplasmic 2 OS=Homo sapiens OX=9606 GN=ACTG1 PE=1 SV=1 | 14349333,89 | 16696858,61 | 15979396,47 | 15641700,96 | 13270763,95 | 15998906,4 | 15619198,66 | 16416483,98 | 12153398,52 | 14463819,44 | 14117408,06 | 14387474,38 | 13325161,1 | 13764217,46 | 14463335,56 | 14338224,05 |
| P25398 | 9 | 4 | 50,2315 | 0,00729274 | 0,07957215 | 1,197071242 | 0,83023638 | COVID | MOCK | 40S ribosomal protein S12 OS=Homo sapiens OX=9606 GN=RPS12 PE=1 SV=3 | 76404,0074 | 58786,96528 | 64374,77164 | 60650,31576 | 70350,26534 | 59676,61039 | 77904,01052 | 82042,92339 | 57766,56439 | 62058,81037 | 59077,46252 | 61630,60039 | 48712,77907 | 52065,67836 | 57150,64491 | 61150,76498 |
| P26641 | 15 | 8 | 113,6057 | 0,00731738 | 0,07957215 | 1,268520512 | 0,82983915 | COVID | MOCK | Elongation factor 1-gamma OS=Homo sapiens OX=9606 GN=EEF1G PE=1 SV=3 | 897289,9105 | 1119031,704 | 787837,7404 | 798279,9899 | 802975,7448 | 1112210,116 | 731048,1802 | 798121,2045 | 577151,1102 | 852612,5669 | 600130,7561 | 676872,9249 | 650688,4469 | 796901,3724 | 732596,5648 | 668174,8711 |
| P32119 | 22 | 18 | 197,7748 | 0,00740759 | 0,07957215 | 1,141914418 | 0,82839112 | COVID | MOCK | Peroxiredoxin-2 OS=Homo sapiens OX=9606 GN=PRDX2 PE=1 SV=5 | 371876,5876 | 433889,9634 | 357143,8547 | 355295,4448 | 325240,5197 | 344744,6159 | 315970,3407 | 338049,0952 | 276292,4795 | 325534,5276 | 300594,2237 | 320613,9679 | 289486,5746 | 332711,9116 | 323278,692 | 320474,8689 |
| O15143 | 21 | 16 | 192,5863 | 0,00764596 | 0,08026943 | 2,888024768 | 0,8246111 | COVID | MOCK | Actin-related protein 2/3 complex subunit 1B OS=Homo sapiens OX=9606 GN=ARPC1B PE=1 SV=3 | 306952,7312 | 676001,2192 | 780127,1689 | 693265,6056 | 151789,4726 | 167085,6374 | 155191,3593 | 168329,7941 | 145514,7545 | 139850,8378 | 147065,4784 | 147483,3683 | 114899,3932 | 105867,2501 | 138647,1257 | 133634,5392 |
| P20273 | 3 | 1 | 20,2496 | 0,00828235 | 0,08211755 | 1,581567453 | 0,81483109 | COVID | MOCK | B-cell receptor CD22 OS=Homo sapiens OX=9606 GN=CD22 PE=1 SV=2 | 9718,059334 | 11565,42695 | 8118,360623 | 8316,335671 | 14324,18764 | 20224,61952 | 15447,36155 | 17954,49769 | 7302,034516 | 7323,338824 | 8233,679103 | 9614,365563 | 6814,403431 | 7301,68712 | 9660,875879 | 10562,35173 |
| Q13404 | 2 | 2 | 13,1345 | 0,00922229 | 0,08814793 | 1,325248709 | 0,80114409 | COVID | MOCK | Ubiquitin-conjugating enzyme E2 variant 1 OS=Homo sapiens OX=9606 GN=UBE2V1 PE=1 SV=2 | 21110,26316 | 27621,7975 | 32232,23084 | 32029,42693 | 28513,26797 | 22752,31803 | 23131,82804 | 28091,77281 | 24158,56718 | 15158,11148 | 22240,86362 | 25891,6404 | 20990,24559 | 13842,19184 | 18401,22164 | 21915,24558 |
| Q16352 | 12 | 4 | 113,1741 | 0,00942101 | 0,08872309 | 1,225826357 | 0,79835659 | COVID | MOCK | Alpha-internexin OS=Homo sapiens OX=9606 GN=INA PE=1 SV=2 | 505279,0893 | 622462,6526 | 512156,7965 | 476334,1413 | 525892,0905 | 578410,7894 | 412827,4441 | 452124,6088 | 421435,1315 | 514631,8097 | 363076,9 | 368606,8671 | 439649,7397 | 491218,3784 | 370208,4776 | 364016,3656 |
| Q16851 | 7 | 4 | 42,2766 | 0,01079407 | 0,09735873 | 1,299360381 | 0,78000047 | COVID | MOCK | UTP--glucose-1-phosphate uridylyltransferase OS=Homo sapiens OX=9606 GN=UGP2 PE=1 SV=5 | 815797,9595 | 1050768,265 | 692252,9318 | 718685,5811 | 713486,1749 | 1018874,607 | 630823,8038 | 693120,1685 | 490743,4406 | 764369,9681 | 503477,3664 | 588520,617 | 569659,4775 | 730782,3431 | 645206,4381 | 581799,8506 |
| A5A3E0 | 39 | 4 | 406,1112 | 0,01166103 | 0,10161901 | 1,228672016 | 0,76914454 | COVID | MOCK | POTE ankyrin domain family member F OS=Homo sapiens OX=9606 GN=POTEF PE=1 SV=2 | 2849422,924 | 3452986,321 | 3647485,387 | 3823027,7 | 3541880,942 | 3692117,707 | 4190957,347 | 4705426,468 | 2840865,563 | 2784349,046 | 3603198,981 | 3693691,496 | 2849702,343 | 2447022,802 | 3043669,965 | 3075406,425 |
| Q562R1 | 31 | 5 | 328,0879 | 0,01174245 | 0,10161901 | 1,161574215 | 0,76815154 | COVID | MOCK | Beta-actin-like protein 2 OS=Homo sapiens OX=9606 GN=ACTBL2 PE=1 SV=2 | 4651778,807 | 5699664,61 | 5715143,768 | 5465787,123 | 4489464,453 | 4325698,456 | 4756932,545 | 5021124,778 | 3781172,656 | 4362294,458 | 4847256,586 | 4713838,486 | 4178618,509 | 3756560,971 | 4274377,907 | 4630030,973 |
| P04083 | 46 | 35 | 464,4802 | 0,01192018 | 0,1017817 | 1,107639909 | 0,76599906 | COVID | MOCK | Annexin A1 OS=Homo sapiens OX=9606 GN=ANXA1 PE=1 SV=2 | 892372,3724 | 1028206,93 | 888495,8844 | 820007,4752 | 820488,6242 | 889386,2694 | 822782,2342 | 875420,783 | 803722,6206 | 801716,08 | 716840,2763 | 725204,2542 | 829494,4301 | 865023,0196 | 829689,5536 | 781602,486 |
| P50570 | 10 | 7 | 51,061 | 0,01386686 | 0,11532815 | 1,784621735 | 0,74367977 | COVID | MOCK | Dynamin-2 OS=Homo sapiens OX=9606 GN=DNM2 PE=1 SV=2 | 101020,0942 | 138966,7257 | 70781,15094 | 64951,45556 | 51239,75854 | 83395,23823 | 43384,91228 | 43352,3538 | 30896,39396 | 49641,64187 | 23708,19504 | 24556,95236 | 56292,87091 | 75369,06971 | 38337,85429 | 35773,07099 |
| P04745 | 15 | 2 | 117,3548 | 0,01464399 | 0,11663779 | 1,800361027 | 0,73535059 | COVID | MOCK | Alpha-amylase 1 OS=Homo sapiens OX=9606 GN=AMY1A PE=1 SV=2 | 61290,00866 | 73466,83985 | 86505,56838 | 74214,87971 | 188814,8275 | 219743,8 | 146041,7742 | 150350,7622 | 66395,5063 | 81461,50125 | 70436,5149 | 68409,204 | 61484,21975 | 59413,65455 | 74924,95125 | 73156,58359 |
| P08865 | 19 | 12 | 153,1392 | 0,01516739 | 0,11663779 | 1,218130159 | 0,72990822 | COVID | MOCK | 40S ribosomal protein SA OS=Homo sapiens OX=9606 GN=RPSA PE=1 SV=4 | 275299,5056 | 298916,1184 | 268321,8882 | 242398,0506 | 171088,042 | 228840,4634 | 212473,4324 | 211298,7161 | 189104,8354 | 220618,7307 | 200465,2349 | 198271,717 | 191310,6091 | 176837,7863 | 195293,2387 | 194955,2177 |
| Q15084 | 4 | 2 | 21,369 | 0,01522801 | 0,11663779 | 1,400318821 | 0,72928622 | COVID | MOCK | Protein disulfide-isomerase A6 OS=Homo sapiens OX=9606 GN=PDIA6 PE=1 SV=1 | 46512,90031 | 53542,89935 | 44624,43189 | 45004,13444 | 36341,00241 | 46641,89515 | 26673,13511 | 28000,10941 | 21453,69902 | 27519,90252 | 25182,42571 | 21712,92102 | 34541,76991 | 40177,14345 | 31806,45258 | 31367,10013 |
| Q8WUM4 | 17 | 12 | 99,4674 | 0,01535834 | 0,11663779 | 1,16255444 | 0,72795471 | COVID | MOCK | Programmed cell death 6-interacting protein OS=Homo sapiens OX=9606 GN=PDCD6IP PE=1 SV=1 | 1676682,141 | 2362107,93 | 1735719,684 | 1837456,685 | 2115809,474 | 2116653,589 | 1795514,82 | 1983000,44 | 1409212,925 | 1769988,926 | 1757810,996 | 1810923,115 | 1762361,238 | 1800753,23 | 1477505,727 | 1649906,792 |
| P10809 | 51 | 40 | 470,3917 | 0,01547498 | 0,11663779 | 1,099511243 | 0,72676957 | COVID | MOCK | 60 kDa heat shock protein_ mitochondrial OS=Homo sapiens OX=9606 GN=HSPD1 PE=1 SV=2 | 780023,4341 | 901481,2526 | 739010,179 | 772141,5097 | 670793,9535 | 811948,978 | 699503,5382 | 742115,9312 | 688447,1505 | 699971,8564 | 677325,3743 | 676338,0233 | 699171,0075 | 724587,1841 | 697935,5806 | 699621,9398 |
| Q9NTX5 | 3 | 2 | 14,4217 | 0,01562799 | 0,11663779 | 1,654219955 | 0,72522428 | COVID | MOCK | Ethylmalonyl-CoA decarboxylase OS=Homo sapiens OX=9606 GN=ECHDC1 PE=1 SV=2 | 17093,45734 | 8317,663488 | 16538,09341 | 16159,79256 | 27627,64607 | 25524,40668 | 24280,56188 | 28728,32691 | 11875,96404 | 13148,4531 | 13904,41836 | 15982,66305 | 10150,86995 | 7692,109487 | 12655,12657 | 13893,96623 |
| P08575 | 54 | 36 | 427,2223 | 0,01566353 | 0,11663779 | 1,187122049 | 0,72486683 | COVID | MOCK | Receptor-type tyrosine-protein phosphatase C OS=Homo sapiens OX=9606 GN=PTPRC PE=1 SV=3 | 1149748,998 | 1410850,579 | 1067696,449 | 1075145,782 | 1183422,817 | 1562377,597 | 1084900,971 | 1180814,439 | 920646,7755 | 1232405,287 | 911527,472 | 1004267,425 | 965025,4662 | 1128174,494 | 1038361,628 | 983213,0477 |
| Q8NE71 | 3 | 2 | 16,7072 | 0,01689551 | 0,12295233 | 1,22704504 | 0,71281037 | COVID | MOCK | ATP-binding cassette sub-family F member 1 OS=Homo sapiens OX=9606 GN=ABCF1 PE=1 SV=2 | 8273,192884 | 9742,883953 | 13148,93494 | 13345,3988 | 13549,06864 | 10591,93208 | 11508,84942 | 13064,86824 | 11318,80331 | 9214,395406 | 9265,998771 | 10012,19146 | 8203,049225 | 8874,996999 | 9156,649781 | 9929,226406 |
| Q15435 | 4 | 2 | 27,0963 | 0,0176006 | 0,12523709 | 1,518934467 | 0,70618627 | COVID | MOCK | Protein phosphatase 1 regulatory subunit 7 OS=Homo sapiens OX=9606 GN=PPP1R7 PE=1 SV=1 | 86834,31681 | 84403,69122 | 60240,8137 | 63509,48145 | 70789,11649 | 99528,13243 | 45137,38882 | 48162,90954 | 44095,88391 | 57443,8902 | 31235,43268 | 29798,68507 | 61758,06471 | 77943,02779 | 36505,54482 | 28981,124 |
| Q9NY65 | 18 | 2 | 216,5365 | 0,01781942 | 0,12540079 | 1,144695596 | 0,70416891 | COVID | MOCK | Tubulin alpha-8 chain OS=Homo sapiens OX=9606 GN=TUBA8 PE=1 SV=1 | 451067,9608 | 369314,5575 | 325033,2774 | 296043,2228 | 380979,8822 | 384161,021 | 349989,3815 | 374500,5303 | 330717,7037 | 304353,6827 | 295181,7797 | 308976,3606 | 326984,5561 | 340822,0717 | 343360,8963 | 310187,4772 |
| Q71U36 | 30 | 5 | 308,0614 | 0,01918328 | 0,12850342 | 1,380923215 | 0,69198092 | COVID | MOCK | Tubulin alpha-1A chain OS=Homo sapiens OX=9606 GN=TUBA1A PE=1 SV=1 | 1143084,293 | 898620,1324 | 582407,8955 | 520944,8681 | 581337,6107 | 655761,2646 | 508033,3859 | 546041,5921 | 519253,4432 | 551735,2449 | 453794,6219 | 439571,787 | 507680,6407 | 563780,8319 | 457346,4281 | 443501,2436 |
| P45880 | 9 | 5 | 62,1734 | 0,01990293 | 0,13005855 | 1,328083535 | 0,68580201 | COVID | MOCK | Voltage-dependent anion-selective channel protein 2 OS=Homo sapiens OX=9606 GN=VDAC2 PE=1 SV=2 | 601863,0985 | 1012914,217 | 558148,1384 | 591286,72 | 706194,5381 | 946121,5646 | 619412,8982 | 669023,2231 | 384257,7364 | 702172,5154 | 450222,1665 | 523882,3776 | 528056,0382 | 689345,5099 | 524259,6696 | 493440,7052 |
| Q15233 | 28 | 19 | 163,4509 | 0,02113569 | 0,13227898 | 1,290552795 | 0,67558976 | COVID | MOCK | Non-POU domain-containing octamer-binding protein OS=Homo sapiens OX=9606 GN=NONO PE=1 SV=4 | 269742,1494 | 302403,1488 | 285996,432 | 298151,7654 | 298282,1048 | 438141,008 | 431169,3765 | 462900,3651 | 244064,2827 | 235437,3905 | 227165,4761 | 236754,9376 | 319324,8682 | 291866,8472 | 309405,3326 | 295355,0191 |
| P06576 | 44 | 28 | 394,1804 | 0,02163812 | 0,13227898 | 1,094743859 | 0,67155464 | COVID | MOCK | ATP synthase subunit beta_ mitochondrial OS=Homo sapiens OX=9606 GN=ATP5F1B PE=1 SV=3 | 931063,9262 | 1029853,055 | 846339,4553 | 836614,8904 | 861480,9422 | 865260,47 | 859405,5487 | 933480,99 | 776088,465 | 830696,0649 | 715624,9188 | 795719,9667 | 839052,7804 | 899591,6321 | 831591,2641 | 855174,0402 |
| Q9H0U4 | 10 | 1 | 85,5639 | 0,02168828 | 0,13227898 | 1,252090971 | 0,67115564 | COVID | MOCK | Ras-related protein Rab-1B OS=Homo sapiens OX=9606 GN=RAB1B PE=1 SV=1 | 172855,5825 | 193420,4731 | 150842,9235 | 143897,8821 | 213463,6233 | 275898,2655 | 204953,184 | 218715,6625 | 147956,8101 | 176579,3214 | 148952,1362 | 141717,7991 | 167686,3892 | 169938,5733 | 158300,6051 | 146003,5359 |
| P52597 | 27 | 16 | 178,018 | 0,02215027 | 0,13349417 | 1,100850729 | 0,66751325 | COVID | MOCK | Heterogeneous nuclear ribonucleoprotein F OS=Homo sapiens OX=9606 GN=HNRNPF PE=1 SV=3 | 164069,3978 | 175882,5626 | 176811,927 | 164810,9296 | 170964,7004 | 190722,2981 | 188292,6484 | 210796,0263 | 168402,1694 | 150751,3429 | 162256,2015 | 151532,1566 | 166949,9839 | 158555,0707 | 176758,9688 | 175008,5177 |
| Q16666 | 7 | 3 | 43,6495 | 0,02251324 | 0,13349417 | 2,180701955 | 0,6646915 | COVID | MOCK | Gamma-interferon-inducible protein 16 OS=Homo sapiens OX=9606 GN=IFI16 PE=1 SV=3 | 53610,48069 | 72469,23731 | 51691,35778 | 47946,44238 | 210017,4665 | 226688,3834 | 136061,8042 | 139717,0203 | 46919,62045 | 65558,08591 | 36078,7602 | 36163,04682 | 55045,11586 | 98297,28586 | 45907,95397 | 46259,57635 |
| Q14498 | 4 | 1 | 21,4073 | 0,02367617 | 0,13457303 | 1,346780757 | 0,6558779 | COVID | MOCK | RNA-binding protein 39 OS=Homo sapiens OX=9606 GN=RBM39 PE=1 SV=2 | 62818,19929 | 63242,04563 | 84407,43062 | 74004,33978 | 73343,1661 | 76216,65885 | 88198,37039 | 91150,10441 | 54987,56574 | 51906,89711 | 71795,06703 | 67553,33777 | 51814,26502 | 24842,12057 | 65997,81168 | 66544,78177 |
| P61158 | 40 | 26 | 333,0327 | 0,02374589 | 0,13457303 | 1,397646282 | 0,65536013 | COVID | MOCK | Actin-related protein 3 OS=Homo sapiens OX=9606 GN=ACTR3 PE=1 SV=3 | 707519,5436 | 853954,3476 | 874472,1305 | 814204,0539 | 1567955,311 | 1162953,034 | 1466305,86 | 1537644,784 | 779878,3575 | 841692,5242 | 770569,9974 | 807900,4989 | 777835,1163 | 758351,3667 | 870399,3421 | 822044,4724 |
| Q8IYD8 | 6 | 3 | 41,6171 | 0,02533053 | 0,1422943 | 1,197812076 | 0,64389643 | COVID | MOCK | Fanconi anemia group M protein OS=Homo sapiens OX=9606 GN=FANCM PE=1 SV=2 | 23531,36007 | 25515,36926 | 30074,31743 | 28111,88098 | 24702,49855 | 28269,60597 | 30851,08928 | 27262,30053 | 22953,88335 | 25124,3524 | 28353,32649 | 29067,55901 | 18243,02778 | 16904,6267 | 20098,7634 | 21518,79638 |
| P28838 | 10 | 5 | 54,3367 | 0,02717169 | 0,15130971 | 1,311382225 | 0,63126005 | COVID | MOCK | Cytosol aminopeptidase OS=Homo sapiens OX=9606 GN=LAP3 PE=1 SV=3 | 128509,4033 | 116605,5776 | 143212,526 | 129134,4792 | 81645,67121 | 83188,4706 | 80081,13277 | 81453,44243 | 80578,87098 | 100781,9193 | 83837,58442 | 93166,53405 | 72715,30746 | 64983,854 | 72239,33359 | 75163,22756 |
| Q92688 | 10 | 4 | 127,5564 | 0,02784357 | 0,15371454 | 1,205406188 | 0,62681721 | COVID | MOCK | Acidic leucine-rich nuclear phosphoprotein 32 family member B OS=Homo sapiens OX=9606 GN=ANP32B PE=1 SV=1 | 174987,2543 | 188883,803 | 170284,0122 | 157463,4255 | 199628,9497 | 292365,429 | 202473,3465 | 207501,2149 | 168034,3257 | 183669,2839 | 154426,672 | 156854,2402 | 166608,4521 | 177952,139 | 158502,968 | 155985,4796 |
| Q9BQE3 | 23 | 0 | 293,4555 | 0,0284492 | 0,15571563 | 1,341248564 | 0,6228852 | COVID | MOCK | Tubulin alpha-1C chain OS=Homo sapiens OX=9606 GN=TUBA1C PE=1 SV=1 | 1050640,549 | 800236,3853 | 532860,5822 | 477023,2118 | 555570,4422 | 606249,6739 | 469314,4327 | 507537,4133 | 498445,8547 | 512300,0986 | 419718,9065 | 404787,6521 | 488572,4329 | 538076,727 | 440343,3612 | 425201,7822 |
| P49903 | 7 | 3 | 45,2622 | 0,02931114 | 0,15907384 | 1,192599598 | 0,61740334 | COVID | MOCK | Selenide_ water dikinase 1 OS=Homo sapiens OX=9606 GN=SEPHS1 PE=1 SV=2 | 89074,25505 | 121703,8904 | 95564,40591 | 97817,36703 | 81676,74923 | 109589,2721 | 108417,0072 | 106599,859 | 79933,32208 | 70513,63262 | 90927,78269 | 112986,6468 | 69262,43749 | 73047,53911 | 94275,50152 | 88612,98942 |
| Q9UKY7 | 5 | 3 | 36,0884 | 0,03046029 | 0,16223784 | 1,266378911 | 0,61029401 | COVID | MOCK | Protein CDV3 homolog OS=Homo sapiens OX=9606 GN=CDV3 PE=1 SV=1 | 18086,3195 | 18994,65332 | 22204,5968 | 18302,02639 | 16566,54264 | 13072,44935 | 13364,5845 | 13773,53897 | 14978,6765 | 13170,06331 | 13007,74396 | 14012,6384 | 13408,79842 | 8129,619987 | 14645,09728 | 14748,86965 |
| P21796 | 19 | 8 | 127,4355 | 0,03151836 | 0,16409924 | 1,248754067 | 0,60393883 | COVID | MOCK | Voltage-dependent anion-selective channel protein 1 OS=Homo sapiens OX=9606 GN=VDAC1 PE=1 SV=2 | 370035,0907 | 355629,6095 | 279436,605 | 265406,822 | 281002,1958 | 295092,3206 | 211154,1134 | 232699,3743 | 230362,897 | 237352,0647 | 185244,8336 | 184005,7853 | 285670,2422 | 294020,9922 | 203699,362 | 213836,9532 |
| Q8N163 | 13 | 8 | 90,3861 | 0,03307262 | 0,16479976 | 1,11337356 | 0,59491428 | COVID | MOCK | Cell cycle and apoptosis regulator protein 2 OS=Homo sapiens OX=9606 GN=CCAR2 PE=1 SV=2 | 975592,6562 | 1076213,848 | 1020755,93 | 956003,34 | 965341,5955 | 1174577,933 | 878863,181 | 911714,4896 | 797608,4799 | 881195,7409 | 833508,7978 | 858451,7058 | 850752,009 | 1048549,944 | 971712,047 | 906821,9445 |
| P22314 | 46 | 24 | 400,1482 | 0,03309453 | 0,16479976 | 1,31527336 | 0,5947896 | COVID | MOCK | Ubiquitin-like modifier-activating enzyme 1 OS=Homo sapiens OX=9606 GN=UBA1 PE=1 SV=3 | 1730862,12 | 2954183,867 | 1835519,565 | 1854094,185 | 1530605,101 | 2069582,241 | 1309146,751 | 1338410,563 | 1149882,852 | 1646299,19 | 1204778,279 | 1261494,484 | 1414978,959 | 1774583,819 | 1339864,106 | 1325506,407 |
| Q8WYJ6 | 20 | 11 | 173,3209 | 0,03342316 | 0,16479976 | 1,104721445 | 0,59292762 | COVID | MOCK | Septin-1 OS=Homo sapiens OX=9606 GN=SEPTIN1 PE=1 SV=2 | 338832,5129 | 395839,1686 | 366788,9102 | 347296,9284 | 460671,9953 | 416065,3219 | 394510,1493 | 424766,977 | 375350,188 | 338361,8044 | 358284,3332 | 372351,3266 | 376414,4884 | 344104,0741 | 335649,3593 | 346149,5087 |
| P68363 | 24 | 0 | 312,7716 | 0,03377226 | 0,16479976 | 1,331929291 | 0,59096627 | COVID | MOCK | Tubulin alpha-1B chain OS=Homo sapiens OX=9606 GN=TUBA1B PE=1 SV=1 | 1213093,142 | 998225,6061 | 642279,3648 | 571630,5043 | 669007,5281 | 794322,9338 | 561121,8327 | 611150,8136 | 596856,8977 | 659260,7781 | 489206,5785 | 484621,8636 | 609429,424 | 675817,0192 | 529899,5467 | 505323,4185 |
| P14174 | 7 | 3 | 42,5398 | 0,03401324 | 0,16479976 | 1,234429848 | 0,58962222 | COVID | MOCK | Macrophage migration inhibitory factor OS=Homo sapiens OX=9606 GN=MIF PE=1 SV=4 | 225692,0413 | 275154,2949 | 284523,2368 | 272198,7239 | 179137,8745 | 173159,0948 | 190067,9725 | 193001,9655 | 174568,9807 | 142790,9125 | 189613,9143 | 212983,8635 | 175905,9999 | 174297,597 | 185758,5113 | 196520,153 |
| P23368 | 4 | 2 | 20,7732 | 0,03428531 | 0,16479976 | 1,579693626 | 0,5881143 | COVID | MOCK | NAD-dependent malic enzyme_ mitochondrial OS=Homo sapiens OX=9606 GN=ME2 PE=1 SV=1 | 11245,10696 | 10793,30324 | 9398,282234 | 8874,760838 | 23569,00316 | 24303,04203 | 19834,46943 | 23118,9073 | 10905,84331 | 11686,38207 | 10504,17425 | 11148,53507 | 10419,76799 | 10119,92894 | 8963,618091 | 9265,869608 |
| P62277 | 8 | 5 | 50,2149 | 0,034741 | 0,16479976 | 1,35177204 | 0,58561096 | COVID | MOCK | 40S ribosomal protein S13 OS=Homo sapiens OX=9606 GN=RPS13 PE=1 SV=2 | 58158,21822 | 55320,12931 | 65454,14585 | 63613,69958 | 98034,23892 | 128411,9638 | 83659,70167 | 90299,19407 | 58526,0614 | 62841,42511 | 53891,98505 | 48714,3969 | 58395,22423 | 80964,20017 | 59145,83686 | 53156,75572 |
| Q15365 | 20 | 13 | 176,9771 | 0,03754271 | 0,17278317 | 1,292818956 | 0,57080159 | COVID | MOCK | Poly(rC)-binding protein 1 OS=Homo sapiens OX=9606 GN=PCBP1 PE=1 SV=2 | 641682,2022 | 739651,7031 | 521645,4592 | 503023,686 | 488073,6767 | 533508,5185 | 404660,9973 | 460787,4056 | 360109,9337 | 396127,6807 | 289605,5109 | 258462,222 | 521291,8151 | 566346,3438 | 476029,1899 | 452703,898 |
| P62701 | 18 | 15 | 136,8069 | 0,03904681 | 0,17613563 | 1,353533902 | 0,56323586 | COVID | MOCK | 40S ribosomal protein S4_ X isoform OS=Homo sapiens OX=9606 GN=RPS4X PE=1 SV=2 | 256780,7597 | 338485,6896 | 218937,4591 | 214694,7951 | 152432,4126 | 176413,1052 | 147488,8842 | 168748,375 | 201232,0964 | 151315,9284 | 119640,3592 | 110101,0701 | 172798,0181 | 192020,4231 | 147112,0037 | 142528,939 |
| P24752 | 12 | 5 | 100,4992 | 0,04000049 | 0,17913372 | 1,406229246 | 0,55856777 | COVID | MOCK | Acetyl-CoA acetyltransferase_ mitochondrial OS=Homo sapiens OX=9606 GN=ACAT1 PE=1 SV=1 | 1834013,265 | 2201132,025 | 1431992,012 | 1826442,771 | 1232187,843 | 1799985,065 | 1120571,26 | 1174705,414 | 819381,6786 | 1100345,022 | 535918,0165 | 718929,1212 | 1144897,186 | 1866676,351 | 1367415,496 | 1421524,009 |
| P60174 | 53 | 42 | 406,8849 | 0,04047723 | 0,18000987 | 1,104558399 | 0,55627027 | COVID | MOCK | Triosephosphate isomerase OS=Homo sapiens OX=9606 GN=TPI1 PE=1 SV=3 | 977234,4623 | 1300513,164 | 1115393,564 | 1049992,205 | 934824,2555 | 1106745,265 | 1042529,447 | 1140127,482 | 929552,0374 | 1048051,125 | 903365,3953 | 911475,4659 | 951160,0841 | 1074645,177 | 1023102,182 | 1005549,021 |
| P60891 | 9 | 1 | 58,0364 | 0,04174758 | 0,18311608 | 1,160999636 | 0,55026116 | COVID | MOCK | Ribose-phosphate pyrophosphokinase 1 OS=Homo sapiens OX=9606 GN=PRPS1 PE=1 SV=2 | 114123,0961 | 130313,4693 | 118216,5436 | 121754,7978 | 151206,2291 | 168624,2856 | 138739,1515 | 157390,7155 | 138389,7137 | 103907,3563 | 130119,3185 | 133874,2261 | 109856,546 | 106210,541 | 106058,5802 | 119360,3185 |
| P81605 | 3 | 2 | 22,2086 | 0,04338902 | 0,18599542 | 1,149751767 | 0,54272941 | COVID | MOCK | Dermcidin OS=Homo sapiens OX=9606 GN=DCD PE=1 SV=2 | 23382,38833 | 23552,17448 | 24653,73183 | 23758,3605 | 21878,31072 | 21255,93096 | 23444,62345 | 22564,97078 | 15906,65116 | 20427,85681 | 20362,43125 | 22883,34114 | 17289,13742 | 14943,90509 | 23848,07663 | 24799,75073 |
| P00367 | 41 | 13 | 283,2415 | 0,04390818 | 0,1862158 | 1,788003476 | 0,54039943 | COVID | MOCK | Glutamate dehydrogenase 1_ mitochondrial OS=Homo sapiens OX=9606 GN=GLUD1 PE=1 SV=2 | 968540,8457 | 959163,494 | 1392388,564 | 1437456,49 | 384233,3431 | 465574,0576 | 399895,828 | 449281,2536 | 451808,0553 | 439371,0455 | 446993,2118 | 455098,2681 | 442397,8293 | 393430,0085 | 496271,8192 | 485659,5843 |
| P14649 | 11 | 6 | 88,8707 | 0,04649787 | 0,19167416 | 1,170217654 | 0,52912995 | COVID | MOCK | Myosin light chain 6B OS=Homo sapiens OX=9606 GN=MYL6B PE=1 SV=1 | 173116,9087 | 176885,5126 | 250393,6054 | 258665,675 | 178576,0011 | 242361,2018 | 228879,367 | 213225,5595 | 159299,3314 | 163166,4164 | 187697,4193 | 186124,6999 | 173094,3142 | 190331,2338 | 213073,0239 | 198823,4158 |
| P10515 | 5 | 3 | 26,1554 | 0,04689518 | 0,19167416 | 1,257549527 | 0,52745084 | COVID | MOCK | Dihydrolipoyllysine-residue acetyltransferase component of pyruvate dehydrogenase complex_ mitochondrial OS=Homo sapiens OX=9606 GN=DLAT PE | 18483,36828 | 20908,1491 | 13145,00447 | 13481,11421 | 16649,85615 | 23108,98728 | 14418,46515 | 16967,55759 | 15404,64878 | 18584,2808 | 11331,06677 | 10986,92149 | 14201,49552 | 16902,47825 | 11409,54231 | 10250,81878 |
| P07737 | 49 | 34 | 339,1714 | 0,04702137 | 0,19167416 | 1,13482732 | 0,52692017 | COVID | MOCK | Profilin-1 OS=Homo sapiens OX=9606 GN=PFN1 PE=1 SV=2 | 2036389,044 | 2757335,013 | 2102745,404 | 1998344,796 | 2058768,038 | 2698821,534 | 2388825,064 | 2490393,065 | 1848186,719 | 2363947,073 | 1950516,831 | 2107607,842 | 1870783,917 | 2263021,148 | 1971235,853 | 1954605,288 |
| Q9Y230 | 5 | 1 | 32,7822 | 0,04752779 | 0,19167416 | 1,237366708 | 0,52480341 | COVID | MOCK | RuvB-like 2 OS=Homo sapiens OX=9606 GN=RUVBL2 PE=1 SV=3 | 215624,0591 | 187451,2643 | 204568,2046 | 255668,5041 | 190406,3103 | 183684,8576 | 295351,8766 | 312577,4913 | 153229,135 | 161582,69 | 204544,6068 | 248774,2199 | 162414,364 | 171372,4715 | 182143,2441 | 207277,735 |
| P62736 | 94 | 47 | 556,8248 | 0,04788645 | 0,19167416 | 1,164452606 | 0,52331653 | COVID | MOCK | Actin_ aortic smooth muscle OS=Homo sapiens OX=9606 GN=ACTA2 PE=1 SV=1 | 5013047,381 | 6249116,504 | 6954903,518 | 7136512,452 | 6329603,392 | 5719404,505 | 7288421,431 | 7867563,114 | 5182439,149 | 4779017,906 | 6316662,808 | 6793629,351 | 5274238,084 | 4788640,015 | 5831549,022 | 6169685,799 |
| P17987 | 18 | 9 | 101,683 | 0,04788897 | 0,19167416 | 1,139113848 | 0,5233061 | COVID | MOCK | T-complex protein 1 subunit alpha OS=Homo sapiens OX=9606 GN=TCP1 PE=1 SV=1 | 97007,62767 | 87354,00533 | 97626,36217 | 94172,36642 | 100548,4152 | 108094,2586 | 125300,5894 | 134331,1427 | 93476,86464 | 95690,26095 | 101404,2327 | 96140,85467 | 85551,66424 | 85028,08267 | 94462,65567 | 89553,87496 |
| Q9UBT2 | 4 | 2 | 24,3589 | 0,05030579 | 0,2000968 | 1,234141048 | 0,51354186 | COVID | MOCK | SUMO-activating enzyme subunit 2 OS=Homo sapiens OX=9606 GN=UBA2 PE=1 SV=2 | 50839,672 | 39076,0294 | 37355,84841 | 32218,40943 | 26000,28017 | 31592,24326 | 27378,50851 | 27890,90966 | 25430,37178 | 28150,88512 | 24237,92112 | 24487,77794 | 27856,78243 | 35939,55887 | 26685,40538 | 27892,63787 |
