## Supplementary material for "SARS-CoV-2 Uses CD4 to Infect T Helper Lymphocytes": Table S3

### Supplementary Table 3

| Antibody | Fluorochrome | Clone | Manufacture Catalogue |
|---|---|---|---|
| Anti-CD4 (Human) Monoclonal | FITC | RPA-T4 | BD Biosciences Cat.: 555346 |
| Anti-CD4 (Human) Monoclonal | APC | SK3 | BD Biosciences Cat.: 566915 |
| Anti-CD4 (Human) Monoclonal | BUV661 | SK3 | BD Biosciences Cat.: 612962 |
| Anti-CD3 (Human) Monoclonal | PercP-Cy5.5 | SP34-2 | BD Biosciences Cat.: 552852 |
| Anti-CD3 (Human) Monoclonal | BV480 | UCHT1 | BD Biosciences Cat.: 51-9015420 |
| Anti-CD8a (Human) Monoclonal | PE-Cy7 | RPA-T8 | eBioscience (ThermoFisher) Cat.: 25-0088-42 |
| Lineage Cocktail 1 (Human)- CD3, CD14, CD16, CD19, CD20, CD56) | FITC | SK7 (CD3), MfP9 (CD14), 3G8 (CD16), SJ25C1 (CD19), L27 (CD20), NCAM16.2 (CD56) | BD Biosciences Cat.: 552852 |
| Zombie Aqua™ Fixable Viability Kit | BUV496 | - | BioLegend Cat.: 423101 |
| Anti-CD14 (Human) Monoclonal | BUV805 | M5E2 | BD Biosciences Cat.: 612902 |
| Anti-CD16 (Human) Monoclonal | BUV496 | 3G8 | BD Biosciences Cat.: 612944 |
| Anti-CD19 (Human) Monoclonal | BUV563 | SJ25C1 | BD Biosciences Cat.: 51-9016630 |
| Anti-CD45 (Human) Monoclonal | APC-Cy7 | 2D1 | BD Biosciences Cat.: 557833 |
| Anti-CD123 (Human) Monoclonal | PE-Cy5 | 9F5 | BD Biosciences Cat.: 51-9015428 |
| Anti-CD4 (Human) Polyclonal | Purified | - | RehaBiotech IM:0566 |
| Anti-ACE2 (Human) Polyclonal | Purified | - | RehaBiotech IM:0060 |
| Anti-ACE2 (Human) Polyclonal | PE | - | RehaBiotech IM:0060 |
| Anti-sCoV-2 | Purified | - | |
| Goat Anti-IgG (Human) Polyclonal | Alexa 488 | | ThermoFisher - A11013 |
| Anti-IgG (Human) | | | |
| Anti-IgG (mouse) | | | |
| Anti-IgG (Rabbit) | Peroxidase | | GE Healthcare: RPN4301 |
| Anti-Spike | - | HC2001 | GeneScript: A02038 |
