## Supplementary material for "SARS-CoV-2 Uses CD4 to Infect T Helper Lymphocytes": Table S4

| Cycle Threshold | | | |
|---|---|---|---|
| Figure 1 A. CD4 and CD8 infection | | | |
| Mock CD4 | CoV-2 CD4 | Mock CD8 | CoV-2 CD8 |
| 36,066 | 29,877 | 35,426 | 33,517 |
| 37,251 | 30,119 | 37,946 | 34,070 |
| 37,119 | 29,449 | 36,066 | 36,416 |
| Undetermined | 29,326 | 35,072 | 36,027 |
| 35,670 | 31,402 | 39,053 | 35,015 |
| 35,464 | 31,821 | 38,385 | 35,516 |
| | | | 36,939 |
| | | | 35,516 |
| | | | 33,464 |
| | | | 33,166 |

| Cycle Threshold | | | | | |
|---|---|---|---|---|---|
| Figure 1 D. Temporal CD4 infection | | | | | |
| Mock | CoV-2 2h | CoV-2 6h | CoV-2 12h | CoV-2 24h | CoV-2 48h |
| 33,245 | 23,202 | 21,556 | 23,728 | 23,352 | 23,725 |
| 33,271 | 23,592 | 21,683 | 23,755 | 24,293 | 23,875 |
| 33,271 | 23,422 | 21,408 | 24,831 | 22,300 | 21,702 |
| 29,962 | 23,619 | 21,331 | 24,555 | 22,827 | 22,423 |
| 33,920 | 22,361 | 22,188 | 23,432 | 21,818 | 25,832 |
| 32,177 | 22,888 | 21,953 | 22,782 | 21,238 | 25,996 |
