## Supplementary figures and images for "SARS-CoV-2 Uses CD4 to Infect T Helper Lymphocytes"

### Full unedited gel for Extended Figure 4D.pdf

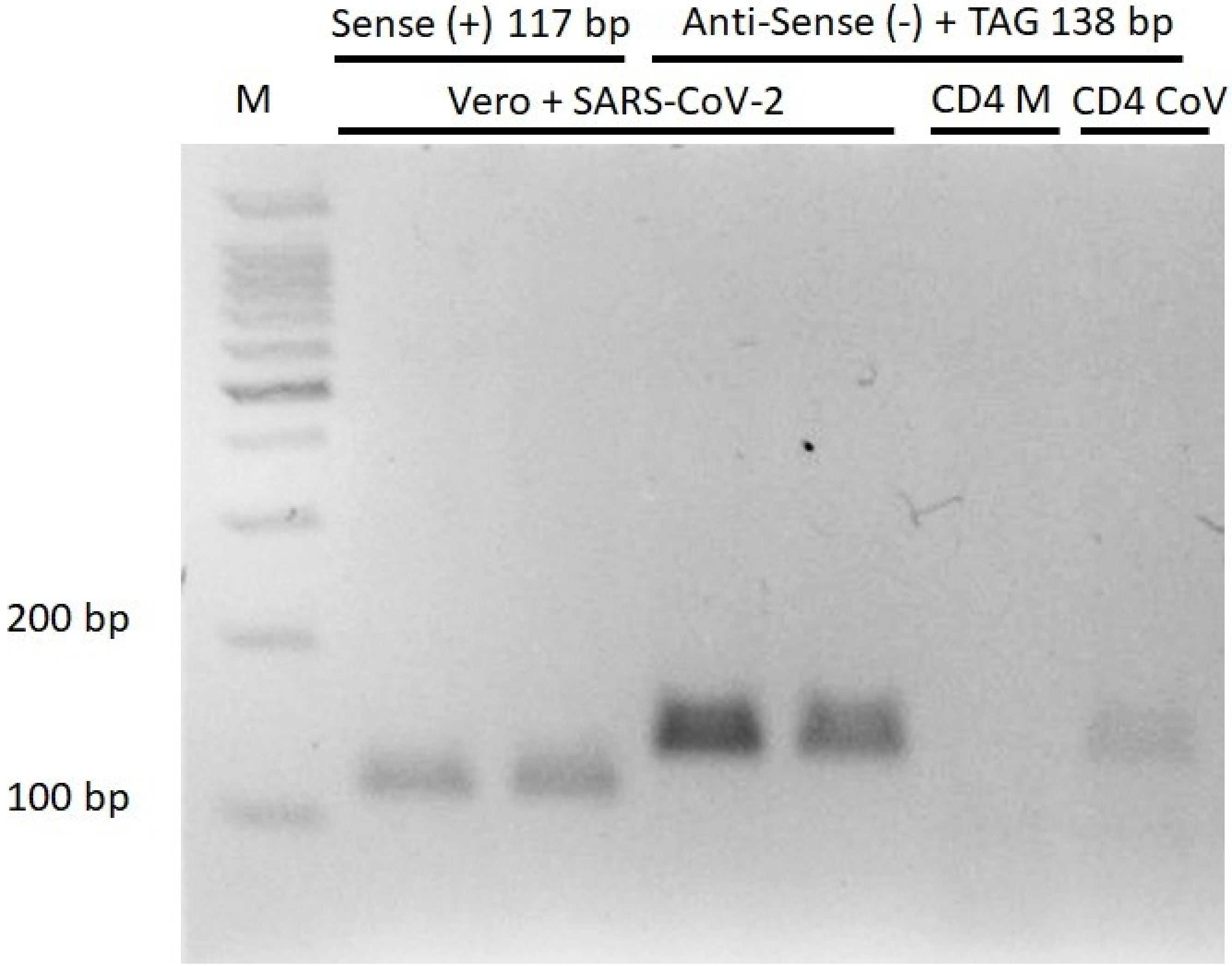

### Full unedited gel for Extended Figure 7A.pdf

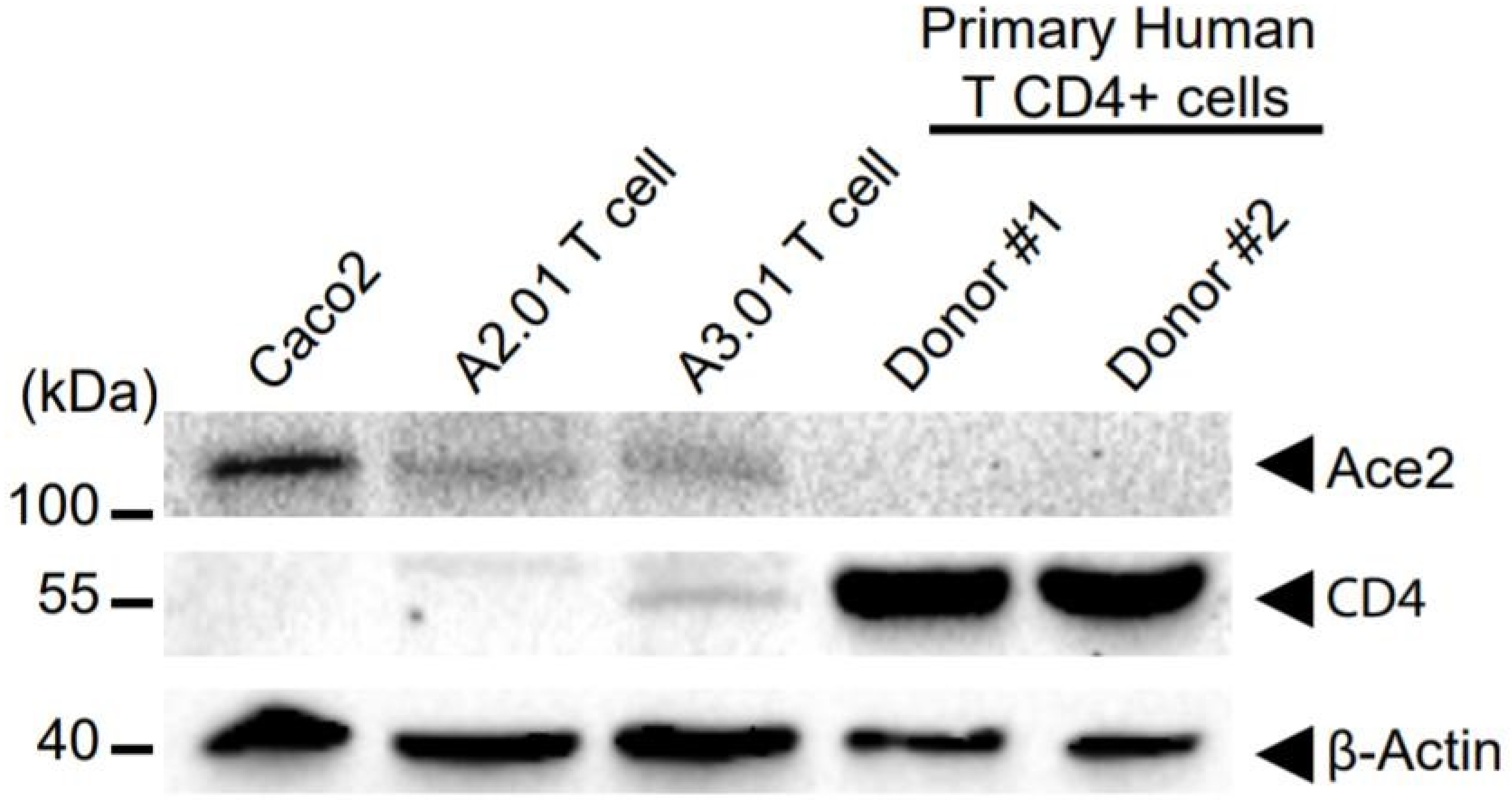

### Full unedited gel for Figure 3A.pdf

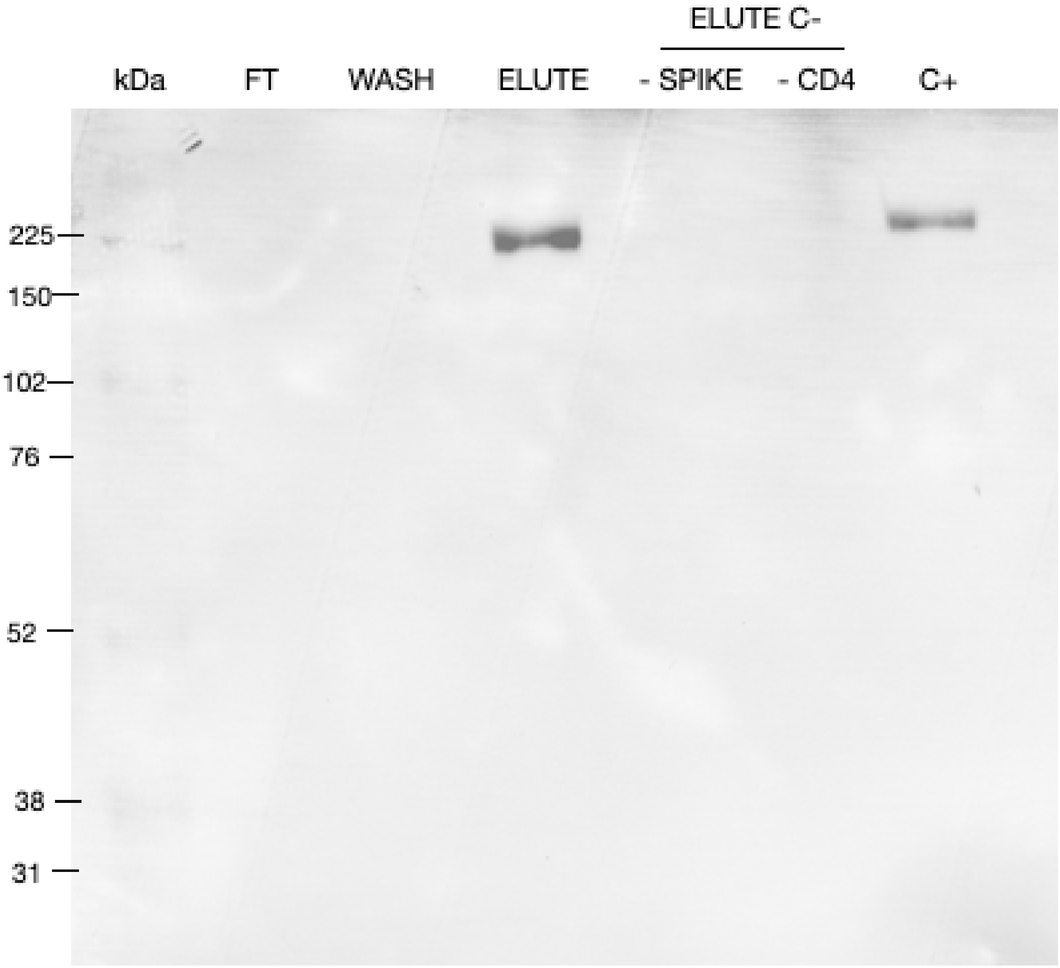


Streptavidin-HRP detection (Biolegend cat. num. 405210).

### Full unedited gel for Figure 3G.pdf

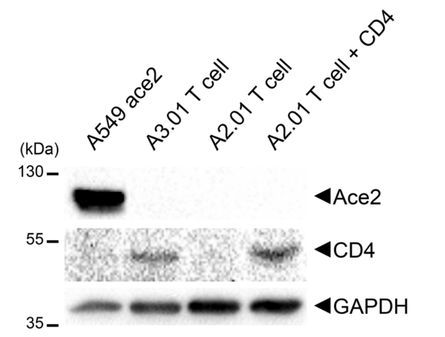

### Full unedited gel for Figure 4E.pdf

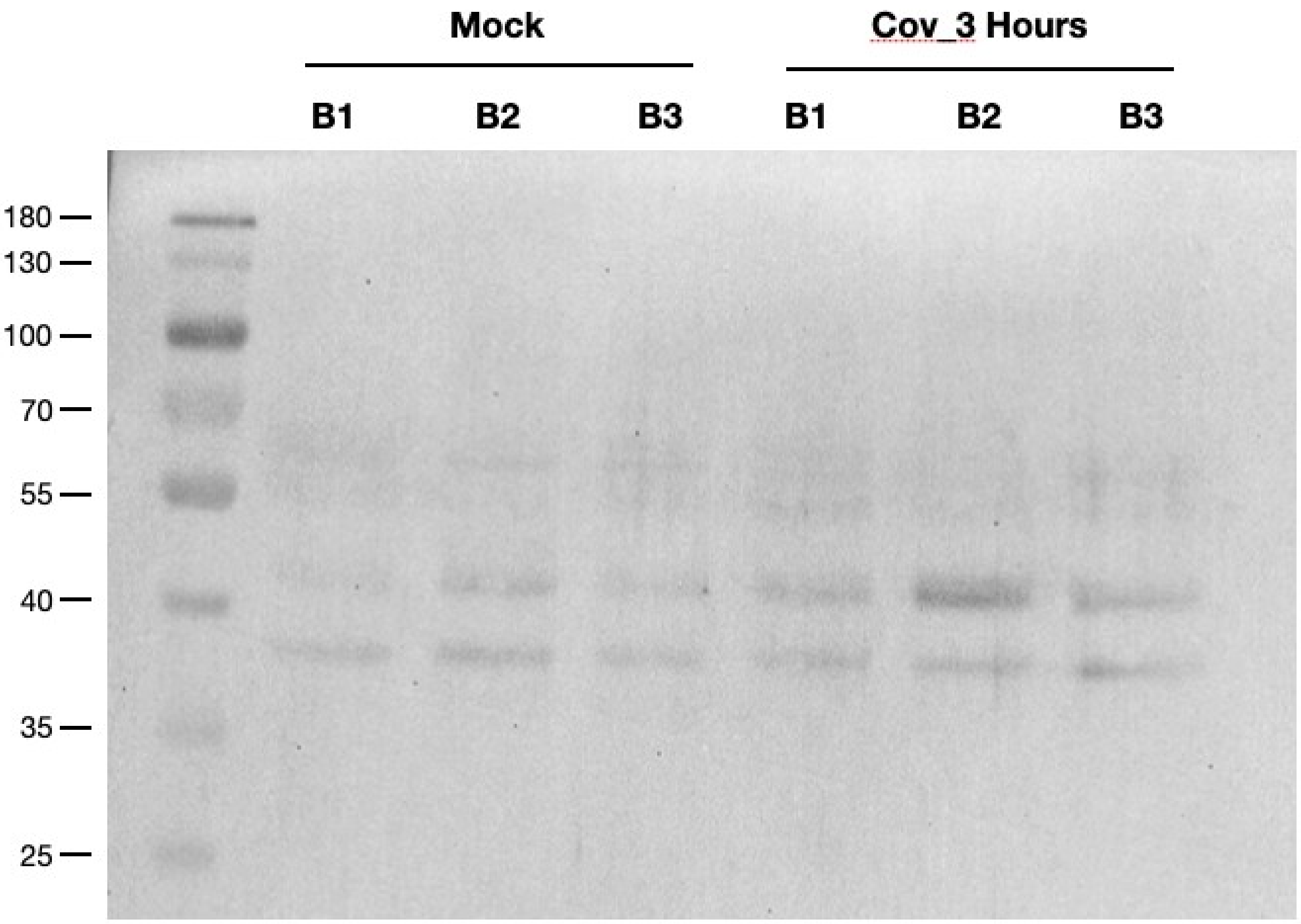


p-CREB_Cell signaling #9198 (ser133)

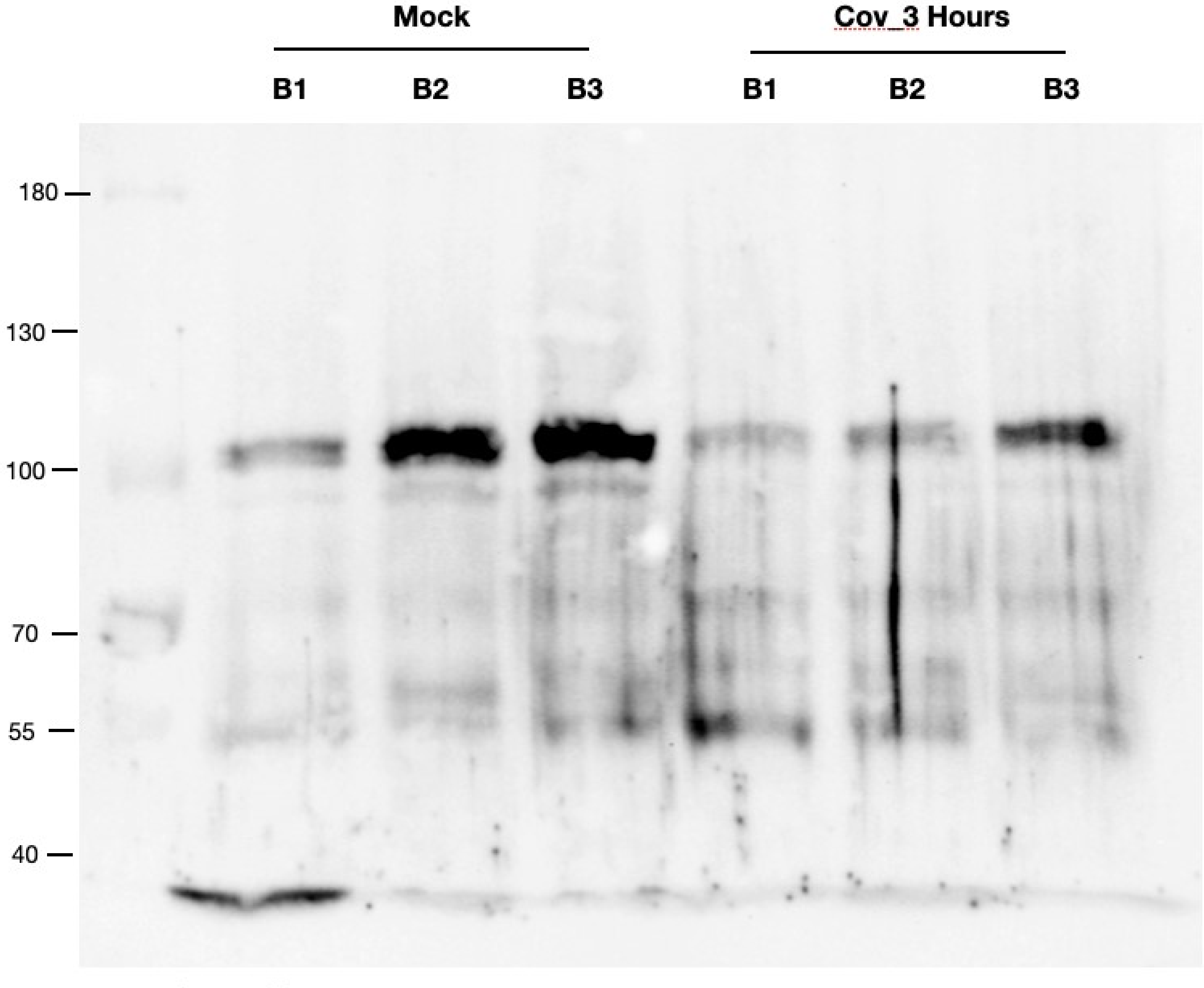

p-STAT3_ SC-8059_1:250

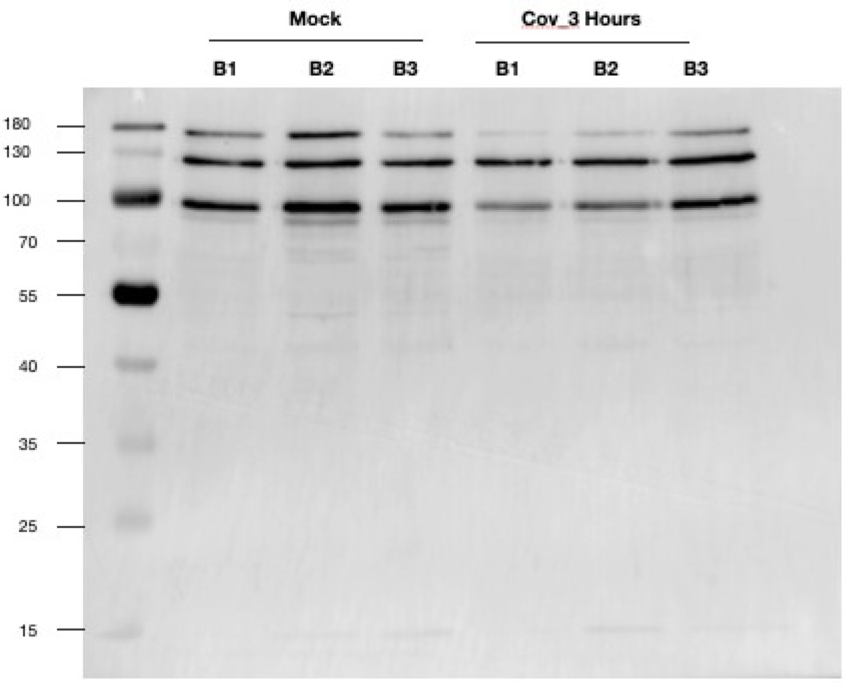


Total STAT3_Cell Signaling # 4904

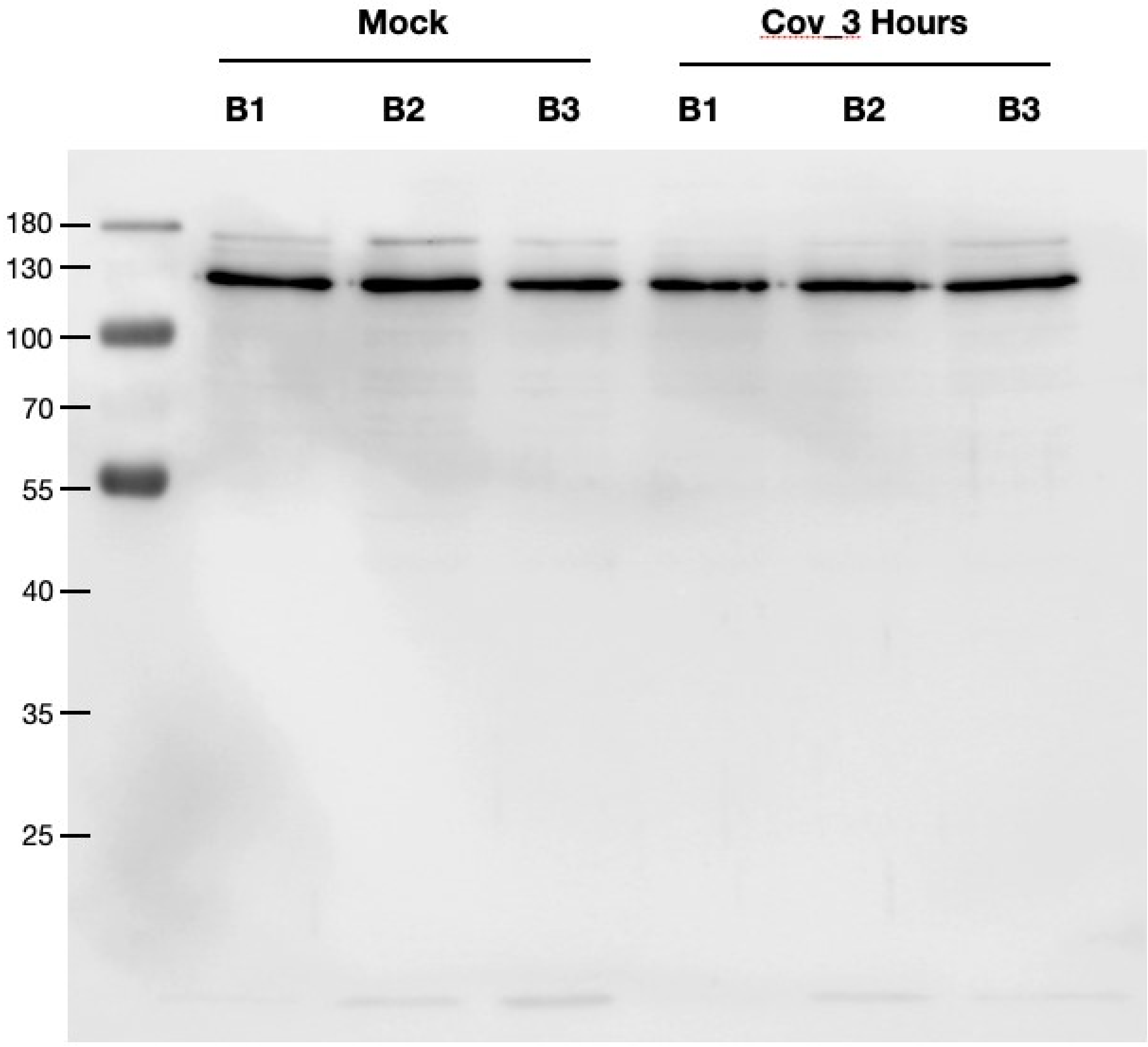


Vinculin_Abcam #18058
